# An allele-based model of coronavirus evolution under population immunity

**DOI:** 10.1101/2025.09.15.25335781

**Authors:** Carl P. Simon, James S. Koopman, Marisa C. Eisenberg, Luis Zaman, Matthew Andres Moreno, Austin Polanco

## Abstract

Most epidemic models represent viral evolution at the level of whole strains, assigning transmissibility and immune escape directly to variants. However, in reality, mutations occur at specific genomic sites, and the epidemiological consequences of viral evolution depend on how these allele-level changes interact with population immunity.

We develop a compartmental model in which viral strains are defined by combinations of alleles across genomic sites, while transmission, immunity, waning, and mutation operate at the allele level. This framework allows evolutionary pressures acting on individual genomic components to propagate across multiple strains.

Even in a minimal system with two sites and two alleles per site, the model generates rich dynamics. Epidemic waves of an initially seeded strain produce an immune landscape that often favors genetically complementary strains. More generally, four mechanisms shape strain success: increases in prevalence of a strain due to increased transmissibility or immunity-avoidance of its underlying alleles, complementarity effects driven by population immunity, founder effects that indirectly favor the initially circulating strain, and adjacency effects associated with mutation pathways.

These results show that immune structure can redirect viral evolution in ways not captured by strain-based models and provide a simple framework linking viral genetics with epidemic dynamics.

## I. Introduction

Coronaviruses evolve through mutations at many genomic sites, and the biological effects of these mutations are often site-specific [Da24, Ing21]. Laboratory studies have identified mutations that alter receptor binding, immune escape, replication, and other viral functions [Li16, Ch23]. Yet most population-level epidemic models represent viral evolution at the level of whole strains or variants, assigning transmissibility and immune escape directly to the strain. For example, Saad-Roy and colleagues have published a series of path-breaking papers [Sa24b, Sa21a, Sa24a, Sa20, Sa23, Wa21, Es24] on the transmissibility and immune-response of SARS-CoV-2, all focused around the infections and reinfections of a single strain.

While useful for short-term variant competition, that approach does not capture how selection acts on the underlying genomic components from which strains are assembled.

Here we develop an alternative framework: a population-level epidemic model in which viral traits are defined at the **allele level**, while transmission dynamics unfold at the **strain level**. A strain is defined by its combination of alleles, and mutation occurs through allele replacement at individual sites.

This formulation allows us to ask how changes in transmissibility, immune escape, waning, or infection duration associated with a single allele propagate across all strains that carry it.

This allele-based perspective is motivated by a broader biological question. The evolutionary trajectory of a coronavirus is not determined by transmissibility alone. It also depends on the immune landscape created by prior infections [Ko22]. As immunity accumulates in the population, strains that were initially disfavored may become selectively advantaged because they encounter less cross-immunity. That is, immune memory can introduce ecological balancing among strains via negative frequency-dependent selection. Thus the relevant evolutionary information is not just the currently dominant strain, but also includes host susceptibility patterns shaped by circulating allelic variation.

Our goal is not to reproduce a specific epidemic, but to identify **general mechanisms** linking mutation, immunity, and transmission. Although motivated by SARS-CoV-2, the framework applies broadly to coronaviruses and other RNA viruses.

Even in a minimal two-site, two-allele system, the model reveals three key phenomena:

- Epidemic waves generate immune landscapes favoring complementary strains
- Initial conditions produce persistent founder effects
- Immune structure determines whether evolution proceeds locally or via larger jumps

These results show that **population immunity is not just a constraint—it dynamically shapes evolutionary pressures**.

Our model focuses on the virus characteristics. So, as a first step, all hosts are treated as identical. We model processes that are biologically rather than sociologically determined and ignore all the effects of population heterogeneity and *contact* patterns that could be affecting the factors on which we focus.

The paper is organized as follows. Section 2 presents the compartmental model. Section 3 introduces the baseline parameterization and corresponding model behavior. Section 4 identifies the main evolutionary mechanisms revealed by the model and shows how they are altered by changes in transmissibility, duration, waning, and immune strength. (The Supplementary Material contains a more systematic presentation of the effects of such changes.) Section 5 compares allele-based and strain-based immunity formulations and discusses the implications for coronavirus evolution more broadly. Section 6 briefly examines how antibody-dependent enhancement (ADE) shifts model behavior and assesses the possibility it manifests characteristic population-level indicators.

## 2. Model

### 2.1 Viral genetic structure and immunity states

We consider a simplified coronavirus with two mutable genomic sites, labeled site 0 and site 1 (Figure 1). Each site has two possible alleles:

- alleles 0-0 and 0-1 at site 0 and
- alleles 1-0 and 1-1 at site 1.

**Figure 1.**
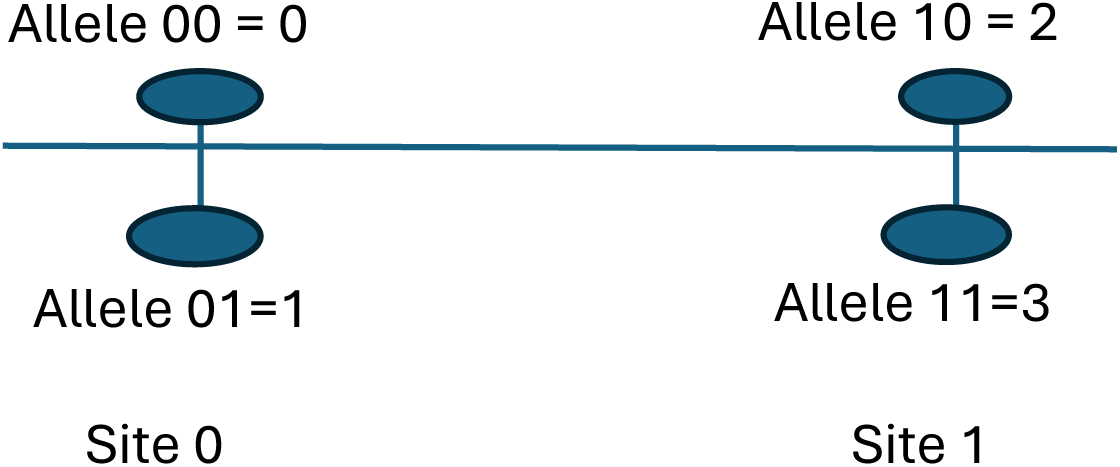
Two-sites, with two alleles per site.

We will refer to these four alleles not in the above binary notation, but by their decimal equivalents:

- alleles 0-0 and 0-1 at site 0 become alleles 0 and 1
- alleles 1-0 and 1-1 at site 1 become alleles 2 and 3.

A *strain* is defined by a choice of an allele at each site. Our model thus has four possible strains: 02, 03, 12, and 13.

Mutation occurs by replacement of one allele at one site by the alternative allele at that site. Thus, strain 02 can produce strain 12 by mutation at site 0 or produce strain 03 by mutation at site 1. Because transmission, mutation, and immune response are all represented at the allele level, the same allele can influence multiple strains.

#### Immunity

Immunity, and its complement susceptibility, plays a central role in our model. We calculate a host’s overall immunity to a strain as a function of immune exposure to that strain’s constituent alleles.

For each allele ℎ, individuals occupy one of four immunity levels:

- Level 3: no immunity; no previous infection by a strain which includes that allele.
- Level 2: first stage of “full immunity,” which arises just after recovery from an infection by a strain with that allele active.
- Level 1: second stage of full immunity to infection by that allele, but immunity whose strength is beginning to wane.
- Level 0: fully waned immunity, which may retain some level of immunity.

We track these immunity indicators as characteristics of each model compartment.

#### Waning

After infection by a strain containing allele ℎ, immunity to that allele transitions to level 2. Immunity then wanes from level 2 to level 1 at rate *w*_*h*2_, and from level 1 to level 0 at rate *w*_*h*1_.

### 2.2 Population compartments

The model contains three classes of population compartments:

1. **Never-infected susceptibles**, denoted *S*, consisting of individuals with no prior acquired immunity to any allele.
2. Recovered susceptibles, denoted

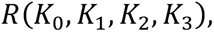

where *K*_*h*_ ∈ {0,1,2,3} is the current immunity level to allele *h*.
3. **Infectives**, denoted

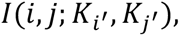

where (*i*, *j*) is the infecting strain and *i*^′^, *j*^′^ are the alternative alleles at the two sites. Since infection with strain (i, j) resets immunity to alleles *i* and *j* to level 2 upon recovery, only immunity levels to the complementary alleles *i*^′^and *j*^′^ need be tracked during infection.

For example, *R*(2,3,2,3) denotes individuals who have recently recovered from strain 02 and have never been infected with strains containing alleles 1 or 3. Likewise, *I*(0,2; 3,3) denotes individuals currently infected with strain 02 who have no prior immunity to alleles 1 or 3. Upon recovery, these individuals move into *R*(2,3,2,3).

For notational convenience, we often identify the never-infected susceptible compartment *S* as *R*(3,3,3,3).

### 2.3 Transmission, susceptibility, and mutation

#### Allele-level immunity and susceptibility

We represent immunity to allele *h* by two components:

- *m*_*h*_, the peak immune strength against strains containing allele *h*, and
- *k*_*h*_, the multiplier determined by the current immunity level *K*_*h*_.

We define

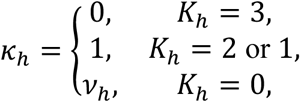

where *v*_*h*_ is the residual immunity after complete waning.

Thus, current immunity to allele *h* is represented by *m*_*h*_*k*_*h*_, and susceptibility to that allele by 1 − *m*_*h*_*k*_*h*_.

For a susceptible individual in compartment *R*(*K*_0_, *K*_1_, *K*_2_, *K*_3_), the susceptibility to infection by strain (i, j) is

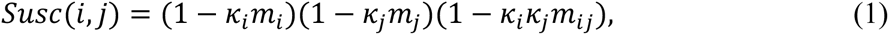

where *m*_*i*j_ represents joint immunity to the pair of alleles *i* and *j*. This term allows for epistatic or strain-level immune effects in addition to allele-level effects.

#### Transmission

We write bhi for the transmission rate for an encounter between an infected with strain (h,i) and a never-infected susceptible. In our allele-based approach we view bhi as a combination of the transmissibility bh of allele h at site 0 and the transmissibility bi of allele i at site 1, possibly modified by an epistasis factor ehi, which we ignore for the moment:

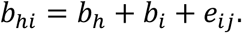

We discuss the choice of multiplicative versus additive combined transmissibility and total susceptibility in the Supplementary Material.

If an infective carrying strain (i, j) encounters a susceptible in compartment *R*(*K*_0_, *K*_1_, *K*_2_, *K*_3_), the effective transmission factor is

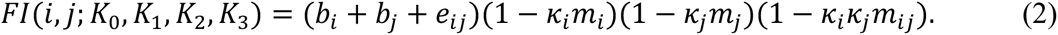

Reduction in transmissibility via multiplication by a susceptibility factor is a common practice in existing literature; for example, [Sa20, Sa24a, Sa24b, Bu25].

Let

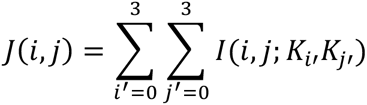

denote the total number of individuals currently infected with strain (i,j). Then, the incidence into a given infected compartment *I*G*i*, *j*; *H*_*i*_′, *H*_j_′ I in any time period Δt is proportional to

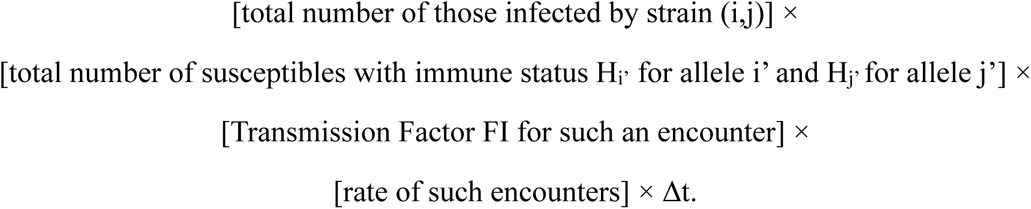

where *c* is the **effective contact rate**, the rate of possibly infection-transmitting contacts in this homogeneously mixing population.

For example, the number of additions (new infecteds) to compartment I(1,3; H0, H2) in a time period Δt is:

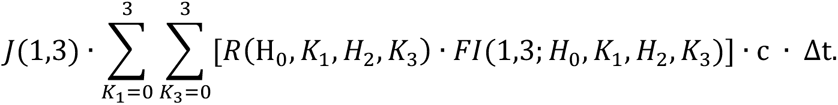

#### Mutation

With respect to our sites of interest, we assume mutation sufficiently rare as to be negligible except where amplified by within-host selection. That is, we consider mutation toward an allele only when the infectee has weak immunity to that allele (i.e., when the immunity level is 0 or 3). Thus mutation from (i, j) to (i^′^, j) depends on weak immunity to *i*^′^, and mutation from (*i*, *j*) to (*i*, *j*^′^) depends on weak immunity to *j*^′^.

The corresponding mutation probabilities are denoted *f*_*ii*_′ and *f*_jj_′. Thus, we handle mutation during transmission, modeled as a one-step change at a single genomic site, conditional on weak immunity to the destination allele. Note that this formulation also discounts as negligible the probability of simultaneous mutation at both sites of interest.

For example, the expression for Δ*I*(1,2; *H*_0_, *H*_3_), the number of new strain (1,2) infections in time interval Δt, would include mutations from strain (1,3) to strain (1,2) when immunity to allele 2 is weak, i.e., K2 ∈ {0, 3}:

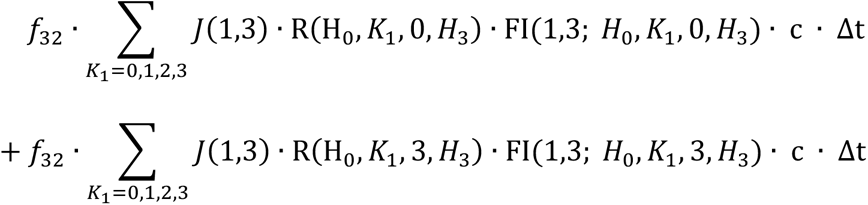

and mutations from strain (0,2) to (1,2) when immunity to allele 1 is weak, i.e., K1 ∈ {0, 3}:

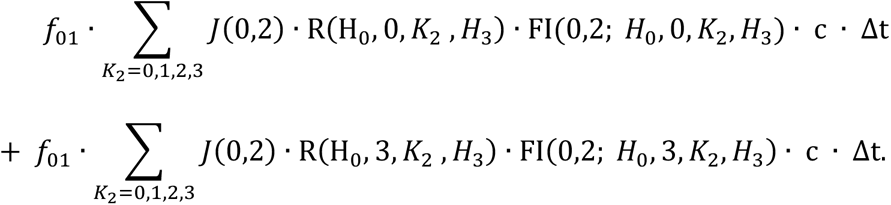

To achieve the complete dynamics for 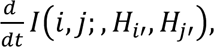 we add in expressions for

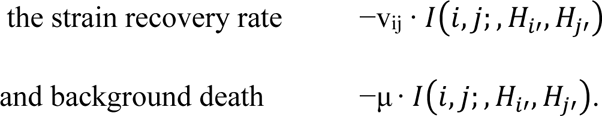

On the other hand, the complete expression for 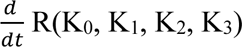 includes:

- + : Recoveries of infected;
- −: new infections via contacts with infected in the J(*i*, j)′s;
- + and − : Waning between R(K0, K1, K2, K3) levels; and
- − : Background deaths.

We present the complete set of equations in the Supplementary Material.

Our compartmental population model—with two sites, two alleles per site, and four levels of immunity—yields a system with nearly 300 differential equations. Moving to three sites would require 4,096 compartments; hence, our choice of a two-site model as a first step. One possible route to relaxing limitations in population homogeneity and site count is agent-based simulation, which we have begun exploring as a basis for future work. The system was solved numerically in Berkeley Madonna and independently verified in Python; the Python code is provided in the Supplementary Material, the Madonna code is provided in the archive [Mar20, Har20, Pau20].

### 2.4 Population susceptibility

We track the population-level susceptibility to each strain. For strain (h,j), define

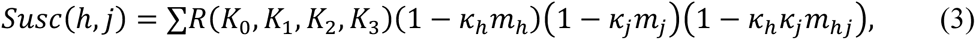

summed over all admissible recovered compartments.

The product

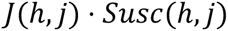

plays a role analogous to the effective reproduction number R for strain (h,j): a strain tends to generate a new epidemic wave when the number currently infected times the number susceptible to that strain, J(h,j) · Susc(h,j), increases past some critical threshold.

In the special case where *m*_*h*_ = 1 for all alleles and *v*_*h*_ = 0, population susceptibility to strain (h,j) reduces to the fraction of the population lacking immunity to both alleles *h* and *j*. More generally, however, the weighted form (3) provides a natural measure of the immune landscape experienced by each strain. See the Supplementary Material for details.

## 3. Baseline epidemic dynamics

### 3.1 Baseline parameterization

We begin by examining the behavior of the system under a baseline parameterization designed to isolate the interaction between transmission, immunity, and mutation. We present the baseline parameters in Table 1.

**TABLE 1.**
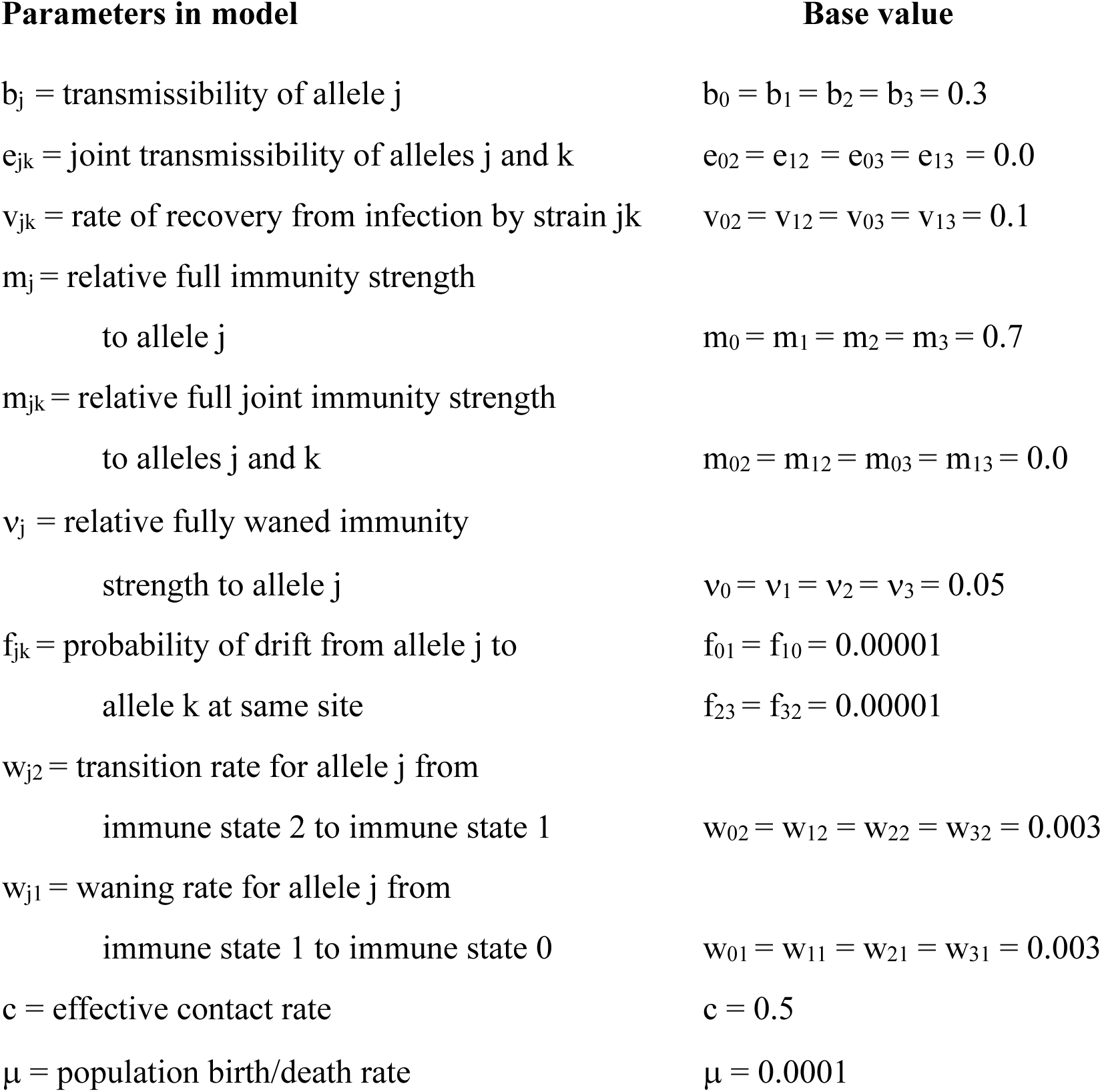
Baseline Parameter Values.

Unless otherwise stated, allele transmissibility, immune strengths, waning rates, and recovery rates are initially taken to be homogeneous across alleles and strains. These values are intended to generate illustrative epidemic dynamics rather than to fit any one coronavirus outbreak in specific.

For legibility, we will usually write the names of the R-compartments, the I-compartments, and the total strain sums without commas or parentheses. For example, we write R(2,3,2,3) as R2323, I(1,2; 3,1) as I1231, and J(0,2) simply as 02.

We normalize total population size to 1. In most simulations the epidemic is initiated by a small seed of strain 02 infections in an otherwise fully susceptible population:

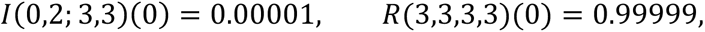

with all other compartments initially zero.

Under the baseline values *b*_0_ = *b*_1_ = *b*_2_ = *b*_3_ = 0.3, *c* = 0.5, and 𝑣_02_ = 0.1, the initial strain 02 has approximate basic reproduction number

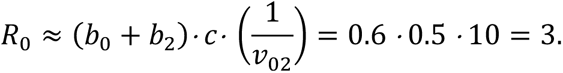

This is broadly consistent with early SARS-CoV-2 estimates, but the purpose of the baseline system is conceptual rather than empirical calibration.

### 3.2 No-mutation baseline

To understand the susceptibility dynamics most clearly, we first consider the no-mutation case by setting all mutation probabilities *f*_*i*j_ = 0. With strain 02 as the only seed and no other strain allowed to arise, the system reduces to a single-strain epidemic with waning immunity.

Figure 2 shows the prevalence of strain 02 together with the four population susceptibility curves. In the beginning, nearly everyone is susceptible ((A) in Figure 2). The initial wave of strain 02 rapidly depletes the fully susceptible compartment *R*3333, decreasing all four Susc curves (B), while generating strong immunity to alleles 0 and 2. As recovered individuals accumulate in compartments such as *R*2323, population susceptibility to strain 02 falls sharply (D). At the same time, susceptibility to the complementary strain 13 rises (C), because these recovered individuals have little or no immunity to alleles 1 and 3.

**Figure 2.**
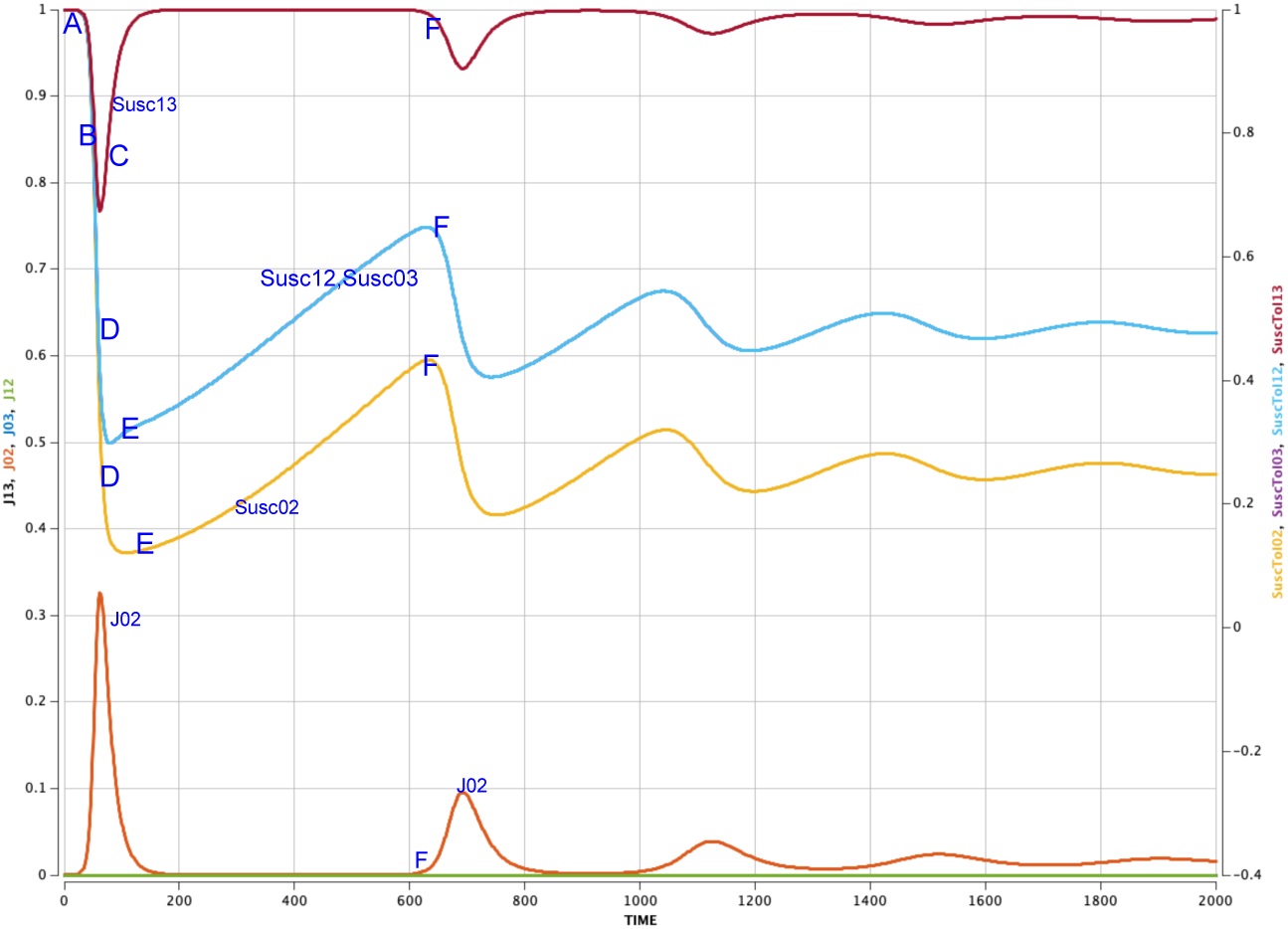
Strain 02 prevalence and the four susceptibility curves for the no-mutation scenario. Lettered annotations are discussed in main text.

Thus, even in the absence of mutation, the epidemic wave of strain 02 reshapes the immune landscape in a highly structured way. After the 02 wave subsides, births and waning gradually replenish susceptibility (E), eventually allowing a later 02 wave to occur (F). The resulting baseline pattern consists of widely separated epidemic waves with low prevalence between them.

The key point of Figure 2 is not merely recurrent infection, but the remaining elevated susceptibility to strain 13 after the 02 wave. This suggests that if mutation is reintroduced, a small number of 13 infections may be sufficient to initiate a large 13 epidemic.

The Supplementary Material contains a more detailed description of the dynamics in Figures 2 and 3.

**Figure 3.**
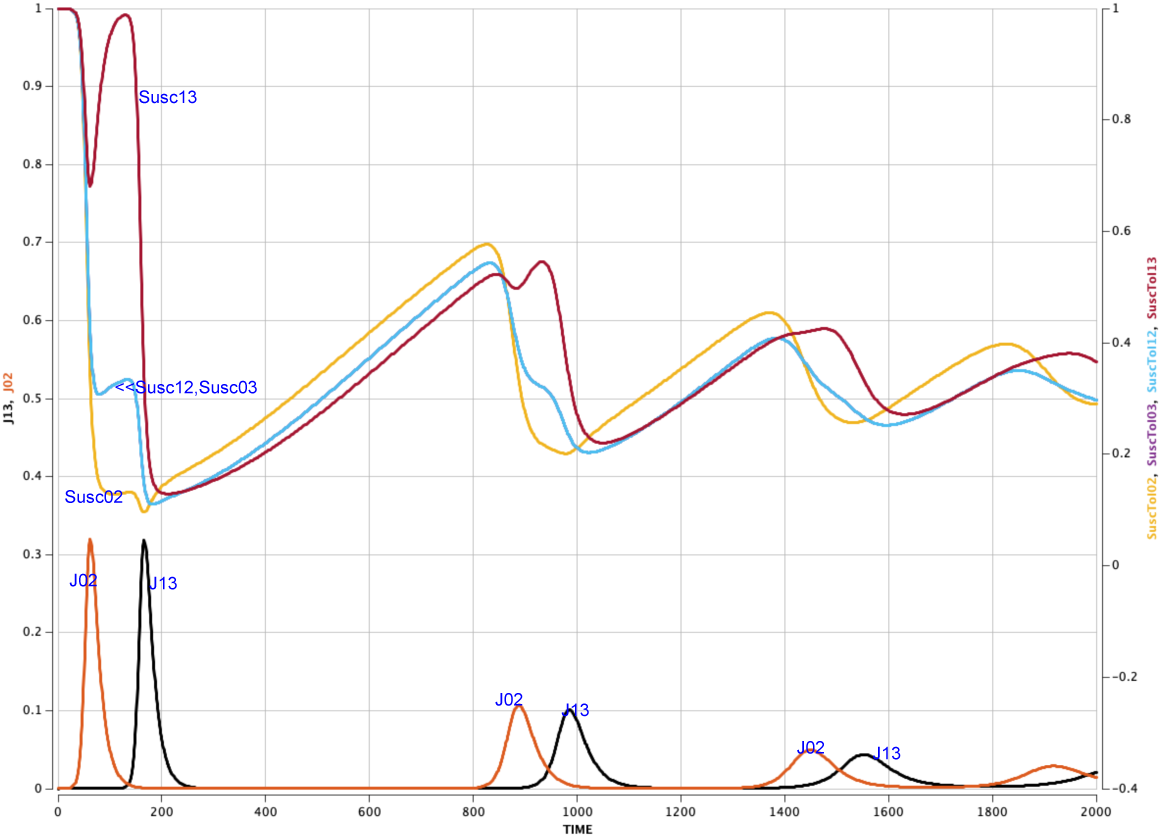
Strain prevalences and susceptibility curves for the basic parameters with mutation.

The model behind Figure 2 is basically the one-strain model that Saad-Roy et al. [Sa20, Sa21a, Sa21b, Sa22, Sa23, Sa24a, Sa24b, Es24, Wa21] use to begin their analyses, before adding vaccination and non-pharmaceutical interventions. Matching expectation, we observe a phenomenon they highlight as “accumulating immunity,” visible in declining heights of epidemic peaks and in the Susc02 curve in Figure 2.

### 3.3 Baseline with mutation

We now return to the full baseline model, reinstating small mutation rates *f*_01_ = *f*_10_ = *f*_23_ = *f*_32_ = 10^-5^. Figure 3 shows the resulting strain prevalences and susceptibility curves.

Under this baseline parameter set, the epidemic does not settle into repeated waves of the seeded strain alone. Instead, the initial wave of strain 02 generates an immune landscape that strongly favors the rise of strain 13, the strain complementary to 02. The dominant pattern therefore consists of alternating waves of strains 02 and 13, separated by intervals of low prevalence.

This behavior is not driven by mutation rate alone. It arises because recovery from strain 02 builds strong immunity to alleles 0 and 2 while leaving comparatively high susceptibility to alleles 1 and 3. By contrast, strains 03 and 12, which are each one mutational step from 02, remain weak because they still contain one allele with widespread immune history in the population.

The mechanism can be seen directly from the susceptibility terms. Immediately after a large 02 wave, much of the population satisfies approximately

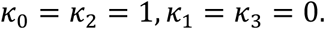

In that regime,

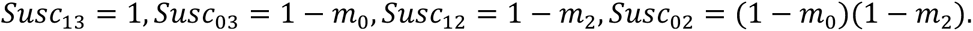

Hence,

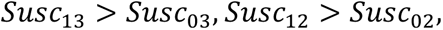

so, the complementary strain 13 encounters the weakest cross-immunity.

In effect, the first 02 epidemic wave creates the ecological conditions under which strain 13 can invade. Thus, even in this minimal two-site system, epidemic history strongly reshapes the future evolutionary trajectory.

## 4. Mechanisms shaping evolution

In the Supplementary Material, we methodically change the parameters to gain insights on how these changes affect the basic model in Figure 3. Table 2 in the Appendix summarizes those effects.

### 4.1 Four mechanisms governing strain success

These baseline dynamics motivate a more systematic examination of the mechanisms that determine which strains succeed. Across the simulations, four mechanisms consistently influence which strains prosper.

The first is **direct effects**: increased transmissibility, prolonged infection duration, weakened immunity, or accelerated immune waning for an allele tends to favor strains containing that allele.

By contrast, the other three effects are indirect in that a change in the characteristic of an allele leads primarily to changes in the prevalences of strains that do not contain that allele.

The second effect is the **complementarity effect**: after an epidemic wave of one strain, the population often becomes most susceptible to the strain with least immunological similarity. In the baseline system, this mechanism strongly favors strain 13 after a wave of strain 02.

The third is the **founder effect**: the seeded strain remains dominant even under conditions that would otherwise favor other strains.

The fourth is the **adjacency effect**: because mutation occurs one allele at a time, strains one mutational step from the current dominant strain are produced more readily than strains requiring two mutations. However, adjacency often competes with complementarity, because nearby strains also inherit greater cross-immunity from the dominant strain.

Taken together, these mechanisms show that strain success cannot be predicted from transmissibility alone. A strain may fail despite favorable transmission parameters if the surrounding immune landscape is unfavorable, while another strain may flourish only because previous epidemic waves have cleared immune space for it.

### 4.2 Direct effects

Uniform changes in transmissibility, waning, or infection duration primarily rescale the baseline dynamics. If all four transmissibilities *b*_*i*_ increase equally, if all waning rates *w*_*h*1_, *w*_*h*2_ increase equally, if all recovery rates 𝑣_*i*j_ decrease equally, or if all immunity strengths *m*_*i*_ decrease equally, the basic alternation between the founder strain and its complement is preserved, but epidemic peaks become higher and inter-wave intervals shorter.

More informative are parameter changes that affect only selected alleles or strains. Parameters with particularly strong direct effects include infection duration, waning rate, and immune strength.

Figure 4 illustrates the effect of prolonging infection for a single strain. When strain 03 is given a longer duration of infection by reducing 𝑣_03_, it becomes the dominant strain in the system. Its complement 12 also becomes more prominent, while strains 02 and 13 lose their baseline dominance. This is a clear example of direct effects overriding the founder-complement pattern of the baseline model.

**Figure 4.**
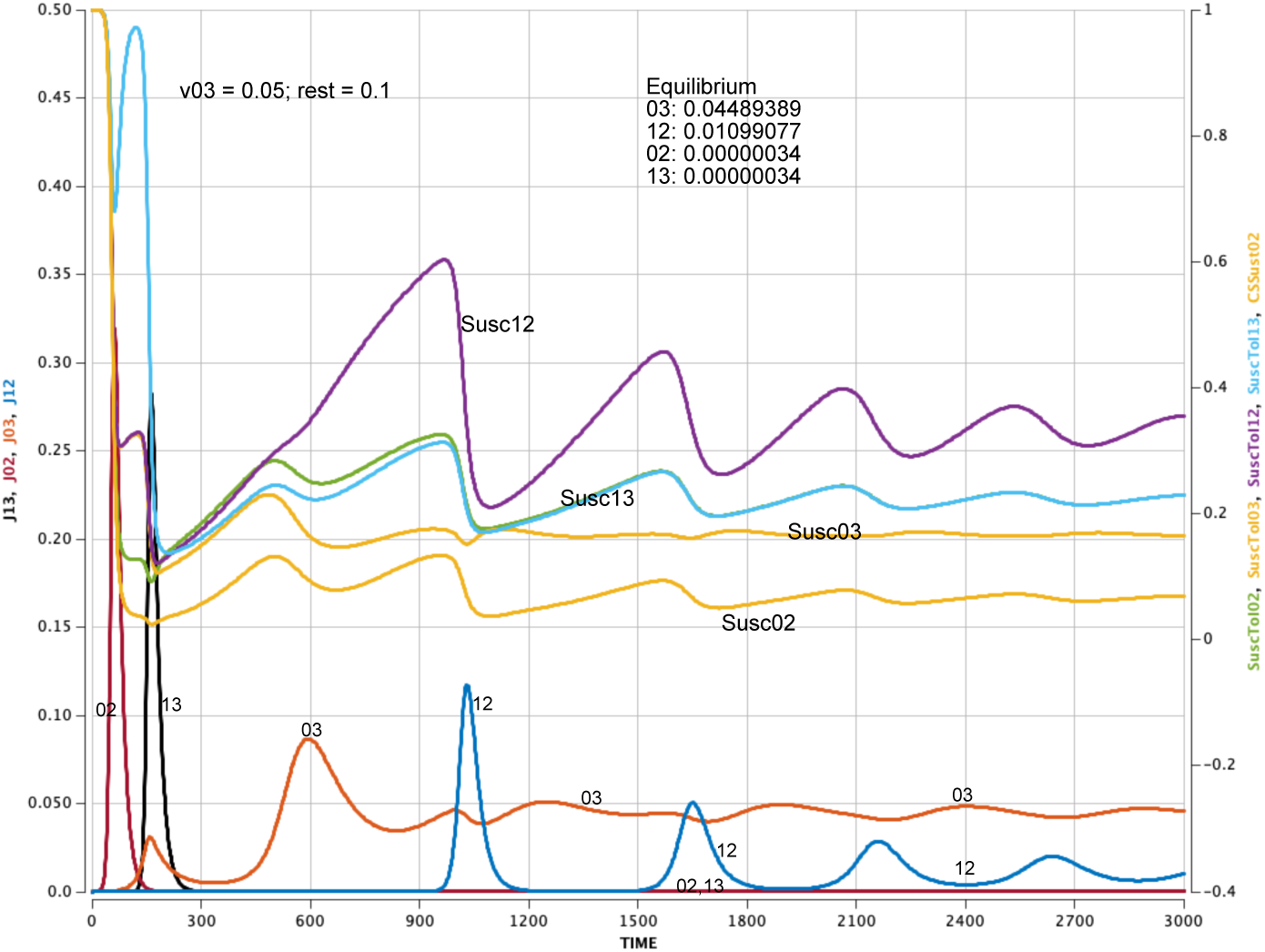
Strain 03 with the longest duration dominates the prevalences (v_03_=0.05, other v_ij_s = 0.1).

Figure 5 shows the effect of weakening immunity to a single allele. When immunity to allele 1 is reduced relative to the other alleles, the strains carrying allele 1, namely 12 and 13, become dominant. In this case the direct advantage associated with lower immune pressure to allele 1 outweighs both the founder effect favoring 02 and the complementarity effect favoring 13 after the first 02 wave.

**Figure 5.**
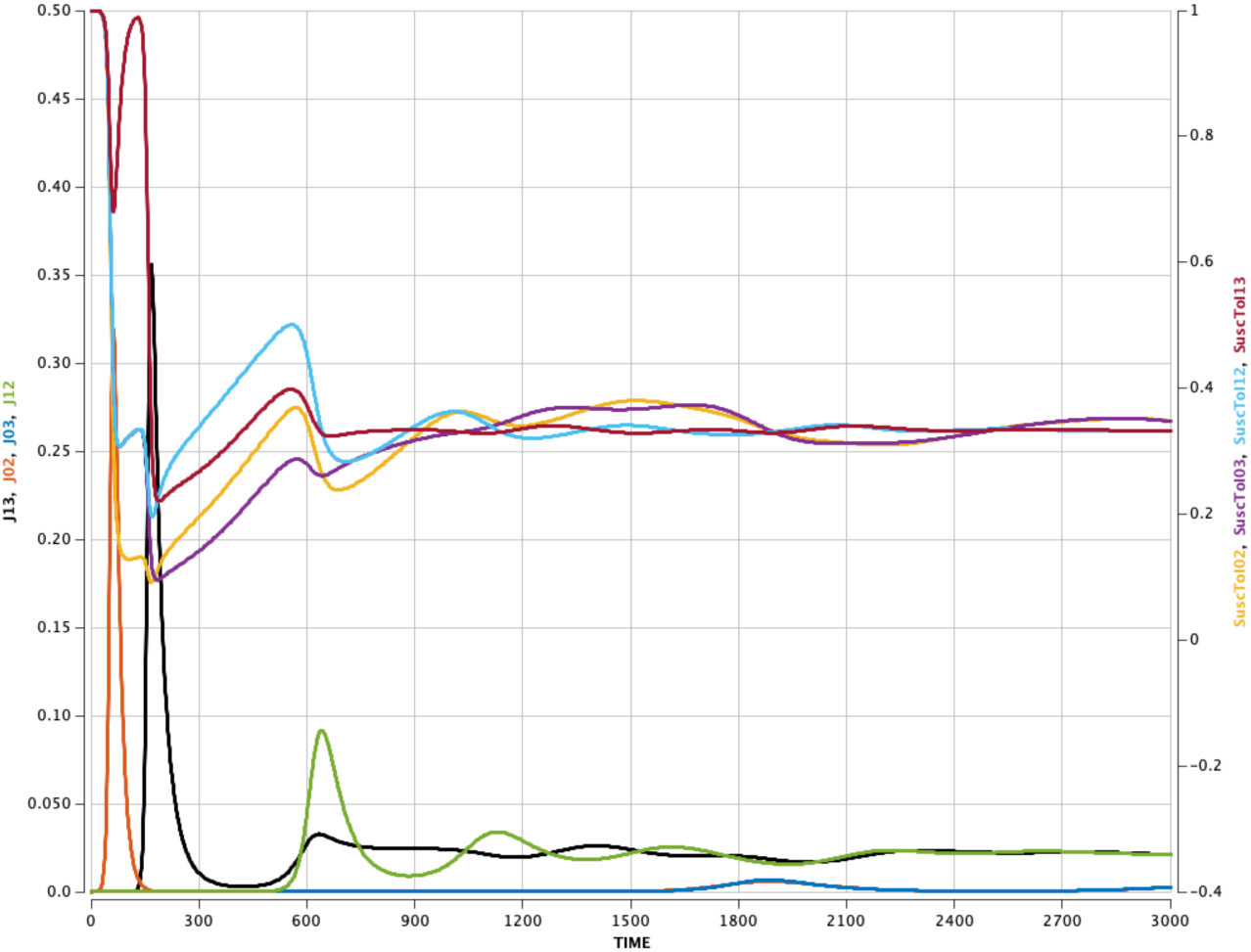
Limiting immunity to allele 1 leads to high prevalences of the strains with allele 1 (m_1_= 0.2, other m’s = 0.7).

Related simulations in the Supplementary Material show analogous behavior when waning to a particular allele is made substantially faster (Figure S7). As [Sa24a, To24] point out, waning rate makes a profound difference in the evolution of a pandemic.

In each case, strains containing the favored allele gain prevalence because they encounter systematically weaker immune suppression.

### 4.3 Complementarity effects

The baseline dynamics in Figure 3 provide the clearest illustration of complementarity: the success of a strain tends to leave greatest susceptibility to its complement. A strain and its complement share no alleles, so epidemic waves of one generate immunity patterns that often favor the other.

This mechanism persists outside the baseline regime. Even when one strain is strengthened by a direct effect, its complement can benefit indirectly through the immune landscape it creates.

Thus, complementarity is not a special property of the homogeneous baseline parameters but a recurring feature of the allele-based model.

Complementarity can also work in reverse: if a strain’s complement is made particularly weak, then the strain itself may lose one of its principal indirect advantages. Figure 6 illustrates this effect. In that simulation, alleles 0 and 2 have much higher transmissibility than alleles 1 and 3, which might be expected to favor the seeded strain 02. Instead, the weakness of 02’s complement 13 diminishes the long-run recovery of 02, and the adjacent pair 12 and 03 ultimately take over. In this case, the suppression of the complement reduces the long-run success of the founder strain itself.

**Figure 6.**
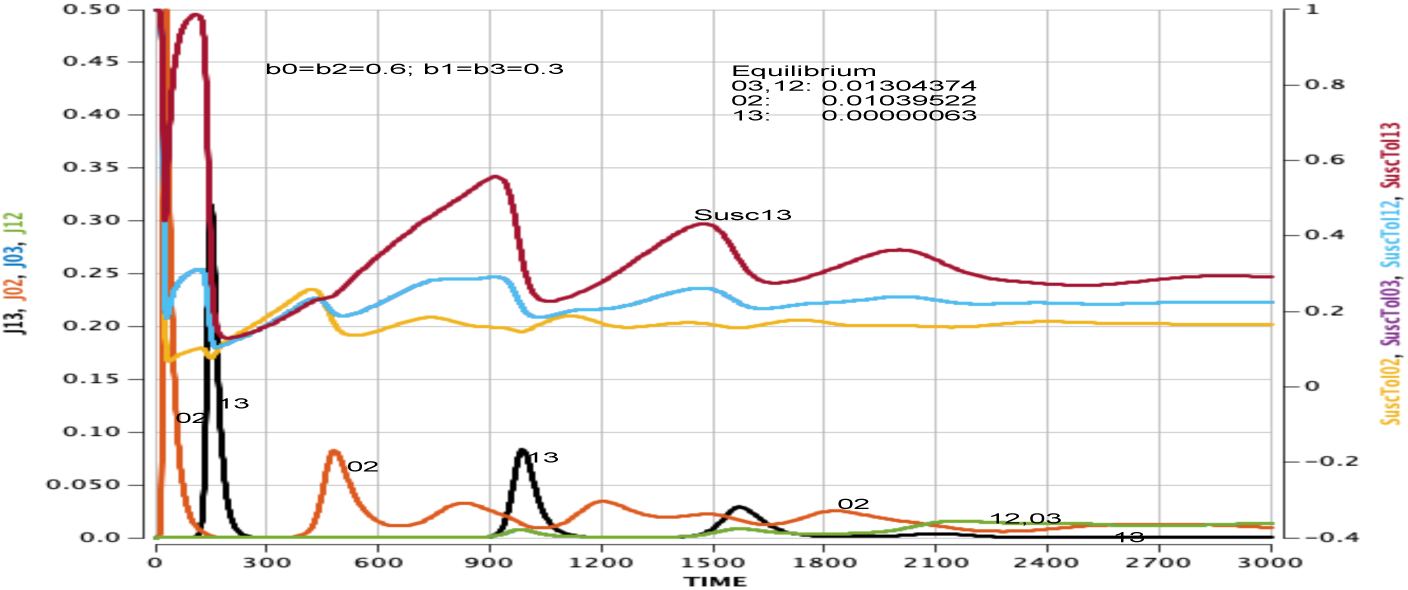
Transmissibility b_0_ = b_2_=0.6; b_1_ = b_3_ = 0.3. Doubling the transmissibility of the seeded alleles 0 and 2 diminishes the prevalence of strain 02 because its complement 13 is so weak.

These examples show that a strain’s long-term success depends not only on its own phenotypic traits, but also on those of its complement.

### 4.4 Founder effects

The founder effect describes the persistent influence of the initially seeded strain on later epidemic dynamics. In many simulations, strain 02 retains an advantage even when other strains contain alleles with more favorable transmissibility or immune parameters.

Figure 7 illustrates this clearly. There, allele 0 has lower transmissibility than alleles 1, 2, and 3, yet the seeded strain 02 remains more prominent than the stronger adjacent strain 12. The initial 02 wave imprints the immune landscape of the population in a way that continues to favor 02 and 13.

**Figure 7:**
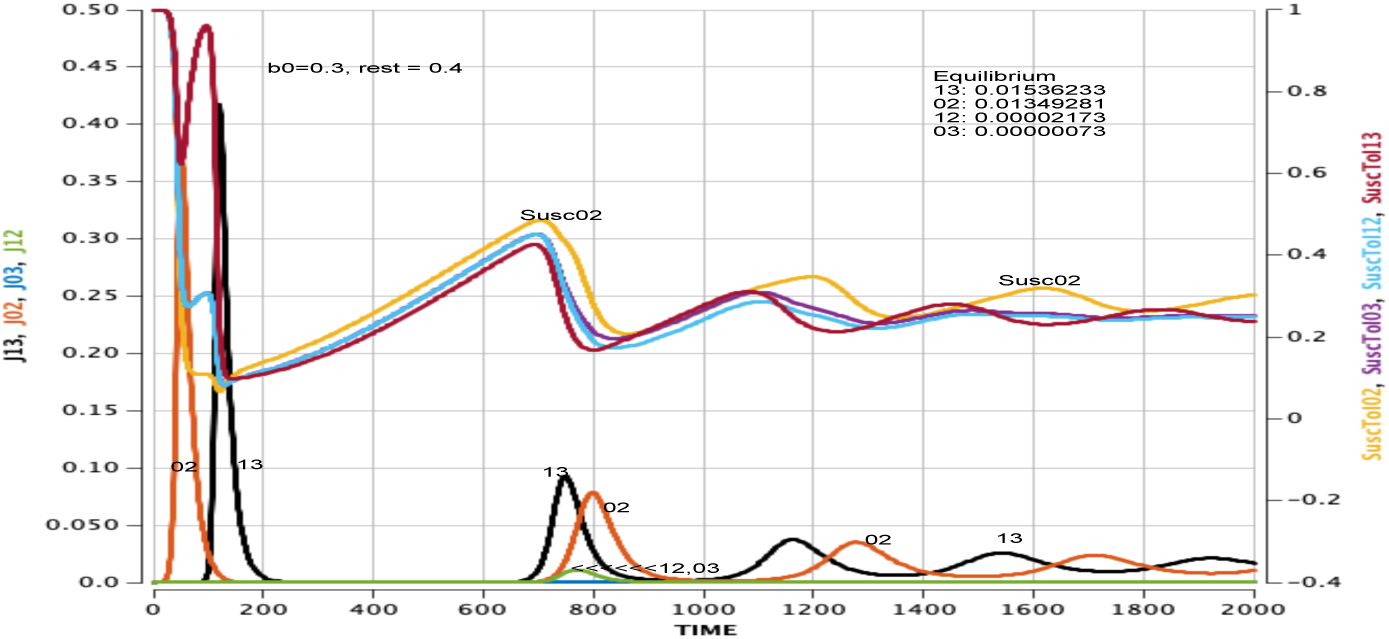
Transmissibility b_0_ = 0.3, b_1_ = b_2_ = b_3_ = 0.4. Despite a lower transmissibility for one of its alleles, seeded strain 02 maintains its presence throughout the waves of epidemics, with higher prevalence than the more contagious strain 12.

Figure 8 shows a related example involving immunity. When immunity to allele 0 is reduced modestly, strains 02 and 03 should both gain a direct benefit. Nevertheless, the long-run pattern remains dominated by seeded strain 02 and its complement 13. The founder effect therefore persists even when the altered parameter should have favored another strain sharing the weakened-immune allele.

**Figure 8.**
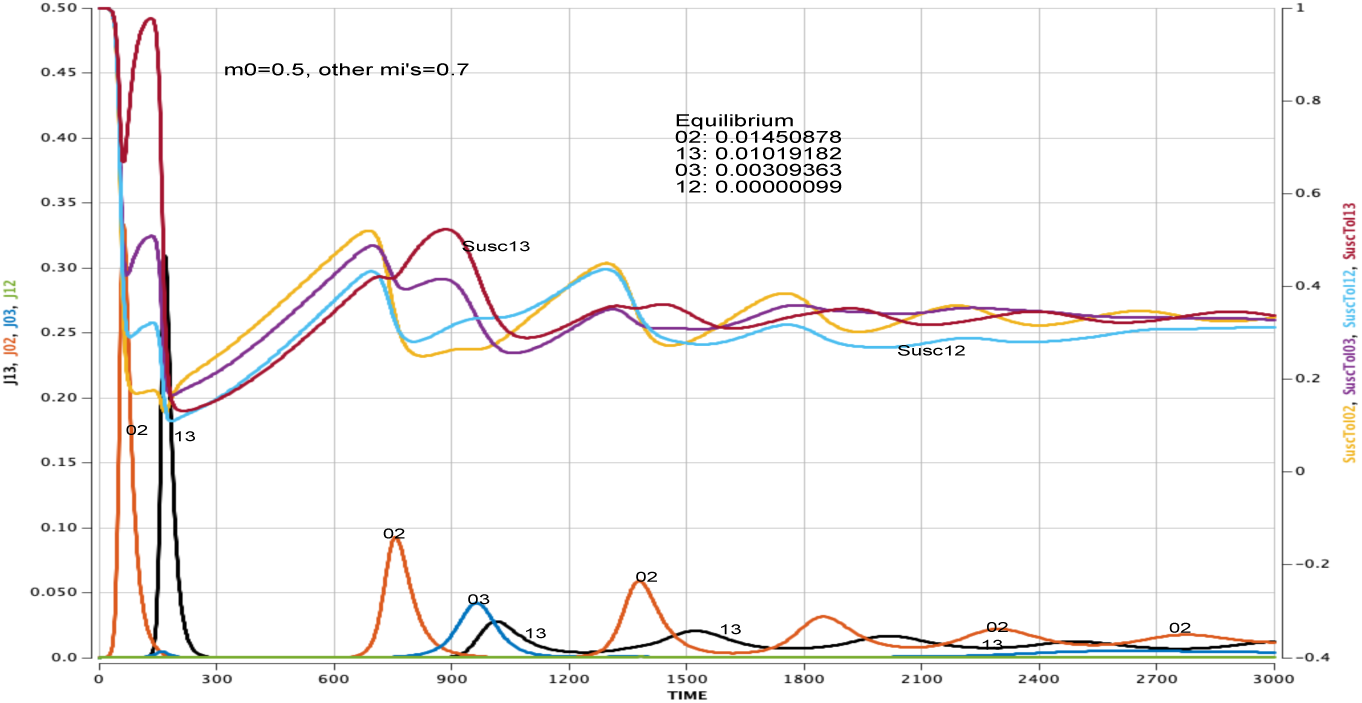
Lowering immunity m_0_ to allele 0 briefly favors strains 02 and 03, but seeded strain 02 and its complement 13 eventually dominate (m_0_ = 0.5, other m_i_’s = 0.7).

A particularly revealing case occurs when immune responses are weak for all alleles. Figure 9 shows that when all *m*_*i*_ are small, all four strains become active and the differences among susceptibility curves narrow. Even in this regime, however, the prevalences of the seeded strain 02 and its complement 13 are three times the prevalences of the other strains. Thus, founder effects do not disappear when immunity weakens.

**Figure 9.**
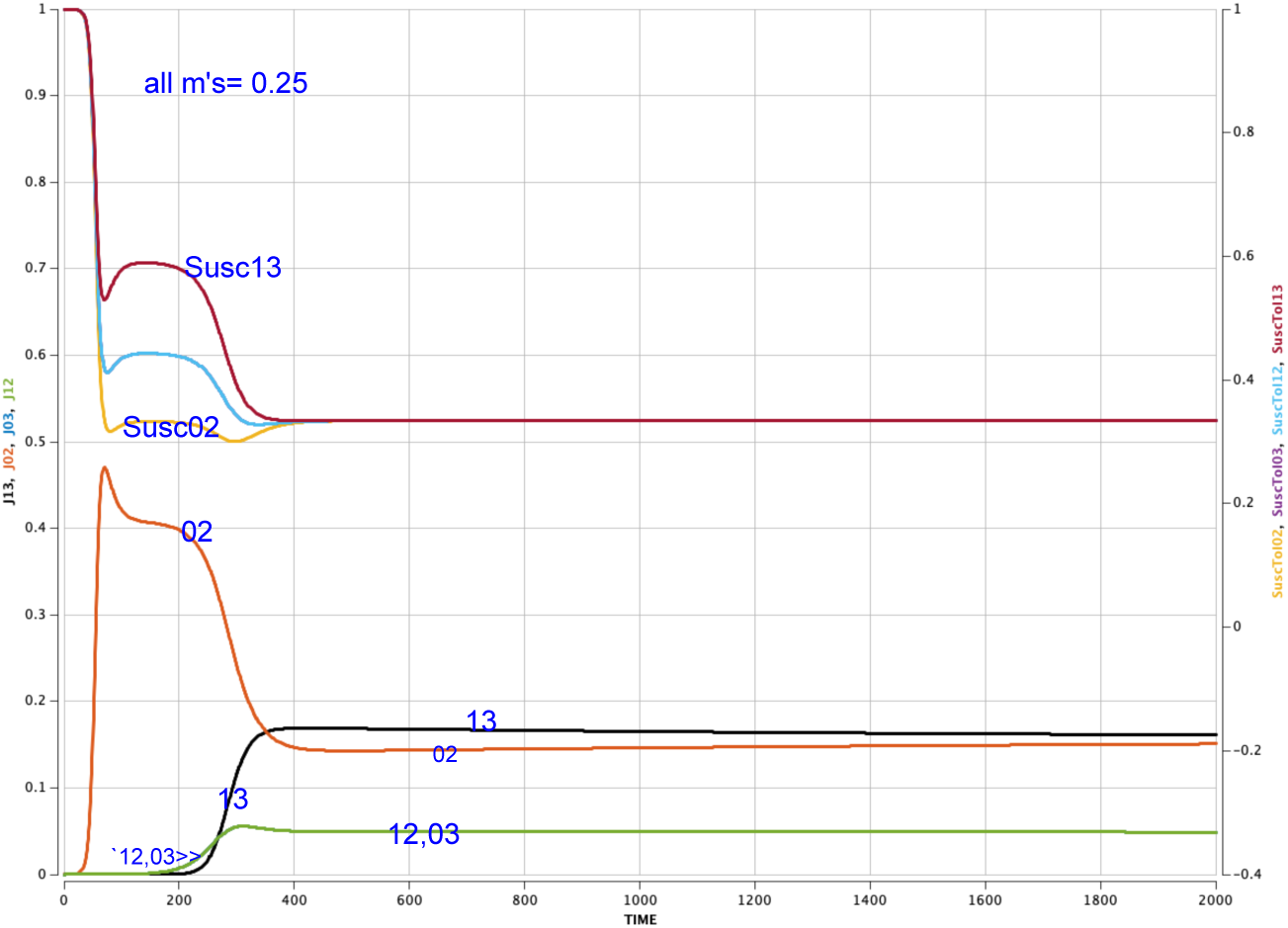
When all allele immunities are low (mi’s = 0.25), infection waves disappear; all four strains appear and stabilize quickly, yet the seed and its complement still dominate.

### 4.5 An Example with All Four Mechanisms

To illustrate the robustness of the Complementarity and Founder Effects, we facilitated the emergence of a single-allele mutation of seeded strain 02 by multiplying the probability of a 0→1 mutation by a factor of 10,000 in the baseline model (in order to drive a quick second peak of strain 12). Figure 10 presents the resulting dynamic. Via the Direct Effect and the Adjacency Effect, strain 12 has a strong early epidemic peak, dominating the simultaneous peak of seeded strain 02. Its complement, strain 03, dominates the follow-up wave. However, strain 13 is now bolstered by its role as complement of 02 and the high 0→1 mutation probability. Strains 02 and 13, with their reenforcing high susceptibilities, once again dominate the pandemic after the first inter-wave pause.

**Figure 10.**
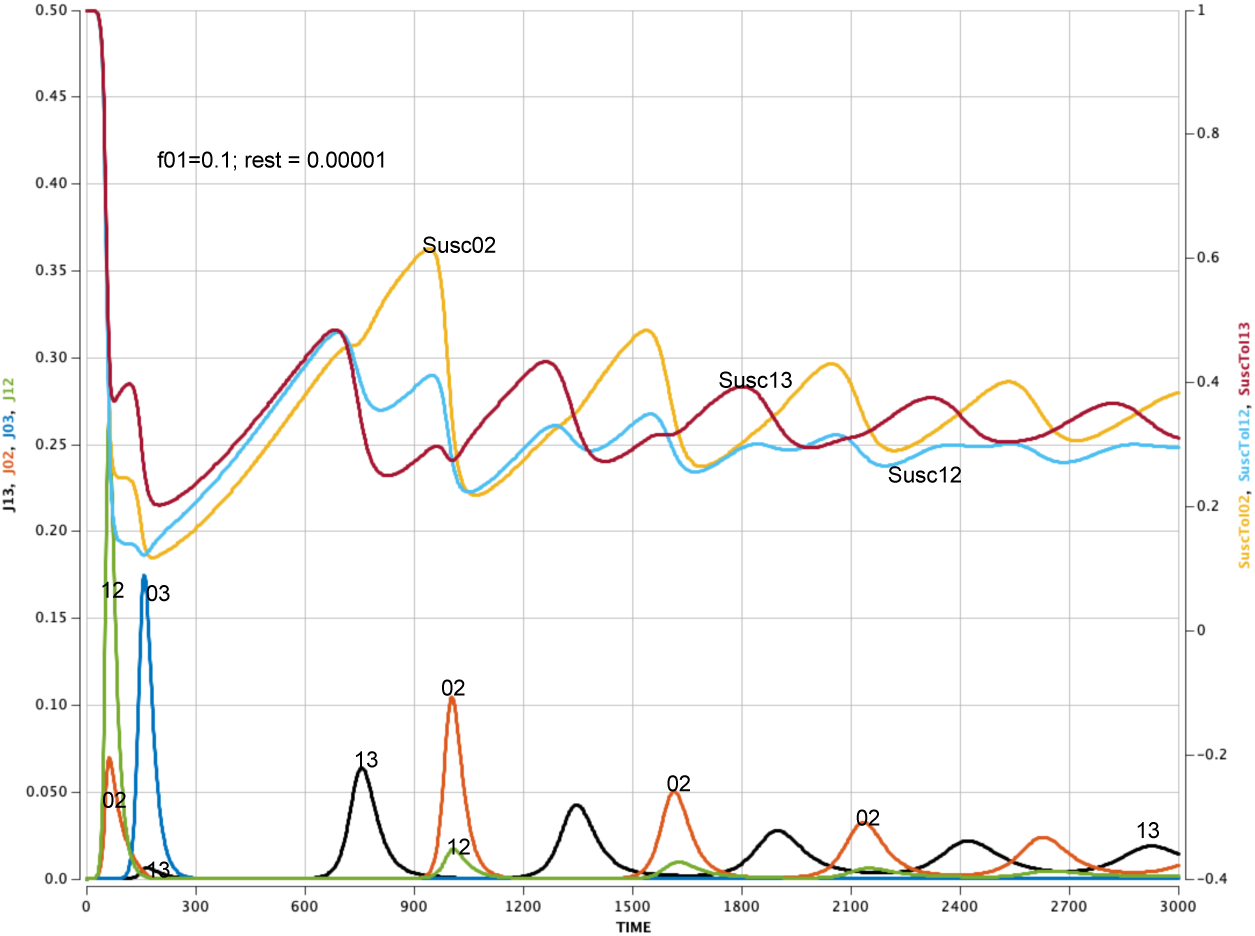
Baseline model with mutation rate f_01_ = 0.1; all other mutation rates = 0.00001.

### 4.6 Immune pressure and the direction of evolution

Across the model, immunity is the main organizing force determining whether evolution proceeds toward nearby mutants or toward complementary strains. Strong allele-level immunity suppresses strains that share alleles with the current dominant strain, creating opportunities for complementary strains. Weak immunity diminishes this suppression, making mutational adjacency more important and allowing nearby strains to establish themselves more easily.

Figure 5 provides an example in which a direct immune advantage dominates both founder and complementarity effects: weakening immunity to allele 1 favors strains 12 and 13. By contrast, Figure 8 shows that when immunity to allele 0 is weakened only moderately, the founder strain 02 and its complement 13 still dominate. Figure 9 shows that when immunity is weak for all alleles, all four strains become active and stable.

At the extreme, when immune pressure against the seeded alleles is absent, mutation may effectively disappear from the epidemic trajectory. Simulations in the Supplement with *m*_0_ = *m*_2_ = 0 show that only strain 02 remains present (Figure S17). In the language of Saad-Roy and colleagues [Sa22], when there is no immune pressure, viral abundance may remain high but selection for immune escape is absent.

These results reinforce the central message of the paper: immunity does not merely reduce transmission. It also shapes the direction of viral evolution by introducing ecological interactions among strains that dynamically reshape the viral fitness landscape.

## 5. Allele-based versus strain-based immunity

### 5.1 Joint immunity and its interaction with allele-level immunity

We now examine how introducing joint immunity terms *m*_*i*j_ alters the baseline dynamics. We find that the effect of these terms depends critically on the strength of allele-specific immunity *m*_*i*_.

When allele-level immunity is strong (e.g., *m*_*i*_ ≳ 0.5), adding moderate joint immunity terms has little qualitative effect on the dynamics. The baseline pattern of alternating waves of the founder strain and its complement remains largely unchanged. In this regime, immune pressure is already strong at the allele level, and additional joint effects do not substantially alter which strains encounter the least cross-immunity.

In contrast, when allele-level immunity is weak (e.g., *m*_*i*_ ≈ 0.3), joint immunity can significantly reshape evolutionary outcomes. Figure 11 illustrates this effect. As the joint immunity parameters *m*_*i*j_ increase:

- the initial waves of strains 02 and 13 become less pronounced;
- strains 12 and 03—each one mutation away from the seeded strain—gain prevalence; and
- for sufficiently large *m*_*i*j_, the adjacent strains 12 and 03 dominate the long-run dynamics.

**Figure 11.**
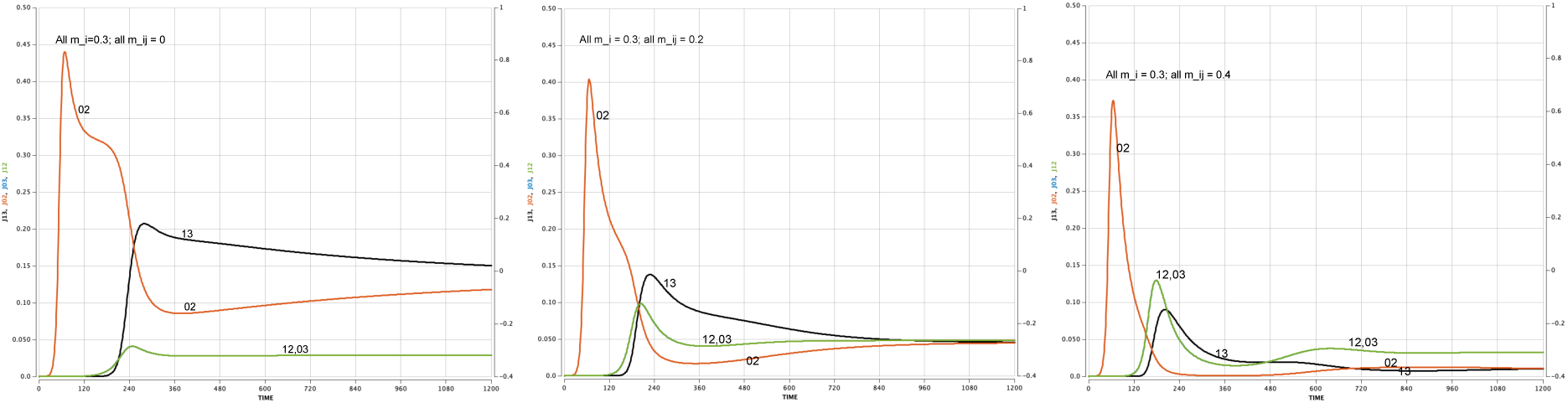
For low values of immunity response m_i_ to all alleles i, the advantages of mutant proximity outweigh the advantages of complementarity as the joint m_ij_’s increase (all m_i_’s=0.3 with all m_ij_’s = 0 (left), all m_ij_’s = 0.2 (center), all m_ij_’s = 0.4 (right)).

In this regime, joint immunity effectively suppresses the complementarity advantage of strain 13 by reducing susceptibility differences across strains. As a result, mutation adjacency becomes the dominant mechanism, and the system favors nearby strains over complementary ones.

These results highlight that the **structure of immunity**—whether it operates primarily at the allele level or at the strain level—can qualitatively change evolutionary trajectories.

### 5.2 Direct comparison with strain-based immunity

To clarify the distinction between allele-based and strain-based formulations, we compare two parameter regimes that produce similar initial epidemic waves but differ in how immunity is structured.

In the first case (allele-based immunity), we set *m*_*i*_ = 0.5 for all alleles and *m*_*i*j_ = 0. Immunity is therefore applied independently at each allele.

In the second case (strain-based immunity), we set *m*_*i*_ = 0 and assign joint immunity *m*_*i*j_ = 0.75 for all strain pairs. This concentrates immunity at the level of whole strains rather than individual alleles.

The two cases are calibrated so that the initial wave of strain 02 is identical. For individuals in compartment *R*2323, the effective susceptibility to reinfection by strain 02 is the same in both cases:

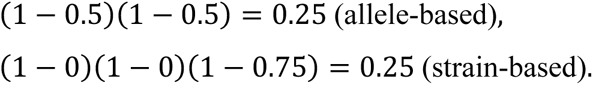

Despite this identical initial condition, the subsequent dynamics diverge sharply (12).

In the allele-based case, recovery from strain 02 generates independent immunity to alleles 0 and 2. This suppresses the adjacent strains 03 and 12, each of which shares one allele with 02, and instead favors the complementary strain 13, which shares neither allele. As a result, the system evolves toward strain 13, and the epidemic alternates between 02 and 13.

In the strain-based case, immunity is not decomposed across alleles. Individuals recovered from 02 do not experience partial protection against strains 03 and 12. These adjacent strains therefore encounter little immune resistance and can expand rapidly. As a result, the epidemic shifts toward strains 03 and 12 rather than toward the complementary strain 13.

This comparison highlights a fundamental distinction:

- In **allele-based immunity**, immune pressure acts on shared genetic components, favoring strains that are maximally distinct in allele space.
- In **strain-based immunity**, immune pressure acts on whole strains, allowing nearby mutants to exploit weak cross-immunity.

### 5.3 Immune structure and evolutionary pathways

The structure of immunity determines whether viral evolution proceeds locally or through larger effective jumps in strain space.

When immunity is primarily allele-based, recovery from infection produces selective pressure against strains sharing individual alleles with the dominant strain. This suppresses nearby mutants and favors strains that differ at multiple sites. As a result, evolution tends to proceed toward complementary strains, generating effective long-range transitions in strain space.

When immunity is primarily strain-based, this selective pressure is weakened. Nearby mutants retain high transmissibility because they are not penalized at the level of individual alleles. In this case, mutation adjacency dominates, and evolution proceeds through incremental steps.

This distinction is biologically important. It suggests that the way host immunity is distributed across viral features—whether it targets specific genomic components or whole antigenic profiles—can determine whether viral evolution follows gradual local exploration or more abrupt shifts toward antigenically distant strains.

## 6. Antibody-dependent enhancement (ADE) as an extension

We briefly examine how the inclusion of antibody-dependent enhancement (ADE) affects the dynamics of the model, as a further illustration of the potential utility of an allele-based approach.

ADE occurs when slightly mismatched antibodies left over from prior infection facilitate viral entry into host cells, thereby increasing susceptibility. This mechanism has been well documented in dengue [Fe99] and has been proposed as a possible, though uncertain, feature of coronavirus infections [Te20, Ik23, Song23, Yip14].

To incorporate ADE, we assign each allele *i* a value *d*_*i*_ representing the potential increase in susceptibility due to prior exposure to that allele. These contributions are added to the transmission term, but only for individuals who have previously been infected with strains containing the relevant allele.

Thus, ADE increases transmission in a state-dependent way: it applies only to individuals with prior immune exposure, and not to naïve susceptibles.

Figure 13 illustrates the effect of increasing ADE parameters under otherwise baseline conditions. The initial wave of the seeded strain 02 is largely unaffected, since most individuals are immunologically naïve at that stage. However, subsequent waves—particularly those of the complementary strain 13—are amplified.

**Figure 12.**
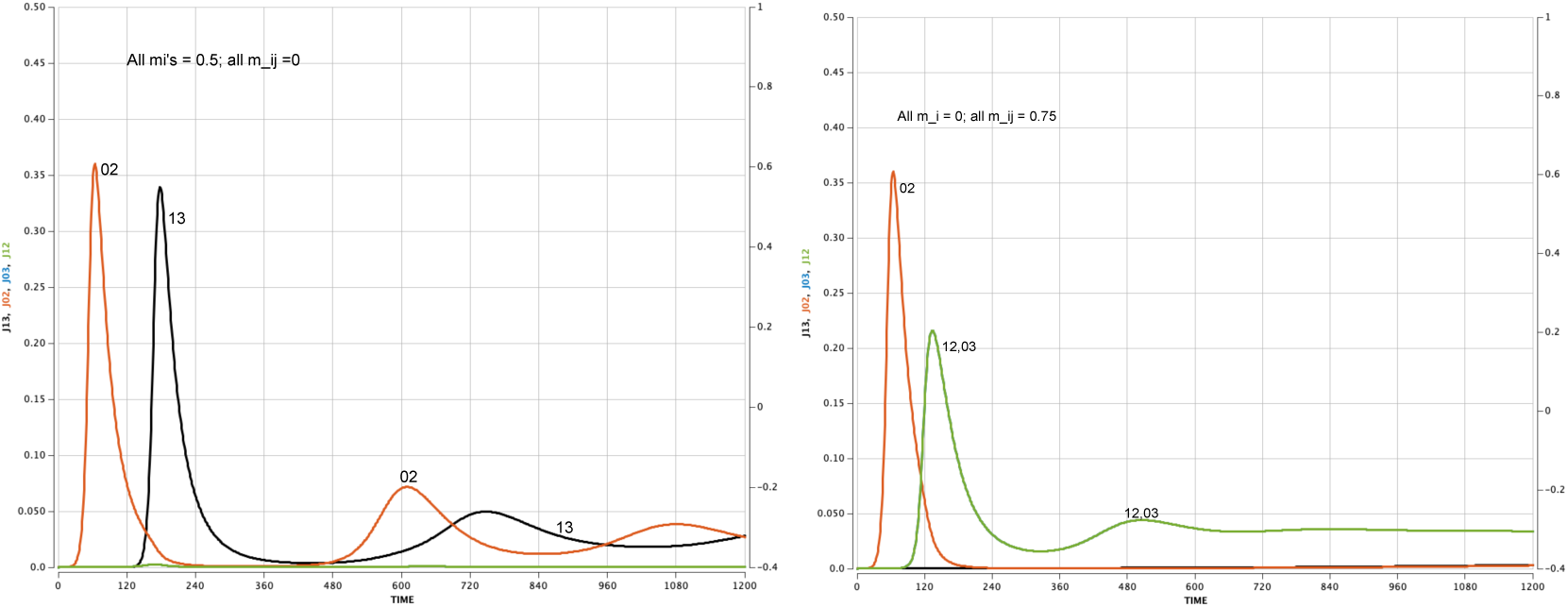
Case 1 (left). All m_i_’s = 0.5, all m_ij_’s =0. With independent allele-based immunities, the recovery-induced immunities for alleles 0 and 2 prevent strains 12 and 03 from gaining a foothold before complementary strain 13 arrives and takes off. Case 2 (right). All m_i_’s = 0, all m_ij_’s = 0.75. In the strain-based case 2, early-forming strains 12 and 03 take off and dominate the later forming 13 strain, and even the seeded 02 strain.

**Figure 13.**
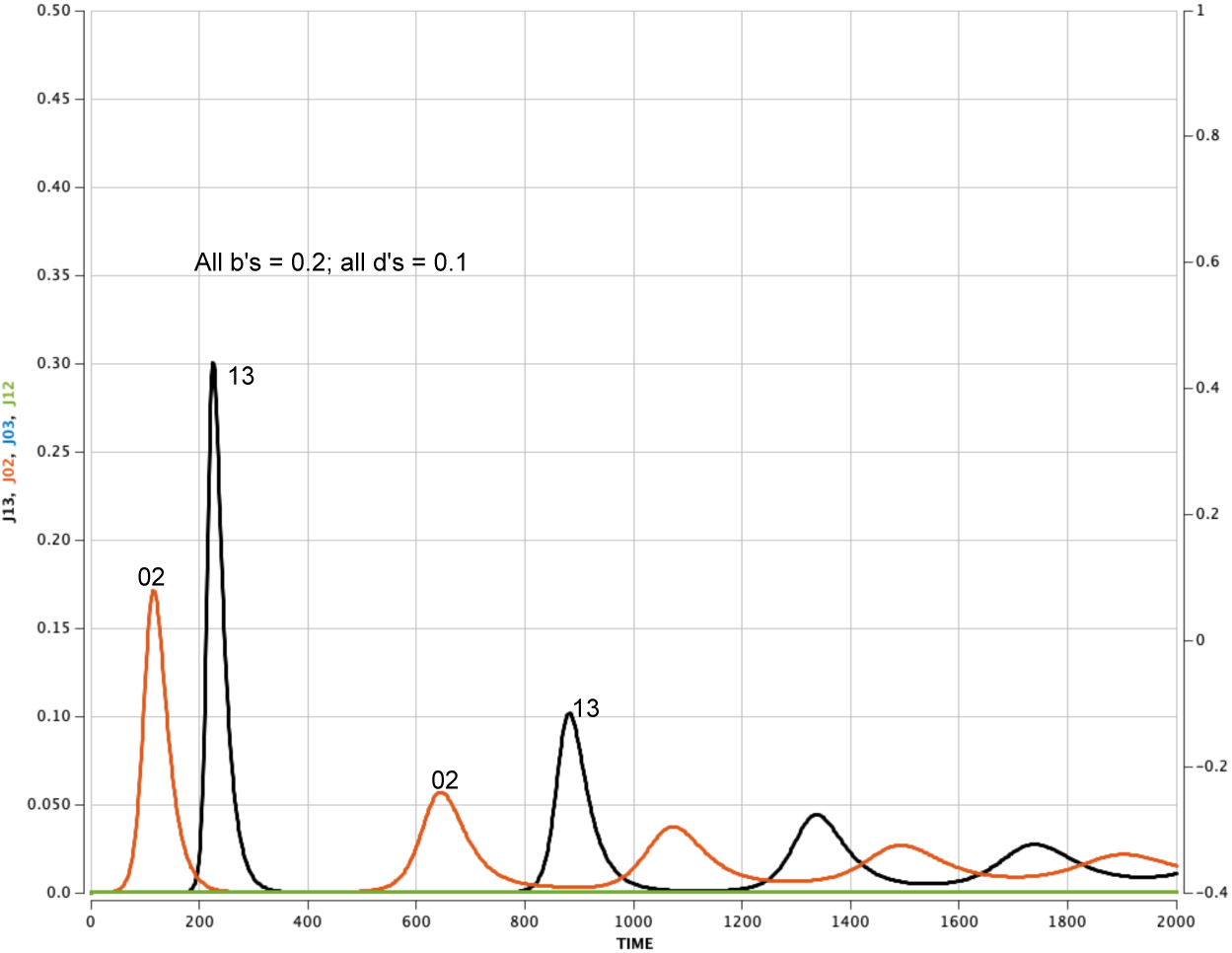
Strain 13 has especially high peaks if an ADE factor (d_i_ = 0.1) is added to the basic model.

As ADE increases:

- peaks of later epidemic waves become higher,
- inter-wave intervals shorten, and
- endemic prevalence between waves increases.

In some cases, peak prevalence of the complementary strain exceeds that of the original founder strain. This reversal does not occur in the baseline model without ADE and suggests that ADE can amplify the complementarity mechanism by increasing transmission precisely in those populations shaped by prior infection.

Although the model is stylized, this result raises the possibility that analysis of population-level epidemic patterns could, in principle, help screen for the possibility of ADE-like mechanisms when combined with more realistic biological assumptions.

## 7. Discussion

This paper develops a novel framework for studying viral evolution under population immunity in which transmission, mutation, and immune response are represented at the level of individual genomic components. Even in its simplest form, the model reveals qualitative features of epidemic evolution that are not apparent in standard strain-based approaches.

The central result is that **the structure of immunity strongly shapes evolutionary trajectories**. In the baseline system, an epidemic wave generated by a given strain does not primarily favor nearby mutants. Instead, it often creates the highest susceptibility to a genetically complementary strain—one that shares the fewest immunologically relevant alleles with the dominant predecessor. This complementarity effect produces alternating epidemic waves and illustrates how epidemic history reshapes the effective fitness landscape.

A second result is the persistence of **founder effects**. The identity of the initially circulating strain can influence long-run dynamics even when competing strains have more favorable transmission or immune parameters. This persistence arises because immune history is organized around the alleles of the founder strain, and because its complementary strain benefits from the same immune structure. In this sense, early epidemic conditions can leave a lasting imprint on evolutionary outcomes.

A third result is that **mutation adjacency alone does not determine evolutionary turnover**. Although mutations occur one allele at a time, nearby strains are also those most exposed to residual cross-immunity. When immune pressure is strong, it suppresses adjacent mutants and instead favors more distant strains. When immune pressure is weak, adjacency becomes dominant and evolution proceeds locally. Mutations in the contemporary COVID-19 pandemic seem more like the former case than the latter. The transition to the Delta strain has nine mutations in the spike protein itself and thirteen mutations in the added regions. The transition from Delta variant to Omicron has more than 50 known mutations, 32 of which are in the spike protein [Ch23, Ing21]. Furthermore, pre-Omicron, antigenic cartography analysis suggests a structured immune landscape, with certain spike protein substitutions exhibiting consistent additive effects across diverse genetic backgrounds [Wi26] — broadly consistent with the “mix-and-match” paradigm of allele-based evolution explored in this work.

Taken together, these results highlight a key distinction between allele-based and strain-based formulations. In strain-based models [K-S21, Getz21], mutation pathways are typically imposed through a predefined network or transition matrix, with cross-immunity defined separately. In the present framework, mutation arises directly from changes at genomic sites, and immune protection can be assigned either to individual alleles or to combinations of alleles. This makes it possible to examine how the internal structure of viral genetics interacts with population immunity to shape evolutionary dynamics. Comparison of these modeling choices shows that accounting for cross-immunity structure between alleles can produce qualitatively different epidemic trajectories, even when their initial behavior is similar.

Although the model is stylized, its mechanisms are biologically motivated. Coronaviruses evolve through mutations at multiple genomic sites, and both immune escape and transmissibility are often associated with specific regions of the genome. The model demonstrates how such site-specific effects can scale up to population-level patterns. In particular, it provides a conceptual explanation for why evolution may sometimes proceed through large effective antigenic shifts rather than through incremental local changes.

The framework is not intended as a predictive tool. It does not incorporate realistic contact structure [So23, El20, Oz21, Kerr21], host heterogeneity [Oz21, Kerr21], vaccination programs or non-pharmaceutical interventions [Dys21, Gi20, Mo21, Sa20, Sa21a, Sa23, Sa24a, Sa21b], or the full genomic complexity of coronaviruses [Ger26, Zvy23]. Instead, it isolates a small number of interacting processes—mutation, waning immunity, and allele-specific transmission—in order to identify general mechanisms. These mechanisms are likely to persist in more detailed models and may help interpret patterns observed in real epidemics.

Several extensions follow naturally. Increasing the number of mutable sites would allow richer genetic structure, more realistic epistatic interactions, and exploration of how complementarity generalizes when antigenic antipodes are diffuse and overwhelmingly distant. Incorporating heterogeneous populations and vaccination would allow the immune landscape to vary across subgroups. Linking allele-specific parameters more directly to experimental virology would provide a bridge between laboratory measurements and population dynamics [Xu15, Top24]. These directions would extend the present framework while preserving its central insight.

More broadly, the results suggest that **population immunity acts not only as a constraint on transmission but also as a driver of evolutionary change**. By shaping which genetic combinations encounter the least resistance, the immune landscape can redirect viral evolution in systematic ways. Capturing this interaction between genetic structure and population immunity is essential for understanding the long-term behavior of evolving pathogens.

## Data Availability

All data produced in the present work are contained in the manuscript.

## APPENDIX

**Table 2.**
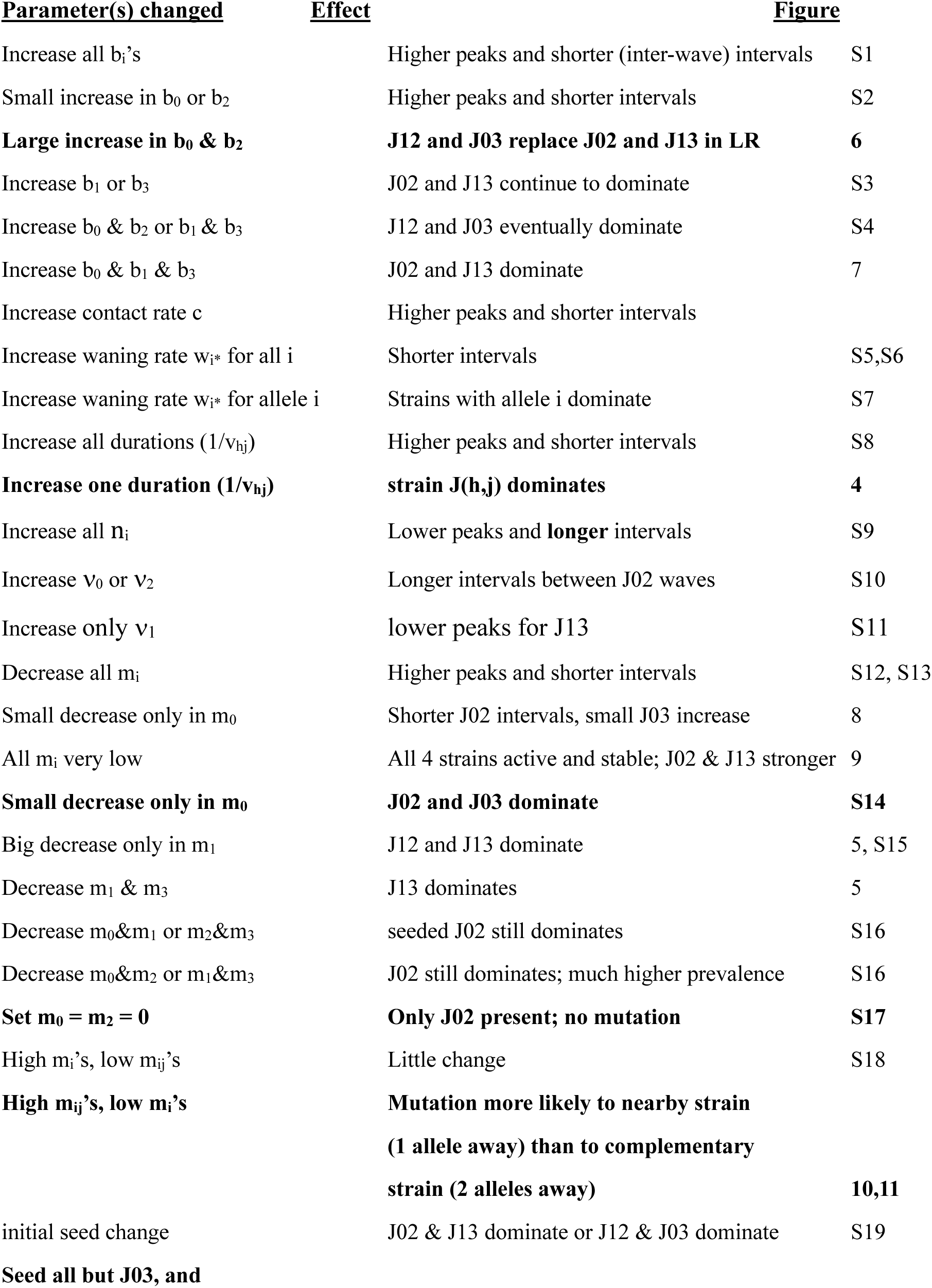
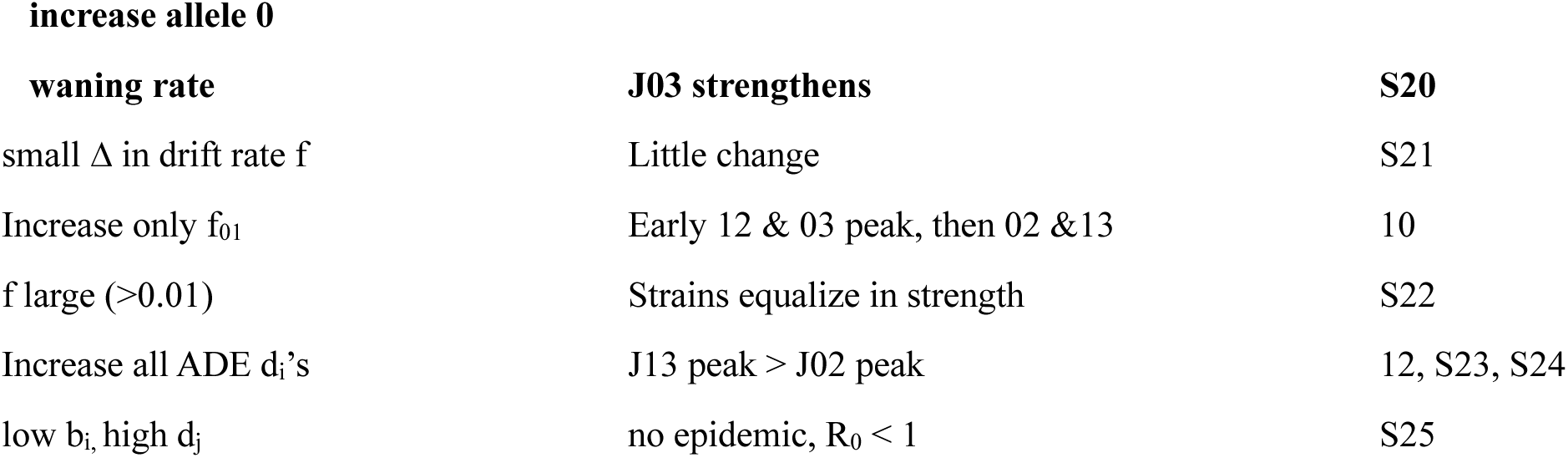
SUMMARY OF EFFECTS OF PARAMETER CHANGES.

| <u>Parameter(s) changed</u> | <u>Effect</u> | <u>Figure</u> |
| --- | --- | --- |
| Increase all $b_i$'s | Higher peaks and shorter (inter-wave) intervals | S1 |
| Small increase in $b_0$ or $b_2$ | Higher peaks and shorter intervals | S2 |
| **Large increase in $b_0$ & $b_2$** | **J12 and J03 replace J02 and J13 in LR** | **6** |
| Increase $b_1$ or $b_3$ | J02 and J13 continue to dominate | S3 |
| Increase $b_0$ & $b_2$ or $b_1$ & $b_3$ | J12 and J03 eventually dominate | S4 |
| Increase $b_0$ & $b_1$ & $b_3$ | J02 and J13 dominate | 7 |
| Increase contact rate $c$ | Higher peaks and shorter intervals | |
| Increase waning rate $w_i^*$ for all $i$ | Shorter intervals | S5,S6 |
| Increase waning rate $w_i^*$ for allele $i$ | Strains with allele $i$ dominate | S7 |
| Increase all durations ($1/v_{hj}$) | Higher peaks and shorter intervals | S8 |
| **Increase one duration ($1/v_{hj}$)** | **strain J(h,j) dominates** | **4** |
| Increase all $n_i$ | Lower peaks and **longer** intervals | S9 |
| Increase $v_0$ or $v_2$ | Longer intervals between J02 waves | S10 |
| Increase only $v_1$ | lower peaks for J13 | S11 |
| Decrease all $m_i$ | Higher peaks and shorter intervals | S12, S13 |
| Small decrease only in $m_0$ | Shorter J02 intervals, small J03 increase | 8 |
| All $m_i$ very low | All 4 strains active and stable; J02 & J13 stronger | 9 |
| **Small decrease only in $m_0$** | **J02 and J03 dominate** | **S14** |
| Big decrease only in $m_1$ | J12 and J13 dominate | 5, S15 |
| Decrease $m_1$ & $m_3$ | J13 dominates | 5 |
| Decrease $m_0$&$m_1$ or $m_2$&$m_3$ | seeded J02 still dominates | S16 |
| Decrease $m_0$&$m_2$ or $m_1$&$m_3$ | J02 still dominates; much higher prevalence | S16 |
| **Set $m_0 = m_2 = 0$** | **Only J02 present; no mutation** | **S17** |
| High $m_i$'s, low $m_{ij}$'s | Little change | S18 |
| **High $m_{ij}$'s, low $m_i$'s** | **Mutation more likely to nearby strain**<br>**(1 allele away) than to complementary**<br>**strain (2 alleles away)** | **10,11** |
| initial seed change | J02 & J13 dominate or J12 & J03 dominate | S19 |
| **Seed all but J03, and** |  |  |
| **increase allele 0** |  |  |
| **waning rate** | **J03 strengthens** | **S20** |
| small $\Delta$ in drift rate $f$ | Little change | S21 |
| Increase only $f_{01}$ | Early 12 & 03 peak, then 02 & 13 | 10 |
| $f$ large ($>0.01$) | Strains equalize in strength | S22 |
| Increase all ADE $d_i$'s | J13 peak $>$ J02 peak | 12, S23, S24 |
| low $b_i$, high $d_j$ | no epidemic, $R_0 < 1$ | S25 |

