## Supplemental Material Text and Figures for "An allele-based model of coronavirus evolution under population immunity"

This Supplementary Material contains the explicit differential equations behind our model and text and figures that were deleted from our original version to improve the efficiency of our current presentation. Part I is an explicit presentation of the system of differential equations in our model. Part II presents quotations from the literature that motivated and guided our allele-based modeling. Part III contrasts different approaches of aggregating allele-based transmissibility to strain transmissibility. Part IV provides a more detailed discussion of the prevalence curves and susceptibility curves in our Basic Model. Part V is the heart of our presentation: a systematic study of the effects of changing each of the model parameters from the values in the Basic Model. Part VI expands on our presentation of possible expansions and extensions of our two-site model. Part VII lists the references for the citations in this Supplement. Part VIII presents the Python code for numerical solution of the system of differential equations presented in Part I.

### I. The Differential Equations

We no present the complete system of differential equations for our model. We begin by reminding the reader of the model's notation.

$S = R(3,3,3,3)$ = size of the compartment of the never-infected

$R(K_0,K_1,K_2,K_3)$ = size of the compartment of previously infected susceptible populations whose current state of immunity to allele i is $K_i$. Sometimes written in vector notation as $R(\mathbf{K})$.

$I$(i, j; $H_h$,$H_m$) = size of compartment for those infected with virus whose active alleles are i (0 or 1) and j (2 or 3), and whose immune states for the corresponding inactive alleles h and m are $H_h$ and $H_m$. Sometimes written as $I(s;\boldsymbol{H})$. Note that we use a comma to separate the designation of different sites and we use a semicolon to separate different lists.

J(i,j) =∑{ $I$(i, j : $H_h$, $H_m$) ; all $H_h$, $H_m$, where h and m are the inactive alleles for strain i,j}

= number of all those infected with the virus strain whose active alleles are i and j, independent of background immune status.

In our Ordinary Differential Equation (ODE) model, two sites define the virus.

This still gives us $1+ 2^6 +4^4 = 321$ compartments:

1 S-compartment + $2^6$ I-compartments + $4^4$ R-compartments.

By our definition of a strain, if both alleles at a site are in immunity state 3 (never infected), both alleles at the other site must also be in immunity state 3. This reduces the number of R-compartments by 30. Furthermore, compartment R(3,3,3,3) is a stand-in for compartment S. So, all in all, there are 290 compartments, and therefore a system of 290 differential equations.

**Transmission rates for the ODE**

$\mathcal{G}(0, 2 ; K_0, K_1, K_2, K_3)$ below is the transmission rate when a member of J(0, 2) contacts a member of compartment R($K_0$, $K_1$, $K_2$, $K_3$), without allowing for mutation.

Result is possibly a new member of I(0,2; $K_1$, $K_3$).

If there is no drift, all the increases to I(0,2; $K_1$, $K_3$) = sum of all $\mathcal{G}(0, 2; K_0, K_1, K_2, K_3)$ over all $K_0$, $K_2$.

$\mathcal{G}(0, 2 ; K_0, K_1, K_2, K_3) \equiv$

$$((\beta_0 + \beta_2 + e_{02} + \delta_0 d_0 + \delta_1 d_1 + \delta_2 d_2 + \delta_3 d_3)$$

$$\times(1 - m_0\kappa_0)(1 - m_2\kappa_2)(1 - m_{02}\kappa_0\kappa_2)$$

$$\times J(0,2) \cdot R(K_0, K_1, K_2, K_3) \cdot c,$$

where $\delta_i = 0$, if $K_i = 3$, and $\delta_i = 1$, if $K_i = 0, 1$, or 2,

$\kappa_i = 0$ if $K_i = 3$; $\kappa_i = 1$ if $K_i = 2$ or 1, and $\kappa_i = \nu_i$ if $K_i = 0$.

The first line in the definition of $\mathcal{G}$ is the pure transmission factor

(transmission terms + ADE terms),

The second line is the susceptibility multiplier,

In the third line we multiply the first two lines by

the number of those infected by strain 02,

the number of susceptibles under consideration, and

the contact rate c.

For the other three strains:

$$\mathcal{G}(1, 2\,; K_0, K_1, K_2, K_3) \equiv \quad ((\beta_1 + \beta_2 + e_{12} + \delta_0 d_0 + \delta_1 d_1 + \delta_2 d_2 + \delta_3 d_3)$$
$$\times (1 - m_1\kappa_1)(1 - m_2\kappa_2)(1 - m_{12}\kappa_1\kappa_2)$$
$$\times J(1,2) \cdot R(K_0, K_1, K_2, K_3) \cdot c,$$

$$\mathcal{G}(0, 3\,; K_0, K_1, K_2, K_3) \equiv \quad ((\beta_0 + \beta_3 + e_{03} + \delta_0 d_0 + \delta_1 d_1 + \delta_2 d_2 + \delta_3 d_3)$$
$$\times (1 - m_0\kappa_0)(1 - m_3\kappa_3)(1 - m_{03}\kappa_0\kappa_3)$$
$$\times J(0,3) \cdot R(K_0, K_1, K_2, K_3) \cdot c,$$

$$\mathcal{G}(1, 3\,; K_0, K_1, K_2, K_3) \equiv \quad ((\beta_1 + \beta_3 + e_{13} + \delta_0 d_0 + \delta_1 d_1 + \delta_2 d_2 + \delta_3 d_3)$$
$$\times (1 - m_1\kappa_1)(1 - m_3\kappa_3)(1 - m_{13}\kappa_1\kappa_3)$$
$$\times J(1,3) \cdot R(K_0, K_1, K_2, K_3) \cdot c,$$

#### We now include drifts in the dI(0, 2; $K_1$, $K_3$)/dt expression.

There are two kinds of drift/transmission terms in the expression for dI(0, 2; $K_1$, $K_3$)/dt:

1) Terms to indicate drift from another strain into strain 02 upon transmission, and
2) Terms to include transmissions from strain 02 to strain 02 that could have mutated but did not.

Write $f_{ij}$ for the mutation rate for a mutation from allele i to allele j at site 0 or site 1. Recall that such a mutation can only take place if the infectee has low immunity for allele j, i.e., $K_j = 0$ or 3.

**Transmission With Mutation**

To include all the new infections that can mutate to strain 12, write:

$\mathcal{C}(1, 2; K_0, K_3) \equiv$

$\sum \{\mathcal{G}(0, 2; K_0, H_1, H_2, K_3)\ f_{01} |\ H_1=0 \text{ or } 3, \text{ all } H_2\}$ +

$\sum \{\mathcal{G}(1, 3; K_0, H_1, H_2, K_3) \cdot f_{32} \mid H_1 \text{ all}, H_2 = 0 \text{ or } 3\}$ +

$\sum \{\mathcal{G}(0, 3; K_0, H_1, H_2, K_3) \cdot f_{01} \cdot f_{32} |\ H_1 = 0 \text{ or } 3, \text{ and } H_2 = 0 \text{ or } 3\}$.

The first term covers all those 02s who could have mutated to 12 ($H_1$ = 0 or 3) and did, with probability $f_{01}$. The second term covers all those 13s who could have mutated to 12 ($H_2$ = 0 or 3) and did, with probability $f_{32}$. The third term covers all those 03s who could have mutated to 12 ($H_1$ = 0 or 3 and $H_2$ = 0 or 3) and did, with probability $f_{01}f_{32}$.

$\mathcal{C}(0, 2; K_1, K_3) \equiv$

$\sum \{\mathcal{G}(1, 2; H_0, K_1, H_2, K_3) \cdot f_{10} \mid H_0 = 0 \text{ or } 3, H_2 \text{ all}\}$ +

$\sum \{\mathcal{G}(0, 3; H_0, K_1, H_2, K_3) \cdot f_{32} \mid H_0 \text{ all}, H_2 = 0 \text{ or } 3\}$ +

$\sum \{\mathcal{G}(1, 3; H_0, K_1, H_2, K_3) \cdot f_{10}\, f_{32} \mid H_0 = 0 \text{ or } 3, \text{ and } H_2 = 0 \text{ or } 3\}.$

$\mathcal{C}(0, 3; K_1, K_2) \equiv$

$\sum \{\mathcal{G}(1, 3; H_0, K_1, K_2, H_3) \cdot f_{10} \mid H_0 = 0 \text{ or } 3, H_3 \text{ all}\} +$

$\sum \{\mathcal{G}(0, 2; H_0, K_1, K_2, H_3) \cdot f_{23} \mid H_0 \text{ all}, H_3 = 0 \text{ or } 3\} +$

$\sum \{\mathcal{G}(1, 3; H_0, K_1, K_2, H_3) \cdot f_{10}\, f_{23} \mid H_0 = 0 \text{ or } 3, \text{ and } H_3 = 0 \text{ or } 3\}.$

$\mathcal{C}(1, 3; K_0, K_2) \equiv$

$\sum \{\mathcal{G}(1, 2; K_0, H_1, K_2, H_3) \cdot f_{23} \mid H_1 \text{ all}, H_3 = 0 \text{ or } 3\} +$

$\sum \{\mathcal{G}(0, 3; K_0, H_1, K_2, H_3) \cdot f_{01} \mid H_1 = 0 \text{ or } 3, H_3 \text{ all}\} +$

$\sum \{\mathcal{G}(0, 2; K_0, H_1, K_2, H_3) \cdot f_{01}\, f_{23} \mid H_1 = 0 \text{ or } 3, \text{ and } H_3 = 0 \text{ or } 3\}.$

**Transmission Without Mutation**

Next, we include the transmission terms in which no mutation occurs at transmission to

$I(0, 2; K_1, K_3)$, though such a mutation could have occurred. We start with a contact between a member of $J(0,2)$ and a member of a general $R(K_0, K_1, K_2, K_3)$.

$\Psi(0, 2; K_1, K_3) =$

$\sum \{\mathcal{G}(0, 2; H_0, K_1, H_2, K_3) \cdot (1-f_{01}) \mid H_0 \text{ all}, H_2 \text{ all}\},$ if $K_1 = 0 \text{ or } 3, K_3 = 1 \text{ or } 2.$

$\sum \{\mathcal{G}(0, 2; H_0, K_1, H_2, K_3) \cdot (1-f_{23}) \mid H_0 \text{ all}, H_2 \text{ all}\},$ if $K_1 = 1 \text{ or } 2, K_3 = 0 \text{ or } 3$

$\sum \{\mathcal{G}(0, 2; H_0, K_1, H_2, K_3) \cdot (1-f_{01}) \cdot (1-f_{23}) \mid H_0 \text{ all}, H_2 \text{ all}\},$ if $K_1 = 0 \text{ or } 3, K_3 = 0 \text{ or } 3,$

$\sum \{\mathcal{G}(0, 2; H_0, K_1, H_2, K_3) \mid H_0 \text{ all}, H_2 \text{ all}\}$, if $K_1 = 1$ or $2, K_3 = 1$ or $2$

In the first case, drift from 02 to 12 can occur because $K_1$ is weak, but does not occur -- with frequency $(1-f_{01})$.

In the second case, drift from 02 to 03 can occur because $K_3$ is weak, but does not occur -- with frequency $(1-f_{23})$.

In the third case, drift from 02 to 13 can occur because $K_1$ and $K_3$ are weak, but does not.

In the fourth case, $K_1$ and $K_3$ are too strong to allow a mutation.

There are similar expressions for the other three strains resisting mutation:

$\Psi(1, 2; K_0, K_3) =$

$\sum \{\mathcal{G}(1, 2; K_0, H_1, H_2, K_3)\cdot(1-f_{10}) \mid H_1 \text{ all}, H_2 \text{ all}\}$ if $K_0$ is 0 or 3 and $K_3$ is 1 or 2,

$\sum \{\mathcal{G}(1, 2; K_0, H_1, H_2, K_3)\cdot(1-f_{23}) \mid H_1 \text{ all}, H_2 \text{ all}\}$ if $K_0$ is 1 or 2 and $K_3$ is 0 or 3,

$\sum \{\mathcal{G}(1, 2; K_0, H_1, H_2, K_3)\cdot(1-f_{10})\cdot(1-f_{23}) \mid H_1 \text{ all}, H_2 \text{ all}\}$ if $K_0$ is 0 or 3 and $K_3$ is 0 or 3,

$\sum \{\mathcal{G}(1, 2; K_0, H_1, H_2, K_3) \mid H_1 \text{ all}, H_2 \text{ all}\}$ if $K_0$ is 1 or 2 and $K_3$ is 1 or 2.

$\Psi(0, 3; K_1, K_2) =$

$\sum \{\mathcal{G}(0, 3; H_0, K_1, K_2, H_3)\cdot(1-f_{01}) \mid H_0 \text{ all}, H_3 \text{ all}\}$ if $K_1$ is 0 or 3 and $K_2$ is 1 or 2,

$\sum \{\mathcal{G}(0, 3; H_0, K_1, K_2, H_3)\cdot(1-f_{23}) \mid H_0 \text{ all}, H_3 \text{ all}\}$ if $K_1$ is 1 or 2 and $K_2$ is 0 or 3,

$\sum \{\mathcal{G}(0, 3; H_0, K_1, K_2, H_3)\cdot(1-f_{01})\cdot(1-f_{23}) \mid H_0 \text{ all}, H_3 \text{ all}\}$ if $K_1$ is 0 or 3 and $K_2$ is 0 or 3,

$\sum \{\mathcal{G}(0, 3; H_0, K_1, K_2, H_3) \mid H_0 \text{ all}, H_3 \text{ all}\}$ if $K_1$ is 1 or 2 and $K_2$ is 1 or 2.

$\Psi(1, 3; K_0, K_2) =$

$\sum \{\mathcal{G}(1, 3; K_0, H_1, K_2, H_3)\cdot(1-f_{10}) \mid H_1 \text{ all}, H_3 \text{ all}\}$ if $K_0$ is 0 or 3 and $K_2$ is 1 or 2,

$\sum \{\mathcal{G}(1, 3; K_0, H_1, K_2, H_3)\cdot(1-f_{32}) \mid H_1 \text{ all}, H_3 \text{ all}\}$ if $K_0$ is 1 or 2 and $K_2$ is 0 or 3,

$\sum \{\mathcal{G}(1, 3; K_0, H_1, K_2, H_3)\cdot(1-f_{10})\cdot(1-f_{32}) \mid H_1 \text{ all}, H_3 \text{ all}\}$ if $K_0$ is 0 or 3 and $K_2$ is 0 or 3,

$\sum \mathcal{G}(1, 3; K_0, H_1, K_2, H_3) \mid H_1 \text{ all}, H_3 \text{ all}\}$ if $K_0$ is 1 or 2 and $K_2$ is 1 or 2.

#### Recovery Function L:

We next consider the dynamics of recovery from infection as it impacts the R-compartments. Recall that an individual who recovers from, say a strain 02 infection, moves from $I(0,2;K_1,K_3)$ to $R(2,K_1,2,K_3)$. Therefore, R-compartments with exactly zero or one $K_i = 2$ receive no recoveries. If exactly two $K_i$'s = 2, the corresponding alleles i must lie in distinct viral sites. This leaves four cases to consider. Recall that $v_{ij}$ = recovery rate from strain ij.

$L(K_0, K_1, K_2, K_3)$ = rate of increase in $R(K_0, K_1, K_2, K_3)$ from recovering infections:

If $\{K_0 \neq 2 \text{ and } K_1 \neq 2\}$, OR $\{K_2 \neq 2 \text{ and } K_3 \neq 2\}$: then $L(K_0, K_1, K_2, K_3) = 0$,

If $K_1 \neq 2, K_3 \neq 2$: then $L(2, K_1, 2, K_3) = v_{02}\cdot I(0, 2; K_1, K_3)$

If $K_0 \neq 2, K_3 \neq 2$: then $L(K_0, 2, 2, K_3) = v_{12}\cdot I(1, 2; K_0, K_3)$

If $K_1 \neq 2$, $K_2 \neq 2$: then $L(2, K_1, K_2, 2) = v_{03} \cdot I(0, 3; K_1, K_2)$

If $K_0 \neq 2$, $K_2 \neq 2$: then $L(K_0, 2, K_2, 2) = v_{13} \cdot I(1, 3{:}\ K_0, K_2)$

If $K_0 \neq 2$: then $L(K_0, 2, 2, 2) = v_{12} \cdot I(1, 2; K_0, K_3{=}2) + v_{13} \cdot I(1, 3; K_0, K_2{=}2)$

If $K_1 \neq 2$: then $L(2, K_1, 2, 2) = v_{02} \cdot I(0, 2; K_1, K_3{=}2) + v_{03} \cdot I(0, 3; K_1, K_2{=}2)$

If $K_2 \neq 2$: then $L(2, 2, K_2, 2) = v_{03} \cdot I(0, 3; K_1{=}2, K_2) + v_{13} \cdot I(1, 3; K_0{=}2, K_2)$

If $K_3 \neq 2$: then $L(2, 2, 2, K_3) = v_{02} \cdot I(0, 2; K_1{=}2, K_3) + v_{12} \cdot I(1, 2; K_0{=}2, K_3)$

$L(2, 2, 2, 2) = \sum_{\{h=0,1;j=2,3\}} v_{hj} \cdot I(h, j; 2, 2).$

This whole scenario can be summarized by the expression:

$$L(K_0, K_1, K_2, K_3) = \sum_{h\epsilon\{0,1\}} \sum_{j\epsilon\{2,3\}} \mathbf{1}\{K_h = 2\} \cdot \mathbf{1}\{K_j = 2\} \cdot \mathrm{v_{hj}} \cdot I(h, j; K_{1-h}, K_{5-j}),$$

where $\mathbf{1}\{K_h = 2\}$ is the indicator function for $\mathrm{K_h} = 2$:

$$\mathbf{1}\{K_h = 2\} = 1, \text{if } K_h = 2; \text{ and } \mathbf{1}\{K_h = 2\} = 0 \text{ otherwise.}$$

### Immunity Strength Transitions, including Waning

Immunity can decrease at each allele site in the recovered susceptible populations

$\boldsymbol{R}(K_0, K_1, K_2, K_3)$.

Let $\omega_a(K)$ be the rate at which immunity decreases for allele a when their current level of immunity is K. In the text, we write this as $\omega_{aK}$.

$\omega_a(0) = 0$ for all alleles a; we assume those at level 0 have already waned.

$\omega_a(3) = 0$ for all alleles a; the never infected with immunity status 3 have no immunity to wane.

For notation's sake, we include $\omega_a(4) = 0$; there is no immunity level 4, but the notation will be helpful.

The waning immunity terms in the differential equations for $\boldsymbol{R}(K_0, K_1, K_2, K_3)$ are:

$W(K_0, K_1, K_{2,}, K_3). =$

$-(\omega_0(K_0) + \omega_1(K_1) + \omega_2(K_2) + \omega_3(K_3))\ \boldsymbol{R}(K_0, K_1, K_2, K_3)$

$+ \omega_0(K_0+1)\ \boldsymbol{R}(K_0+1, K_1, K_2, K_3)$

$+ \omega_1(K_1+1)\ \boldsymbol{R}(K_0, K_1+1, K_2, K_3)$

$+ \omega_2(K_2+1)\ \boldsymbol{R}(K_0, K_1, K_{2,}+1, K_3)$

$+ \omega_3(K_3+1)\ \boldsymbol{R}(K_0, K_1, K_{2,}, K_3+1)$

The first term after the equal sign presents the decrease to a lower level of each antibody response, thus decreasing the size of compartment $\boldsymbol{R}(K_0, K_1, K_2, K_3)$

The second term after the equal sign catches the "increase" in $\boldsymbol{R}(K_0, K_1, K_2, K_3)$ as those with $K_0+1$ level immunity in allele 0 in $\boldsymbol{R}(K_0+1, K_1, K_2, K_3)$ undergo "waning," and so on.

By definition, $\boldsymbol{R}(K_0, K_1, K_2, K_3) = 0$ if any $K_a=4$.

### Vaccination

We have turned off vaccination in our analysis in the text: q=0. We include here for future reference. We assume vaccines only affected allele 0 and that fraction q of the population per year were vaccinated, starting just after the first epidemic peak. Vaccinations moved susceptibles in compartment $\boldsymbol{R}(K_0, K_1, K_2, K_3)$ into compartment $\boldsymbol{R}(2, K_1, K_2, K_3)$. The recently recovered in $\boldsymbol{R}(2, K_1, K_2, K_3)$ are not vaccinated.

$\frac{d\boldsymbol{R}(K_0,K_1,K_2,K_3)}{dt} = -q \cdot \mathrm{R}(K_0, K_1, K_2, K_3)$ for all $(K_0, K_1, K_2, K_3)$ including $K_0=2$

$$\frac{d\boldsymbol{R}(2, K_1, K_2, K_3)}{dt} = q \cdot [\mathrm{R}(3, K_1, K_2, K_3) + \mathrm{R}(2, K_1, K_2, K_3) + \mathrm{R}(1, K_1, K_2, K_3) + \mathrm{R}(0, K_1, K_2, K_3)]$$

#### The Complete Differential Equations

We can now write out the complete system of differential equations for our model:

#### Susceptible-Compartment Dynamics

$$\frac{dR(3,3,3,3)}{dt} =$$

(births/deaths) $$\mu\left[\sum_{i=0,1;j=2,3} J(i,j) + \sum_{\substack{K_0,K_1,K_2,K_3=0,1,2,3\\ K_0+K_1\neq 6,\, K_2+K_3\neq 6}} R(K_0,K_1,K_2,K_3)\right]$$

(infections)

$$-\boldsymbol{c}\cdot R(3,3,3,3)\cdot J(0,2)\cdot(\beta_0+\beta_2+e_{02})$$

$$-\boldsymbol{c}\cdot R(3,3,3,3)\cdot J(1,2)\cdot(\beta_1+\beta_2+e_{12})$$

$$-\boldsymbol{c}\cdot R(3,3,3,3)\cdot J(0,3)\cdot(\beta_0+\beta_3+e_{13})$$

$$-\boldsymbol{c}\cdot R(3,3,3,3)\cdot J(1,3)\cdot(\beta_1+\beta_3+e_{03})$$

(vaccination) $$-\mathrm{q}\cdot \mathrm{R}(3,3,3,3)$$

For all $K_0, K_1, K_2, K_3 = 0, 1, 2, 3$, such that $K_0 + K_1 \neq 6$ $and$ $K_2 + K_3 \neq 6$,

$$\frac{d\boldsymbol{R}(K_0,K_1,K_2,K_3)}{dt} =$$

(recovery) L$(K_0, K_1, K_2, K_3)$

(waning) + W$(K_0, K_1, K_2, K_3)$

(Background death) $-\mu \cdot \boldsymbol{R}(K_0, K_1, K_2, K_3)$

(New infection) $-\mathcal{G}(0,2;\ K_0, K_1, K_2, K_3) - \mathcal{G}(0,3;\ K_0, K_1, K_2, K_3)$

$-\mathcal{G}(1,2\ ; K_0, K_1, K_2, K_3)\ \ - \mathcal{G}(1,3\ ;\ K_0, K_1, K_2, K_3)$

(Vaccination) $= -\mathrm{q} \cdot \mathrm{R}(K_0, K_1, K_2, K_3)$

$+ \gamma(K_0) \cdot q \cdot [\mathrm{R}(3, K_1, K_2, K_3) + \mathrm{R}(2, K_1, K_2, K_3)\ + \mathrm{R}(1, K_1, K_2, K_3)\ + \mathrm{R}(0, K_1, K_2, K_3)\ ],$

where $\gamma(K_0) \coloneqq \begin{cases} 1, & \text{if } K_0 = 2, \\ 0, & \text{otherwise.} \end{cases}$

### Infection Dynamics

$$\frac{dI(0,2; K_1, K_3)}{dt} =$$

(New infection) $\Psi(0,2;\ K_1, K_3) + \mathcal{C}(0,2;\ K_1,\ K_3)$

(Recovery) $-\ v_{02} \cdot \boldsymbol{I}\ (0,2;\ K_1, K_3)$

(Background death) $-\mu \cdot \boldsymbol{I}\ (0,2;\ K_1, K_3)$

$$\frac{dI(0,3; K_1, K_2)\ )}{dt} = \Psi(0,3;\ K_1,\ K_2) + \mathcal{C}(0,\ 3;\ K_1,\ K_2) - \mathrm{v}_{03} \cdot \boldsymbol{I}\ (0,3;\ K_1,\ K_2) - \mu \cdot I\ 0,3;\ K_1,\ K_2)$$

$$\frac{dI(1,2;K_0,K_3))}{dt} = \Psi(1,2;K_0,K_3) + \mathcal{C}(1,2;K_0,K_3) - \mathrm{v}_{12}\cdot I(1,2;K_0,K_3) - \mu\cdot \boldsymbol{I}(1,2;K_0,K_3)$$

$$\frac{dI(1,3;K_0,K_3)}{dt} = \Psi(1,3;K_0,K_2) + \mathcal{C}(1,3;K_0,K_2) - \mathrm{v}_{13}\cdot I(1,3;K_0,K_2) - \mu\cdot I(1,3;K_0,K_2)$$

#### Parameter values for baseline run.

| Parameters in model | Base value | |
|---|---|---|
| $b_j$ = transmissibility of allele j | $b_0 = b_1 = b_2 = b_3$ | = 0.3 |
| $b_{jk}$ = joint transmissibility of alleles j and k | $e_{02} = e_{12} = e_{03} = e_{13}$ | = 0.0 |
| $v_{jk}$ = rate of recovery from infection by strain jk | $v_{02} = v_{12} = v_{03} = v_{13}$ | = 0.1 |
| $m_j$ = relative full immunity strength | | |
| to allele j | $m_0 = m_1 = m_2 = m_3$ | =0.7 |
| $m_{jk}$ = relative full joint immunity strength | | |
| to alleles j and k | $m_{02} = m_{12} = m_{03} = m_{13}$ | =0.0 |
| $\nu_j$ = relative fully waned immunity | | |
| strength to allele j | $\nu_0 = \nu_1 = \nu_2 = \nu_3$ | =0.05 |
| $f_{jk}$ = probability of drift from allele j to | $f_{01} = f_{10}$ | =0.00001 |
| allele k at same site | $f_{23} = f_{32}$ | =0.00001 |
| $w_{j2}$= transition rate for allele j from | | |
| immune state 2 to immune state 1 | $w_{02} = w_{12} = w_{22} = w_{32}$ | =0.003 |
| $w_{j1}$= waning rate for allele j from | | |

| | | |
|---|---|---|
| immune state 1 to immune state 0 | $w_{01} = w_{11} = w_{21} = w_{31}$ | =0.003 |
| $d_j$ = A.D.E. transmission factor for allele j | $d_0 = d_1 = d_2 = d_3$ | = 0.0 |
| c = effective contact rate | c | =0.5 |
| mu = population birth/death rate | mu | =0.0001 |
| q = vaccination rate | q | =0 |

Initial conditions

R(3,3,3,3) = 0.99999, I(0,2; 3,3) = 0.00001, Rest = 0

Total Population should stay constant at 1.

### II. Quotations that have motivated our work

**Importance of a population approach:**

It is not just differences in infection forces experienced by individuals that generate new infection patterns. How those forces generate different population patterns of immunity is also important. Saad-Roy et al [Sa20]:

> "…relying on the status of infection of an individual as the main observable during an ongoing epidemic is insufficient to characterize the complex immune landscape generated by the pandemic."

**Importance of including virus components in a population model:**

This paper addresses the challenge of Saad-Roy, Metcalf, and Grenfell [Sa22] for

> "epidemiological studies that simultaneously quantify immunity and transmission across populations and at different biological scales … along with the development of cross-scale modeling approaches."

**Importance of including immune response:**

Saad-Roy and colleagues [Sa20, Sa21a, Sa22, Sa24a] argue:

> “variations in the immune response to primary SARS-CoV-2 infections… lead to markedly different immune landscapes and burdens of critical cases…, and illustrate likely complexities in future CoV-19 dynamics..” [Sa20]

And in [Sa24]:

> “Using simple mathematical models, we investigate the medium-term impacts of waning immunity against severe disease on immuno-epidemiological dynamics…. Our results illustrate the necessity to characterize both transmission-blocking and severity-blocking immune time scales.”
>
> “We have determined that the strength of immunity is a central parameter that shapes medium-term immuno-epidemiological dynamics.”
>
> “Our simple models reveal that a large range of outcomes can emerge from uncertainties in both the duration of severity-blocking immunity and the strength of immunity, and from the confluence of these two parameters. In particular, our findings emphasize that the strength of immunity shapes immuno-epidemiological dynamics at multiple resolutions, and that the duration of severity-blocking immunity has a major effect on population-level immune landscapes and potential burdens.”

**No immune response → no selection**

[Sa22]: “if there is no immune pressure, then viral abundance may be high but selection for immune escape is absent.”

**Importance of including measures of population immunity:**

[Me23]: “SARS-CoV-2 variants compete on a fitness landscape shaped by population immunity.”

[Xi26]: “Measuring population immunity is crucial for epidemic preparedness, but methods to translate individual immunity into population-level immunity metrics remain underdeveloped.”.

**Applying the Saad-Roy framework**

[Sa24a]: Saad-Roy and colleagues have, in their own words:

- shown the importance of "the relative susceptibility to infection after waning …(as) a key determinant of post-pandemic trajectories" [Sa20],
- "incorporated two-dose vaccines" into this framework [Sa21b],
- "estimated the potential effects of vaccine nationalism" [Wa21], and
- "examined the impact of accumulating immunity on the potential future burden of chronic disease" [Sa23] ,

all in the framework of the infections and reinfections of a *single strain*. They added a second strain as an "immune-escape variant" in [Sa23], as did [Dys21].

#### III. What is a natural way to formulate $b_{hi}$ in terms of $b_h$ and $b_i$?

In a sense, the functional form is not so important. We will start in our *basic model* with equal $b_h$'s and equal $b_{hi}$'s and be concerned with the impact of both large and small *changes* in individual b's. In an allele-based approach, a small change in, say, $b_0$ would be reflected in small changes of the basic values of *both* $b_{02}$ and $b_{03}$. In a strain-based approach, we would simply change the value of the $b_{hi}$ for the strain (h,i) under consideration.

For simplicity's sake, we compare $b_0 + b_2$ and $b_0 \cdot b_2$. Multiplication would be natural if we were parameterizing frequencies in the sense of Hardy-Weinberg Equilibrium [H08, W08]; after all, Prob(AB) ≈ Prob(A)·Prob(B). On the other hand, following Fisher [F18], the additive version is more natural if we are interested in aggregating effects. Since our concerns are more in line with effects than with frequencies, we use the simpler additive formulation in our model. Once again, for our concerns with incremental changes, it really does not make a difference.

#### IV. More complete description of the nuances in the prevalence curves and susceptibility curves in Figures 2 and 3:

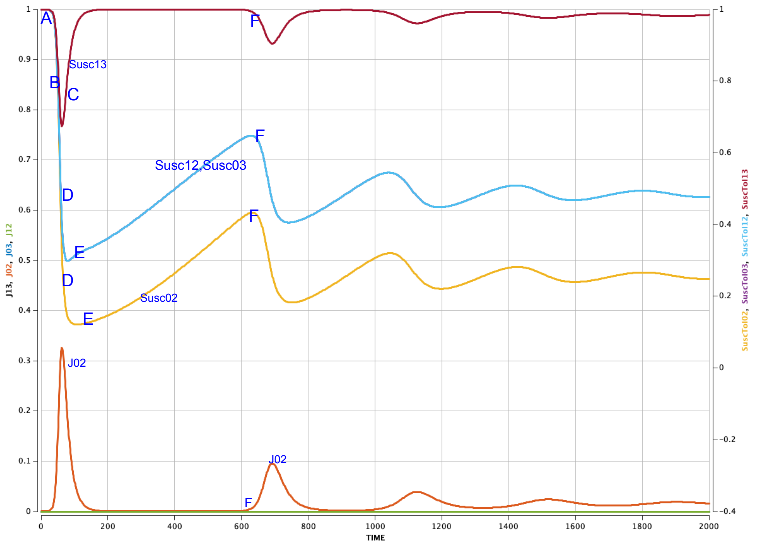

Figure 2: The graphs of strain 02 and of the four susceptibility curves for the no-drift situation.

Despite the simplicity of this one-strain situation, the patterns in Figure 2 occur regularly, and so we describe them here in some detail. The letters on the left refer to the letters in Figure 2.

(A) For t near 0, nearly everyone is in pure susceptible compartment R3333, i.e., compartment S. All the susceptibility curves start near 1 since R3333 is a component of all them.

(B) With this high level of susceptibility, the existence of even a few strain-02 infectives leads to the rapid growth of strain 02, leading to a peak prevalence of ~0.326 at t =65. As the new infections spread, the size of compartment S = R3333 decreases rapidly and so do all the Susceptibility Curves.

(C) The recovered 02-infectives move to R2323 and eventually -- through waning immunity – into R2313, R1323, and R1313. These Rx3y3's and the not-yet-infected R3333's are all full components of Susc13 (the Susceptibility curve for strain 13) and only of Susc13. Around t=65, the growth of these Rx3y3 compartments outpaces the number of new 02 infections; Susc13 begins to rise dramatically, nearly all the way back to level 1.

(D) These Rx3y3's play only a minor role in Susc12 and Susc03 and almost no role at all in Susc02. So these latter susceptibility curves keep falling while Susc13 rises, with Susc12 and Susc03 larger than Susc02.

(E) At some point (t=108), the population immunity reaches a point where the number of 02-infectives are few and far between (~0.04) and new infections are outpaced by new "births" into R3333. This turn-around allows the Susc12, Susc03 and Susc02 curves to begin to rise and to keep rising roughly at the birth rate. This rise is eventually abetted by the full waning of the recovered R2323's to compartment R0303, also a component of all 4 susceptibility curves.

(F) While Susc02 is rising, there are very few strain 02 infectives. However, at some point (t=633) the number of infected with strain 02 times the population susceptibility Susc02 reaches a threshold so that a new strain 02 epidemic wave begins. The new infections in this wave decrease the sizes of R3333 and R0303 so that all four susceptibility curves begin to decline, and the cycle continues.

Given the high susceptibility of strain 13 after the 02 epidemic wave, it would take just a few 13 infections to set off a 13 epidemic wave, as we will soon see.

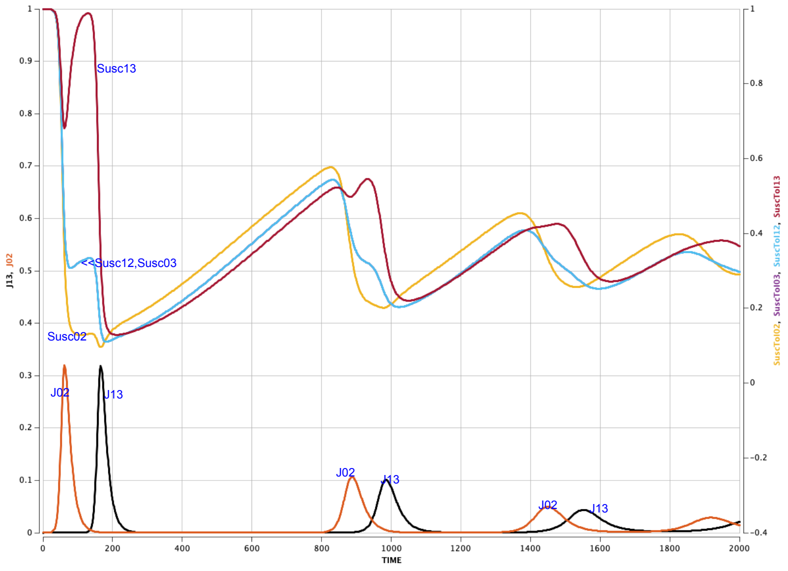

Figure 3. The strain prevalences and the susceptibility curves for the basic parameters.

Figure 3 includes both the four strain prevalences and the "four" susceptibility curves. The latter are pretty much as they are in figure 2, with the obvious exception of Susc13. We see strains 02 and 13 alternating as epidemic waves, with a moderately long gap of negligible infection after each 13 episode. Strains 03 and its complement 12 are barely ever present.

We present the dynamics in more detail:

- Prevalence of strain 02 first peaks at around 32% at t= 62, just as it did in Figure 2. By t=100, over 75% of the population has experienced and recovered from strain 02. (R2323 is at 75% at t=100.) Strain 02 prevalence then bottoms out at 0.000002% around t=425, before achieving its second wave with a peak of ~11% at t=883.
- Meanwhile its complementary strain 13 is invisible until t=52; it does take two mutations to generate a 13 from a 02. But at t=130, susceptibility for 13 reaches almost 100%, so it takes just a few 13s to generate a 13 epidemic wave. This 13-wave peaks at ~32% at t=166, before strain 13 begins its decline, bottoming at 0.000004% around t=600. Then, once again its susceptibility rises as a result of recoveries from the second 02 wave. Strain 13 has its second peak at ~10% at t=985.
- The prevalences of strains 12 and 03 are identical for all time because they are governed by the exact same parameter values and initial conditions. Only one mutation away from the seeded 02, they appear early and peak at ~0.0007% at t=70. However, their susceptibilities do not have prior recoveries to build on, as strain 13 did, and so their prevalence times susceptibility product never reaches a threshold for epidemic takeoff. They never do re-achieve the ~0.0007% of their first "peak."

#### V. A systematic study of the effects of parameter changes on the epidemics described by The Basic Model of Figure 3.

Section Four of the text presents an overview of the effects of parameter changes on the dynamics of our baseline model. In the first part of this Supplementary Material, we present a more detailed discussion of the effects of the various parameter changes to the basic model.

We are interested in two perspectives. First, how do such changes affect the sequence of epidemic waves in the short run. Second, how do such changes affect the total dynamical system. This dynamical system is a non-linear compartmental system, and such systems are characterized by convergence of the dynamic to an equilibrium solution [JS93]. For example, despite the early oscillations presented in Figure 3, those curves converge from the initial small seed of I0233 to the steady state in which

Strain 02 = Strain13 = 0.01173624 and Strain 12 = Strain 03 = 0.00000109.

Solutions starting with the initial seed I1333 with complementary strain13 converge to the same equilibrium. If the initial seed is with Strain 12 or Strain 03, the solution converges to equilibrium solution in which

Strain 03 = Strain12 = 0.01173624 and Strain 02 = Strain 13 = 0.00000109.

Thus, there are at least two asymptotically stable equilibria; extensive simulation suggests that these are the only two. The set of initial conditions whose solution converges to a given equilibrium is called the *basin of attraction* of that equilibrium. Understanding the basins of attraction is a key step in understanding the dynamics of the pandemic under study.

**Changing Transmission Rate b**

The parameter values listed in Section 3 were chosen carefully but somewhat randomly. We examine how the graph of the baseline model in Figure 3 would look for different parameter choices. We start with the question: “What would a universally higher or lower transmission rate mean for a pandemic?”

We start with $b_i$=0.3, as in Section 3, and then look at higher and lower values of the $b_i$. Figure S1 presents the analog of Figure 3 for $b_0 = b_1 = b_2 = b_3$ all equal 0.2, 0.3, 0.4, and 0.5 with the rest of the parameters as in Section 3.

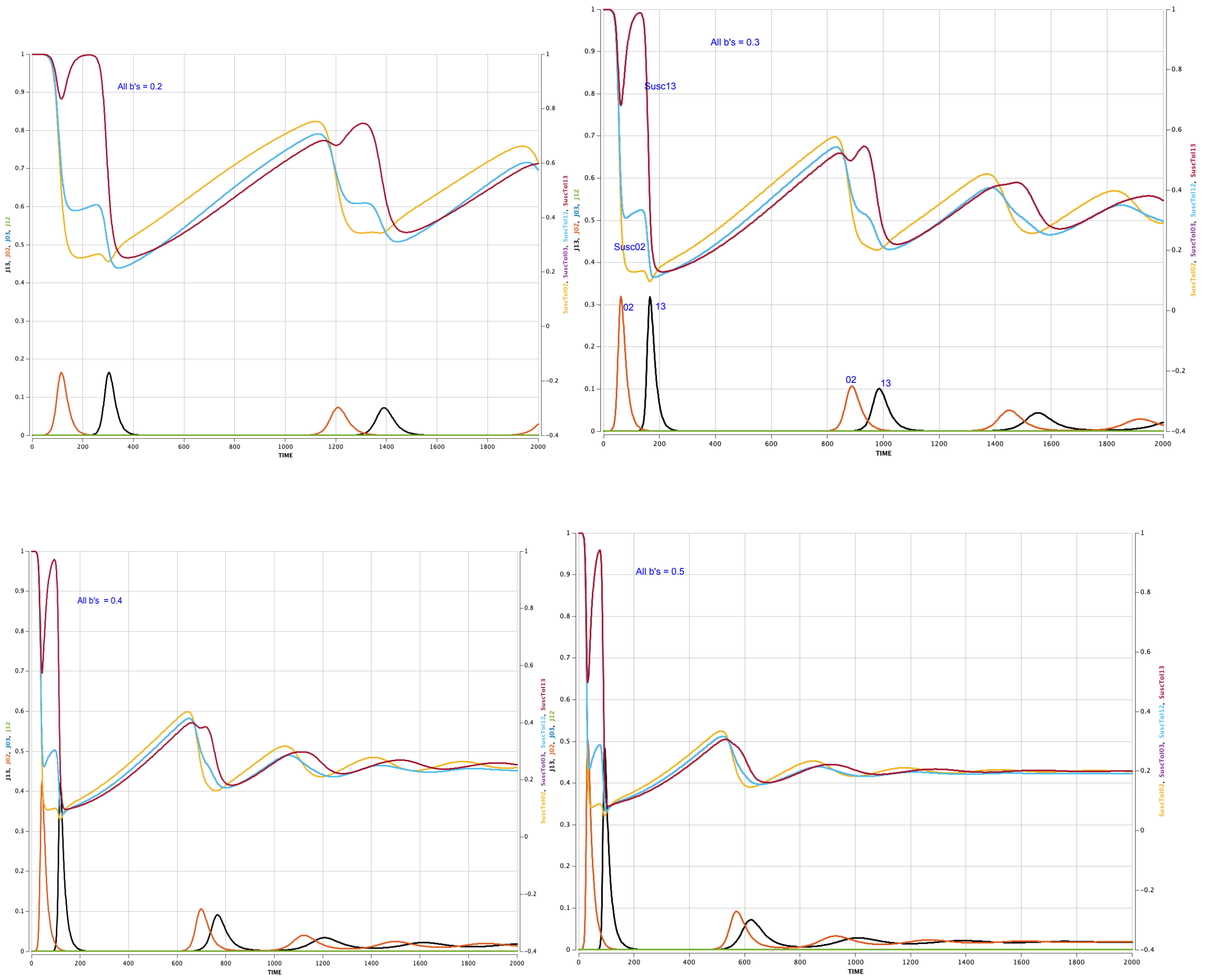

Figure S1. Basic model with all $b_i$ ‘s = 0.2, 0.3, 0.4, 0.5 (clockwise from top left).

The differences in Figures 2, 3, and S1 are interesting:

1. As transmission rate increases, more people become infected and therefore more recover in early time periods. As a result, segment A in Figure 2 is shorter and the downturn in segment B is sooner. The first strain 02 epidemic occurs earlier.
2. As the transmission rate increases relative to the recovery rate, the turn-around on the Susc13 curve occurs at a lower value and at an earlier time. The peak of the strain 02 epidemic curve is sooner and higher.
3. The peak of the Susc13 curve at F occurs sooner; the onset of the 13 epidemic occurs sooner.
4. With the increase in 02 and 13 transmissions and the resulting earlier and higher-peaked 02 and 13 epidemics, the bottom of the susceptibility curves is lower:

around 22% for $b_i$=0.2, 11% for $b_i$ = 0.3, and 7% for $b_i$=0.5.

5. But the higher transmission rate implies a lower threshold for new epidemics. It takes less time for the susceptibility to reach a threshold level for the next 02 epidemic.
   The gap between the first 02/13 epidemics and the second 02/13 epidemics grows shorter as the transmission rate increases.
6. Higher transmission rates lead to higher long run equilibria for the systems:

| All $b_i$'s | Equilibrium for 02 and 13 | Equilibrium for 12 and 03 |
|---|---|---|
| 0.2 | 0.00747092 | 0.00000092 |
| 0.3 | 0.01173624 | 0.00000109 |
| 0.5 | 0.01895679 | 0.00000210 |

In summary, higher overall transmission rates lead to sooner and higher peaked first 02- and 13-epidemics with less time between them, shorter time periods between the first 02/13 epidemics and the following two, and higher long run equilibria.

**Changing one allele-transmission-rate**

In Figure 7 in the text, we worked with $b_0$ = 0.3 slightly lower than $b_1$ = $b_2$ = $b_3$ = 0.4. In Figure S2 we lower $b_0$ even more to 0.15. Despite 02's lower transmissibility, strains 13 and 02 still dominate.

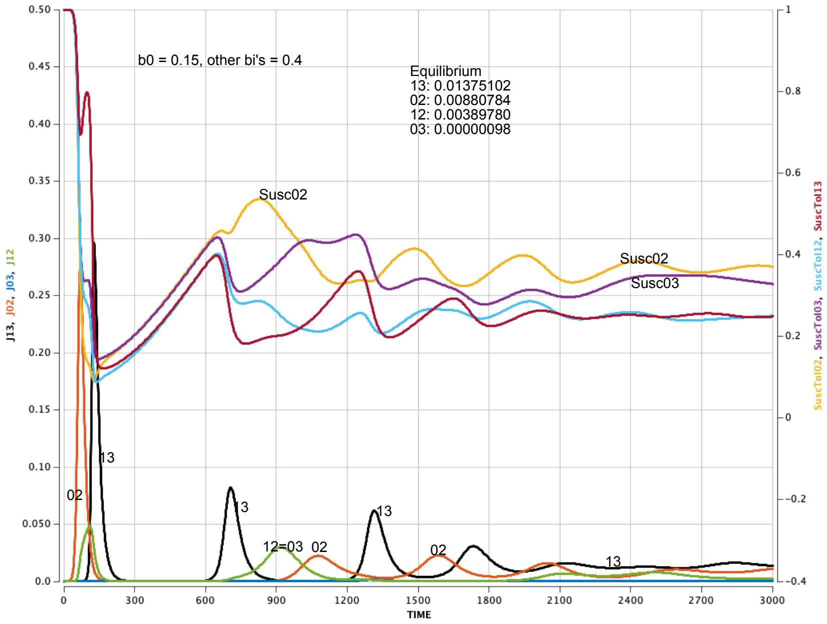

Figure S2. $b_0 = 0.15$, $b_1 = b_2 = b_3 = 0.4$

What happens when we change only $b_1$ or $b_3$, transmissibility of one of the non-seeded alleles? We work with $b_1$ (Figure S3); analogous results hold for varying only $b_3$.

Who gains most from any change in allele 1 transmissibility: strain 12 or strain 13? And how does the seeded strain 02 fare? As expected, strain 02 continues to thrive, even though it has lower transmissibility: founder effect. With this benefit, strain 13 does much better than 12 because of the complementary susceptibility effect. (Strain 13 even surpasses 02 as the third epidemic wave.) In the long run, seeded strain 02 is five orders of magnitude larger than strain 03, despite strain 03's transmissibility advantage.

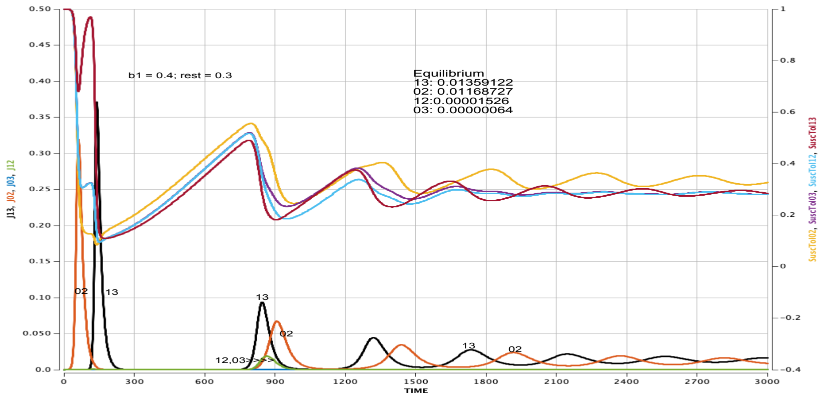

*Figure S3: $b_1 = 0.4$, $b_0 = b_2 = b_3 = 0.3$. Increasing the transmissibility of non-seeded allele 1 aids the complement 13 of seeded strain 02 and therefore of seeded strain 02, with little effect on strain 12.*

#### Changing two allele-transmission-rates

What if we increase both $b_1$ and $b_3$? With low transmissibility for both of its alleles, strain 02 finally loses its seed advantage (Figure S4). It has enough strength for a second wave, but soon virtually disappears. As a result, strain 13 loses its complementary susceptibility advantage; although it has the highest total transmissibility, 13 loses out to the complementary strains 12 and 03 in the long run. The adjacency effect helps strains 12 and 03 mount an epidemic wave even before strain 13 does.

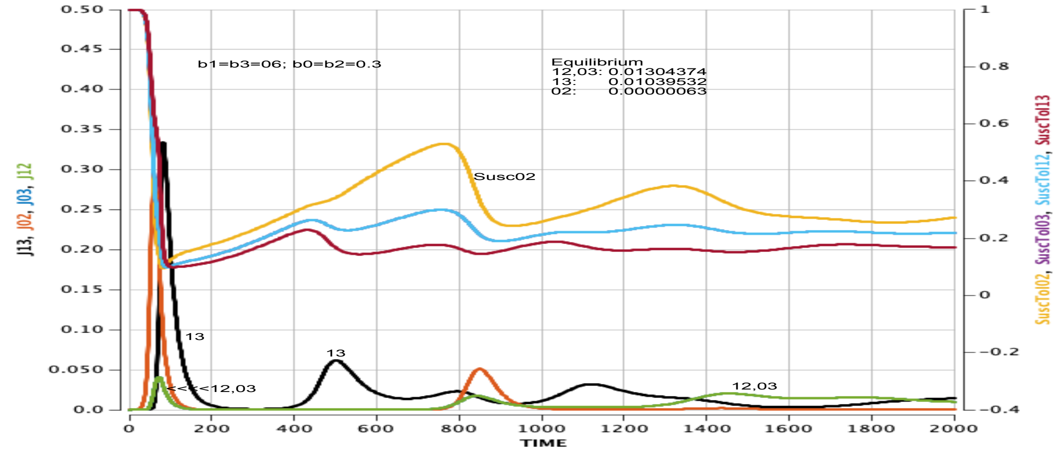

*Figure S4. $b_0 = b_2 = 0.3$, $b_1 = b_3 = 0.6$. Increasing transmissibility of non-seeded alleles 1 and 3 eventually decreases the prevalence of seeded strain 02.*

#### Increase waning rate homogeneously:

In Figure S5, we decrease all waning rates from the basic rate of 0.003 to 0.001. In Figure S6, we increase all waning rates to 0.005. Comparing Figures S5, 3 and S6, we find that faster waning leads to steeper susceptibility curves that dramatically shrink the intervals between pairs of epidemic waves; no other changes from basic model.

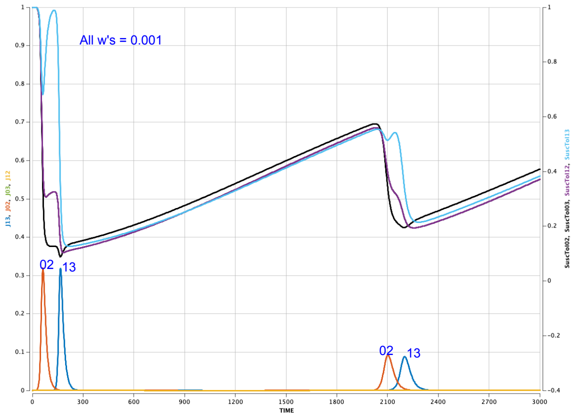

Figure S5. All waning rates at 0.001.

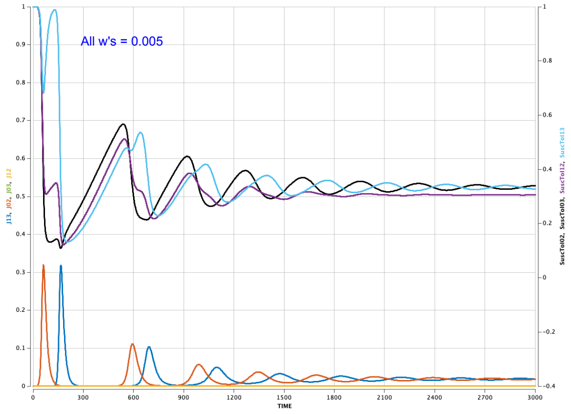

Figure S6. All waning rates at 0.005.

What if we change the waning rate for just one allele, say allele 1? We increased $w_{11}$ and $w_{12}$ from 0.003 to 0.03. Of course, the initial epidemic 02/13 wave pair is not affected by the waning rate. But after these, strains 12 and 13 dominate, as Figure S7 shows. Figure S7 also illustrates the high susceptibility to strain 12 early in the pandemic.

This surge of strains 12 and 13 that results from increased allele-1 waning rates shows the strong “direct parameter influence” of an increased waning rate. As [Sa24a, To24] point out, waning rate makes a big difference.

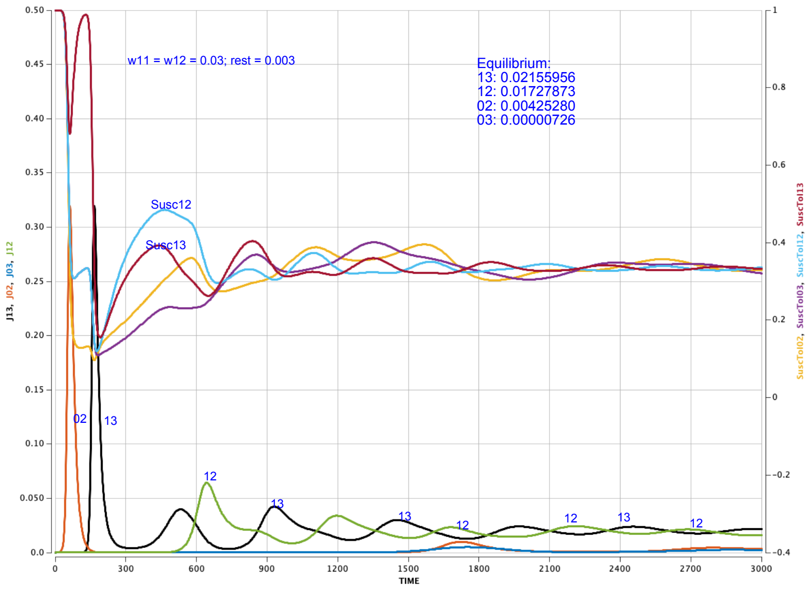

*Figure S7. Increasing the waning rate of allele 1 leads to high prevalences of the strains with allele 1($w_{11}$= $w_{12}$=0.03, other w's = 0.003).*

#### Changing the recovery rate $v_{ij}$, or equivalently the duration of infection $v_{ij}^{-1}$

If we lower the infection recovery rate $v_{ij}$ for all strains from our basic value of 0.1 to, say 0.05 – equivalently doubling the duration of infection for all strains, then the new strains arising via mutation make their appearance, while the pre-mutation strains are still active. The result, as seen in Figure S8, is the disappearance of any epidemic gap, and a four-fold increase in long-run values.

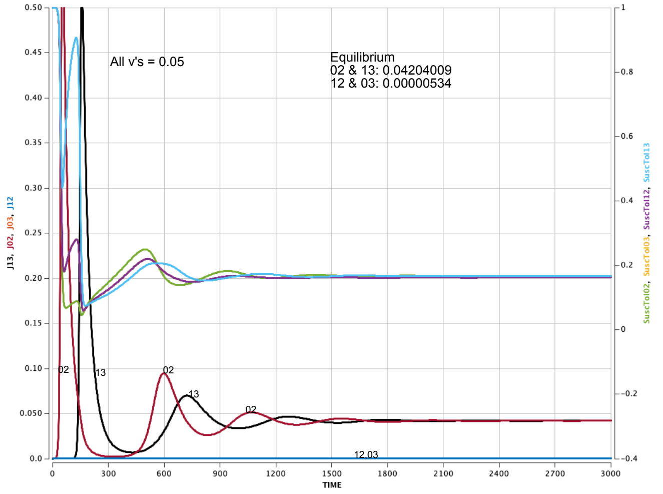

*Figure S8. Slowing all recovery rates from 0.1 to 0.05 strongly increases prevalences.*

Doubling all the recovery rates to 0.2, and thus halving the infection durations, substantially increases the epidemic gaps and leads to a five-fold decrease in long run infection levels.

What if we increase only one duration, say that of strain 03? We change $v_{03}$ from 0.1 to 0.05, keeping the other recovery rates at 0.1. As seen in Figure 4 in the text, strain 03 gains strength impressively. Strain 02 and its complement still start the pandemic, of course. But they quickly lose their advantage as seed and seed complement, and virtually disappear from the graph. Strain 03 with its longer duration dominates ("direct parameter effect") complemented by regular epidemic waves from strain 12 ("complement effect").

Smaller increases in duration of 03 lead to similar, but slightly less dramatic effects. A strain with a longer duration of infection can dominate the pandemic in our model.

**Changing ν, the residual immunity upon total waning**

The parameter $\nu_i \in [0,1]$ represents the relative immunity for allele i at the end of the waning process: $\nu_i = 0$ when there is no immunity to allele i at the end of the waning process. The smaller $\nu_i$ , the lower the overall immunity to infections by strains with allele i, the higher the overall susceptibility to such strains, the higher the transmissibility rate to infection by such strains. We illustrate this by analyzing prevalences for all the $\nu_i$'s = 0, 0.2, and 0.4, with the rest of the parameters at their basic levels.

| ν | 1st 02 epidemic | 1st 13 epidemic | 2nd 02 epidemic | 2nd 13 epidemic |
|---|---|---|---|---|
| 0 | 0.3194 at t=62 | 0.3180 at t =166 | 0.1078 at t=854 | 0.1020 at t=951 |
| 0.2 | 0.3194 at t=62 | 0.3181 at t =166 | 0.1000 at t=1020 | 0.0935 at t=1118 |
| 0.4 | 0.3194 at t=62 | 0.3181 at t =166 | 0.0771 at t=1328 | 0.0697 at t=1426 |

| ν | LR 02/13 equilibrium | LR 12/03 equilibrium |
|---|---|---|
| 0 | 0.01216871 | 0.00000110 |
| 0.2 | 0.01019255 | 0.0000011 |
| 0.4 | 0.00731364 | 0.00000118 |

Of course, increasing immunity parameter $\nu_i$ has no effect on the size or timing of the first 02 and 13 epidemics. The more immunity in the population at the end of waning, the fewer new infections and the longer it takes to reach the susceptibility curve thresholds that signal the start of a new epidemic wave. Fewer infections also implies smaller succeeding epidemic waves and lower equilibrium infection levels.

This higher residual immunity implies fewer infections among the totally waned, i.e., those in compartment R0000. Figure S9 illustrates the larger R0000 compartments with increasing ν.

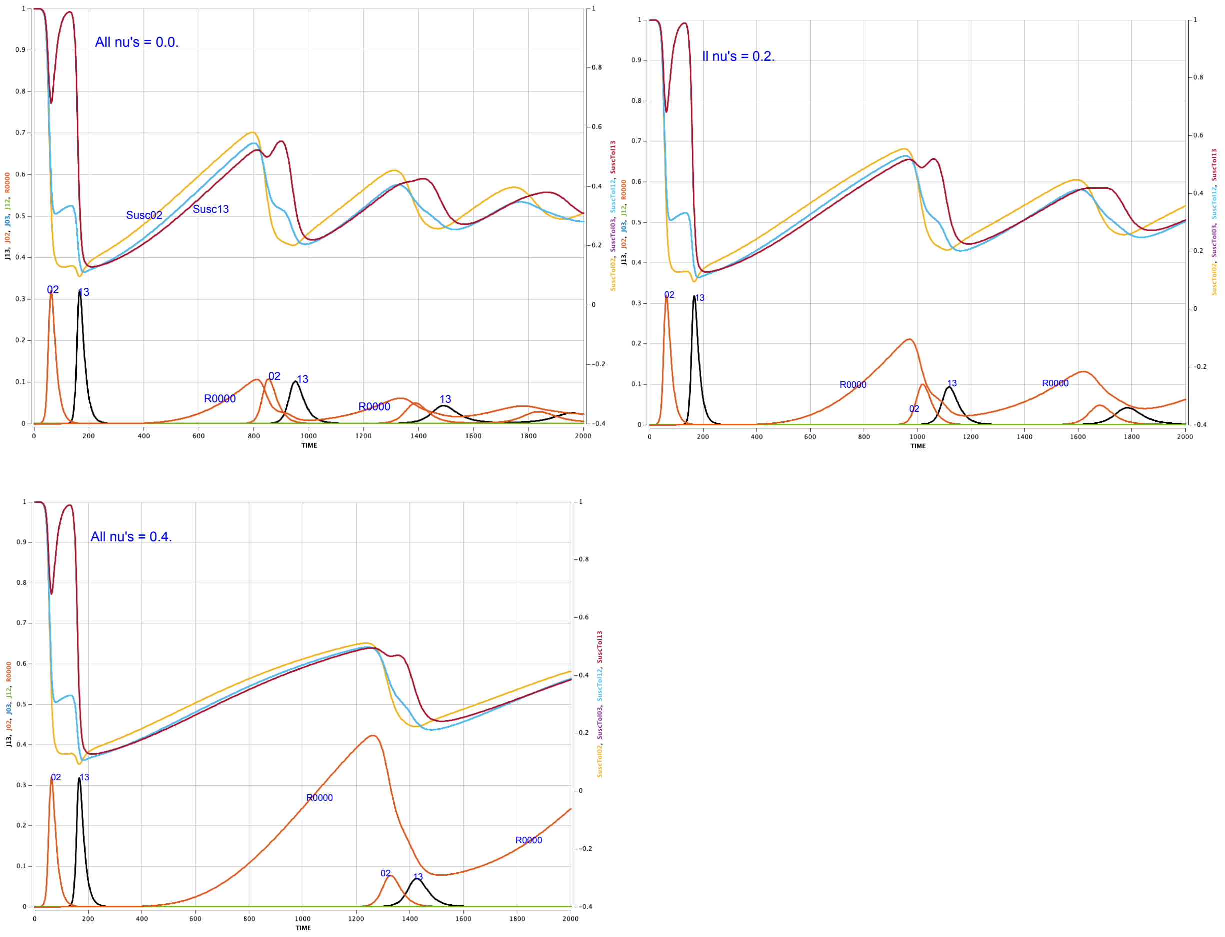

Figure S9. Prevalence and Susceptibility curves for three values of ν: ν = 0.0 (top left), ν = 0.2 (top right) and ν = 0.4 (bottom).

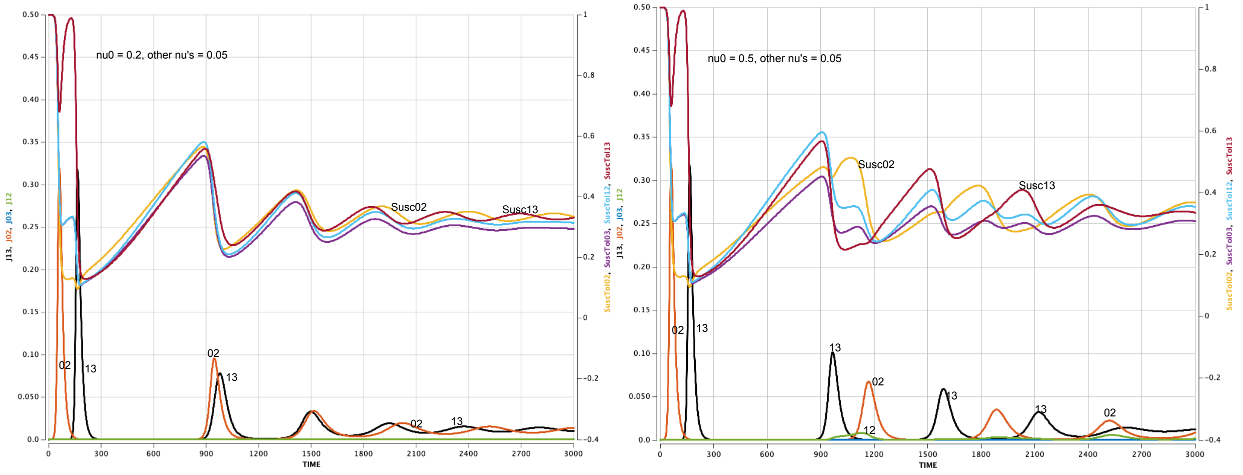

Figure S10. Focusing on allele 0: as $\nu_0$ increases from 0.05 (Figure 3) to 0.2 (left) to 0.5 (right), strain 02 epidemic waves experience lower peaks and longer inter-epidemic gaps.

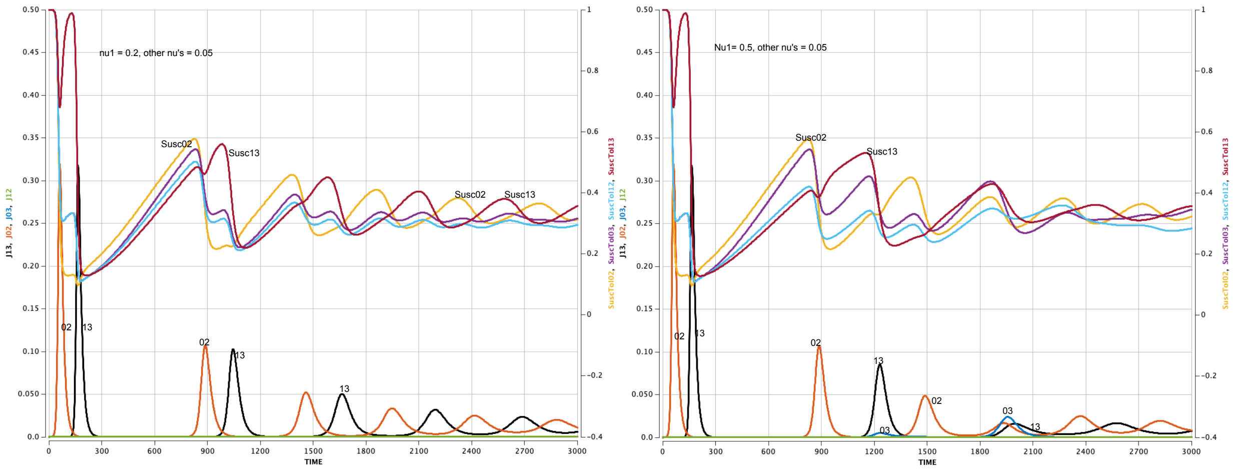

Figure S11. As $\nu_1$ increases from 0.05 (Figure 3) to 0.2 (left) to 0.5 (right), strain 13 epidemic waves experience lower peaks and longer inter-epidemic gaps.

#### Changing relative immunity $m_i$ homogeneously

The parameter $m_i$ represents a susceptible's relative maximal immunity to allele i infections. In Figure S12, we present the graphs for the four strains and their susceptibility curves for all $m_i$ = 0.5, 0.7, and 0.9 respectively. The values of the other parameters are the same as in Figure 3.

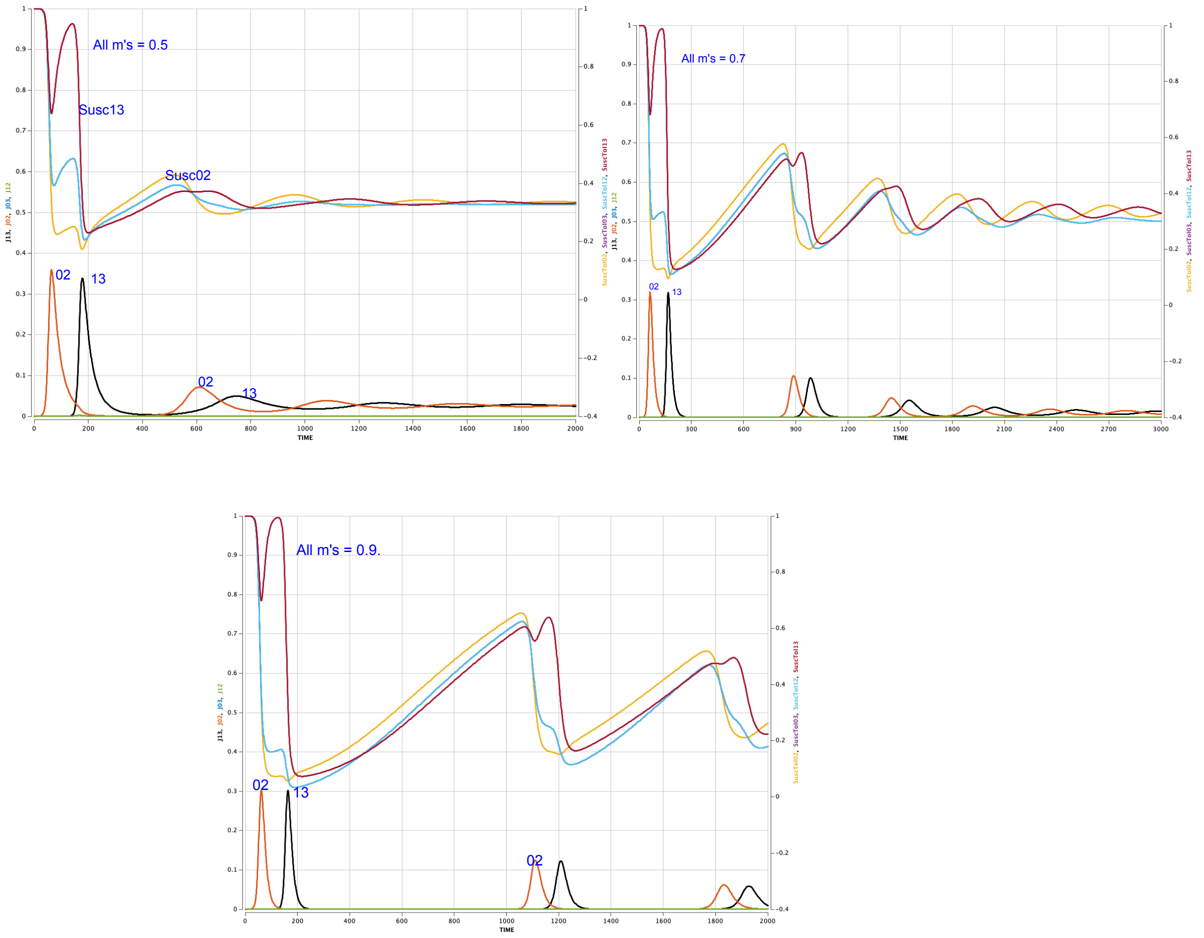

Figure S12. Graphs for 02, 13, Susc02, Susc12=Susc03, and Susc13 for all $m_i$'s = 0.5 (top left), for all $m_i$'s = 0.7 (top right), and for all $m_i$'s = 0.9 (bottom).

In Figure S13, we illustrate the dramatic changes in Susc02 and in the size of compartments R0000 and R3333 for the changes in all the immunities $m_i$ diagrammed in Figure S12.

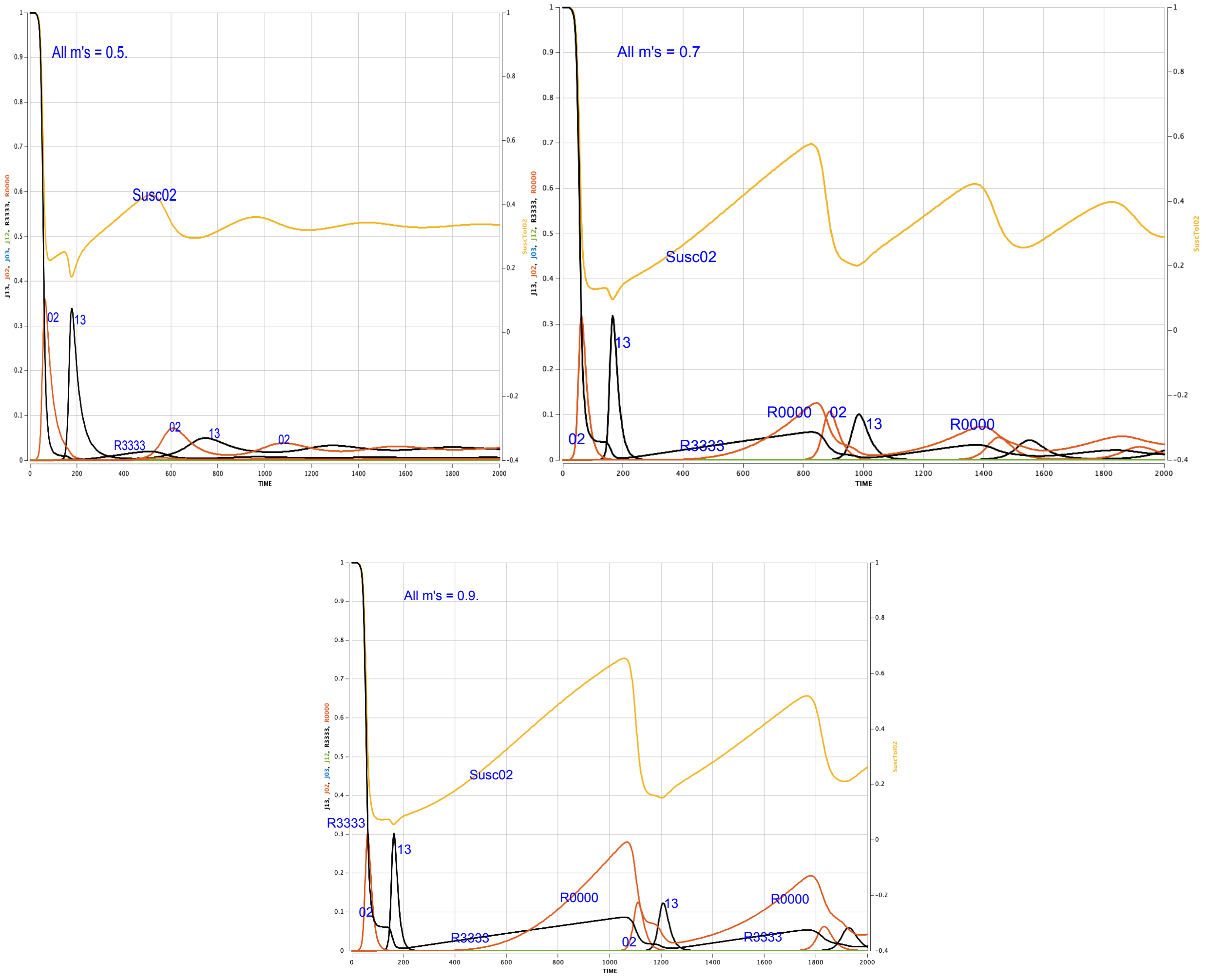

Figure S13. Epidemic curves for strains 02 and 13, and graphs of R3333, R0000 and Susc02 for all $m_i$'s = 0.5 (top left), for all $m_i$'s = 0.7 (top right), and for all $m_i$'s = 0.9 (bottom).

#### Changing relative immunity $m_i$ non-homogeneously

We now allow for variation among the alleles' immune responses, allowing different $m_i$ for different alleles i. We first examine what happens to prevalence patterns as we drop one $m_i$ from 0.7 to 0.5 or even 0.3.

We start by dropping $m_0$ to 0.5. This lower immunity should aid strains 02 and 03. As Figure 8 shows, it does –- to an extent. Epidemic waves of 02 remain at roughly the same height, but the gaps between them are shorter. Strain 03 is a bit stronger and strain 13 a bit weaker; but its role as complement to strain 02 allows strain 13 to soon dominate strain 03, despite 03's immunity advantage.

Figure 8 in the text show what occurs when we drop only $m_0$ from 0.7 to 0.5 and then 0.3. The following tables provide more detail.

**Summary for $m_0$ = 0.5, other $m_i$s = 0.7 (Figure 8)**

| Strain | Prevalences | Equilibrium |
|---|---|---|
| 02 | Waves of same height, but shorter in-between gaps. “dominant.” | 0.01450878 |
| 03 | increased prevalence, including epidemic wave after second 02 wave | 0.01019182 |
| 13 | still strong but lower epidemic peaks and longer gaps | 0.00309363 |
| 12 | miniscule prevalence | 0.00000790 |

**Summary for $m_0$ = 0.3, other $m_i$s = 0.7 .**

| Strain | Prevalences | Equilibrium |
|---|---|---|
| 02 | Similar wave patterns, shorter in-between gaps. “dominant.” | 0.01911205 |
| 03 | biggest gainer, beats out 13 for the second wave | 0.01279877 |
| 13 | much weaker, lower epidemic peaks and longer gaps | 0.00600359 |
| 12 | miniscule prevalence | 0.00000385 |

The same graphs and tables hold for the case where $m_2$ is the uniquely lower $m_i$, if one interchanges 12 and 03 in both.

Lowering the immunity $m_0$ to allele 0 even more, say to 0.3, allows strain 03 to overcome this strain 13 complementarity advantage (Figure S14). In the long run, strains 02 and 03 have the highest prevalences. And they do not need strong complements to flourish.

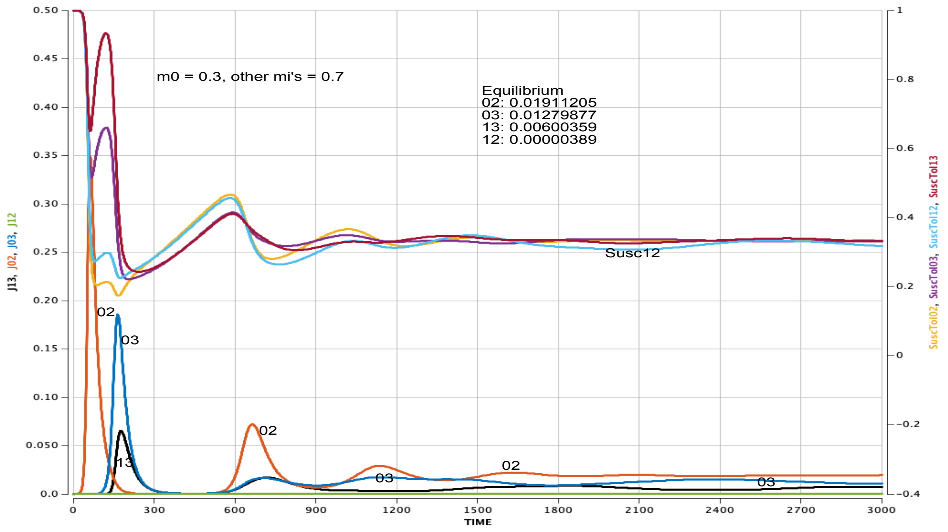

*Figure S14. If we lower immunity $m_0$ to allele 0 a lot, strains 02 and 03 soon dominate ($m_0$ = 0.3, other $m_i$’s = 0.7).*

Figure S15a and the table below it present the analogous patterns when $m_1$ for allele 1 is lowered from 0.7 to 0.5.

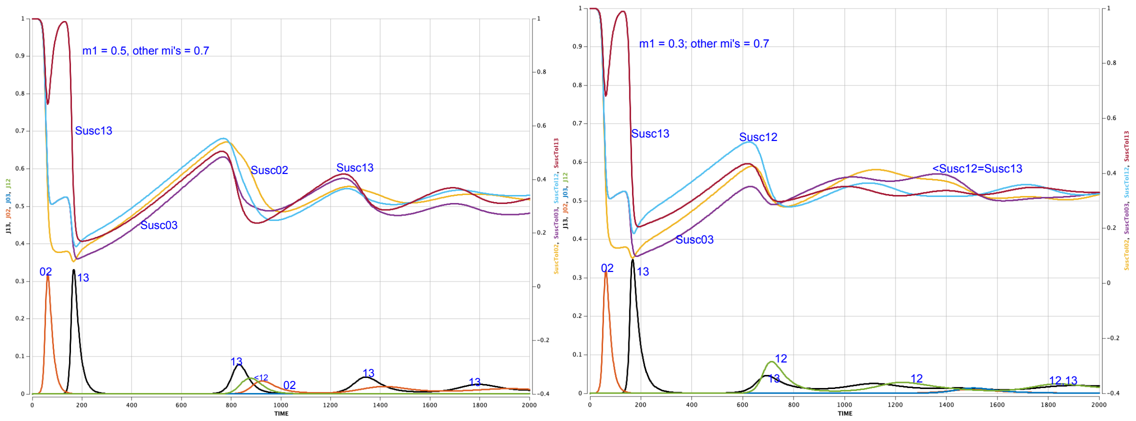

*Figure S15. Prevalences and susceptibilities for $m_1$ = 0.5 (left) and $m_1$ = 0.3 (right), other $m_i$'s = 0.7.*

**Summary for $m_1$ = 0.5, other $m_i$s = 0.7 (Figure S15a).**

| Strain | Prevalences | Equilibrium |
|---|---|---|
| 13 | Waves of same height, but shorter in-between gaps; "dominant." | 0.01450878 |
| 12 | increased prevalence, including epidemic wave after second 02 wave | .01019182 |
| 02 | still strong but lower epidemic peaks and longer gaps | 0.00309363 |
| 03 | miniscule prevalence | 0.00000790 |

If $m_3$ has the lower $m_i$ = 0.5, the patterns is the same as in Figure S14 and the table above, again interchanging 03 and 12.

Figure S15b and the Table below present the patterns when $m_1$ is further decreased to 0.3. Now, seeded strain 02 more or less disappears after the first wave. With the smaller $m_1$, strain 12 is the biggest winner in terms of prevalences.

**Summary for $m_1$ = 0.3, other $m_i$s = 0.7 (Figure S15b).**

| Strain | Prevalences | Equilibrium |
|---|---|---|
| 12 | biggest winner | 0.01911205 |
| 13 | somewhat similar waves, but shorter in-between gaps. | 0.01279877 |
| 02 | "disappears" after its initial wave | 0.00600359 |
| 03 | miniscule prevalence | 0.00000389 |

**Change two $m_i$s.**

Figure S16a shows the patterns when we decrease two alleles -- both at the **same site**. At equilibrium, strains 02 and 13 are at 0.02060084, while strains 12 and 03 are at 0.00000475, in both panels.

Figure S16b shows the patterns when we decrease two allele-immunities -- both at different sites (different right axis scale). In this case, the equilibria are:

| | strain 02 | strain 13 | strains 12 and 03 |
|---|---|---|---|
| | 0.005504 | 0.31599703 | 0.00003449 |
| | 0.31599703 | 0.005504 | 0.00003449 |

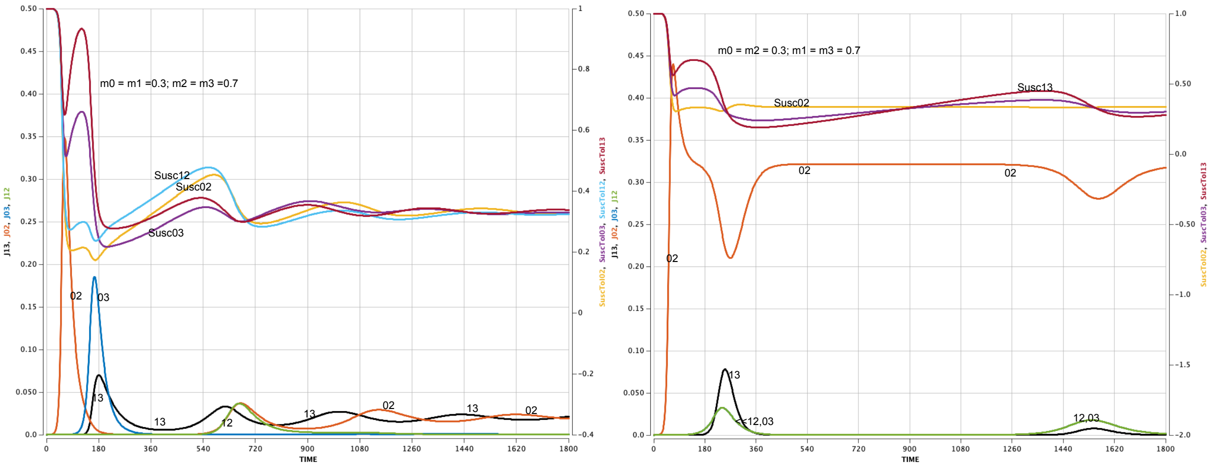

*Figure S16. Prevalences and susceptibilities for $m_0 = m_1 = 0.3$, $m_2 = m_3 = 0.7$ (left panel) and $m_0 = m_2 = 0.3$, $m_1 = m_3 = 0.7$ (right panel).*

Notice the huge difference in total prevalence at equilibrium:

- ~ 4% in Figure S16
- ~ 32% in Figure S17.

In the case of Figure S16 no strain gains an advantage from the reduced immune response. In the case of Figure S16b, a single strain gains a very large advantage.

The Figures also point to the power of the seeded strain and its complement. Even though reduced immunities affect all strains the same by the numbers, strains 02 and/or 13 dominate by far in both figures.

**No immune pressure**

Finally, we look at the case where there is little or no immunity to the seeded strain 02. We set $m_0$ and $m_2 = 0$. The result has shown in Figure S17: no successful mutation. As [Sa22] puts it: “if there is no immune pressure, then viral abundance may be high but selection for immune escape is absent.”

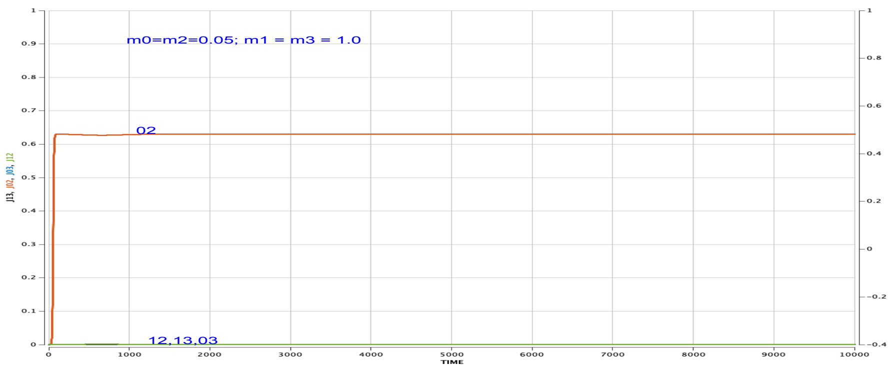

*Figure S17. If there is no immunity for the seeded alleles ($m_0 = m_2 = 0$), seeded strain 02 quickly becomes the only positive strain in the pandemic.*

#### Adding joint immunity

If the $m_i$’s are reasonably large, adding some joint immunity effects has little effect. The Table below shows the values of peaks of the first epidemic and the long run equilibria for all $m_i$=0.7 and all $m_{ij}$s =0, 0.2, and 0.4. Figure S18 shows the graphs of all the strain prevalences and susceptibility curves for all $m_i$’s = 0.7 and both for all joint $m_{ij}$’s = 0 and 0.2. Note the very small differences. For $m_i$’s = 0.9, there is virtually no change as one increases the values of the joint immunities (not shown).

**Table. All $m_i$=0.7: very small changes as $m_{ij}$ increases**

**All $m_i$ =0.7 , all $m_{ij}$ = 0**

| strain | first peak | long run equilibrium |
|---|---|---|
| 02 | 0.3197 at t=62 | 0.01173624 |
| 13 | 0.3180 at t=166 | ‘’ |
| 12&03 | 0.00007 at t=70 | 0.00000109 |

**All $m_i$=0.7 , all $m_{ij}$ = = 0.2**

| strain | first peak | long run equilibrium |
| --- | --- | --- |
| 02 | 0.3154 at t=62 | 0.01124877 |
| 13 | 0.3140 at t=169 | “ |
| 12&03 | 0.00007 at t=70 | 0.00000084 |

**All $m_i$=0.7 , all $m_{ij}$ = = 0.4**

| strain | peak | long run equilibrium |
| --- | --- | --- |
| 02 | 0.3115 at t=62 | 0.0108444 |
| 13 | 0.3104 at t=165 | “ |
| 12&03 | 0.00007 at t=71 | 0.00000105 |

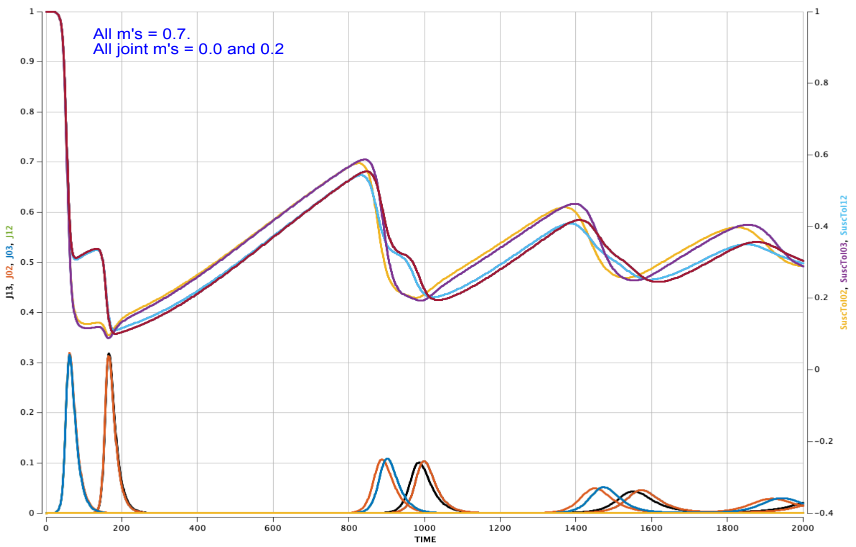

*Figure S18. Graphs of all four strains for all $m_i$'s = 0.7 and all joint $m_{ij}$'s = 0, overlaid with same for all $m_i$'s = 0.7 and all joint $m_{ij}$'s = 0.2.*

#### Changing the initial seed

We examine the effects of varying these initial conditions, always keeping the initial size of a strain at 0.00001.

**Initial seeds with complementary pairs, either 12 and 03 or 13 and 02**

If the initial seeds are a complementary pair, these two strains dominate and, their epidemic waves overlap each other, as seen in Figure S19.

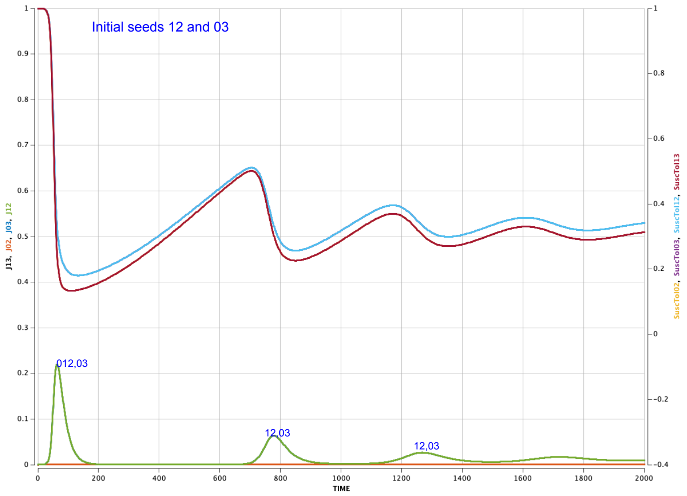

*Figure S19. Prevalences and susceptibility curves for initial seeds 13 and 02.*

**Initial seeds 12 and 13 or 02 and 03.**

The scenario where the two initial seeds are not complementary strains is more complex (and interesting). When we started with 12 and 13 as seeds, one of us found $E_{02}$* as the long run equilibrium, the other found $E_{03}$*. In fact, the initial conditions:

(*) I1233 and I1333 set at 0.00001, I0233 and I0333 set at 0, and complete susceptibles R3333 set at 0.99998, with all other variables =0,

are on the boundary that separates the two basins of attraction. To see this, we ran a series of ten runs, all with the initial I1333(0) set at 0.00001 but with the initial I1233(0) taking on values between 0.000005 (just below 0.00001) and 0.000015 (just above 0.00001).

When I1233(0) < 0.00001 = I1333, the system converged to $E_{02}$*.

When I1233(0) > 0.00001 = I1333, the system converged to $E_{03}$*.

**Three initial seeds**

Next, we start with initial conditions:

I1233 = 0.00001, I1333 = 0.00001, I0233=0.00001, R3333 = 0.99997, all other variables =0 (including I0333).

Now, strains 12, 13, and 02 are seeded, but not strain 03.

Using the parameters in Section 3, we illustrate the prevalences and susceptibility curves in Figure S20. Because of the symmetries in their coefficients and starting values, the prevalence curves for strain 02 exactly overlap those for strain 13, as do the sizes of their recent recovery compartments R2323 and R3232.

We include in Figure S20 the sizes of the main recovery compartments, R3333, R2323, R3232, and R3223, parametrized on the (raised) *right* vertical axis to minimize overlap in the figure.

- Not surprisingly, there is a spike epidemic right away for all three seeded strains in Figure S20. During this spike:
    - The 02 infecteds recover into compartment R2323, where they are highly susceptible to strain 13.
    - The 13 infecteds recover into compartment R3232, where they are highly susceptible to strain 02.
    - With this feedback process, the 02 and 13 epidemic waves achieve a higher peak than does the12 epidemic wave.
- By t=126, 99% of the population has been infected, as seen by compartment of never-infected susceptibles, R3333, at level 0.0066 at t=126.
- After this triple epidemic, nearly all the susceptibles are in compartments R2323, R3232, and especially R3223 (those just recovering from strain 12).
- Spurred by the size of R3223 and the addition of new entrants into R3333, Susc03 reaches a threshold that leads to a strain 03 epidemic wave – even though 03 was the only unseeded strain.
- This 03 wave is naturally followed by a complementary strain 12 wave,
- Meanwhile, those who had recovered from the 02 and 13 epidemics experience waning immunity so that Susc02 and Susc13 grow and reach their thresholds,
    - leading to further 02 and 13 epidemics, and
- eventual dominance at the long run equilibrium $E_{02}$*,

- where strains 02 and 13 equilibrate at level 0.0117624,
- while strains 12 and 03 equilibrate at 0.00000109.

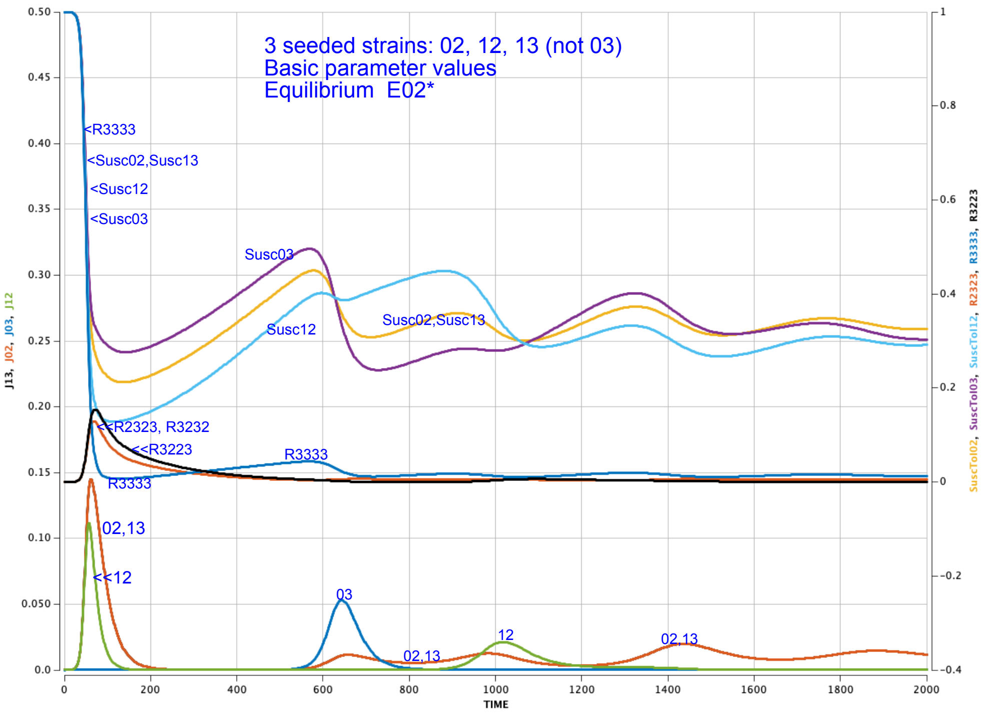

*S20. Prevalence and susceptibility curves (and the relevant R- compartments) when all but strain 03 are initially seeded.*

This is an interesting example because there are epidemic waves for all four strains, even though 02 and 13 dominated in the beginning and at the end. The waning of R3232 and R2323 lead to resurgences of the Susc02 and Susc13 curves.

#### Changing Mutation Rates $f_{ij}$

Next, we examine the effects of changing the mutation rates $f_{ij}$. Figure S21 illustrates the dynamics for uniform mutation rates of 0.000000001, 0.00001 (basic model), 0.0001, and 0.01. Not much happens. As the mutation rate rises, the epidemic peaks fall, and the inter-wave gaps decrease; eventually strains 12 and 03 appear for a while.

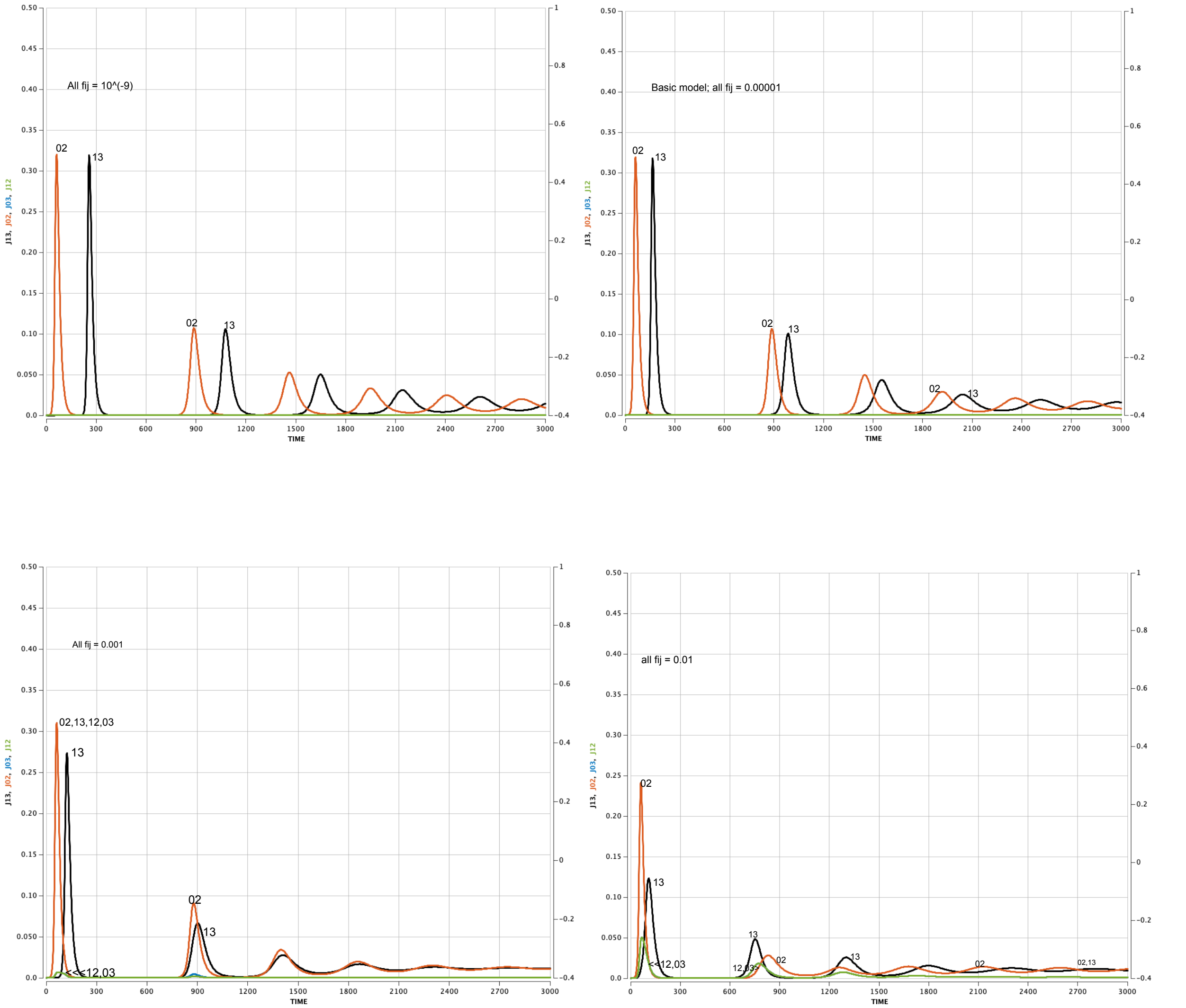

Figure S21. Uniform mutation rates $f_{ij}$ = 0.000000001 (top left), 0.00001 (basic model, top right), 0.001 (bottom left), 0.01 (bottom right)

At the extreme of all four $f_{ij}$ = 0.1, the four epidemic curves are more or less identical (Figure S22).

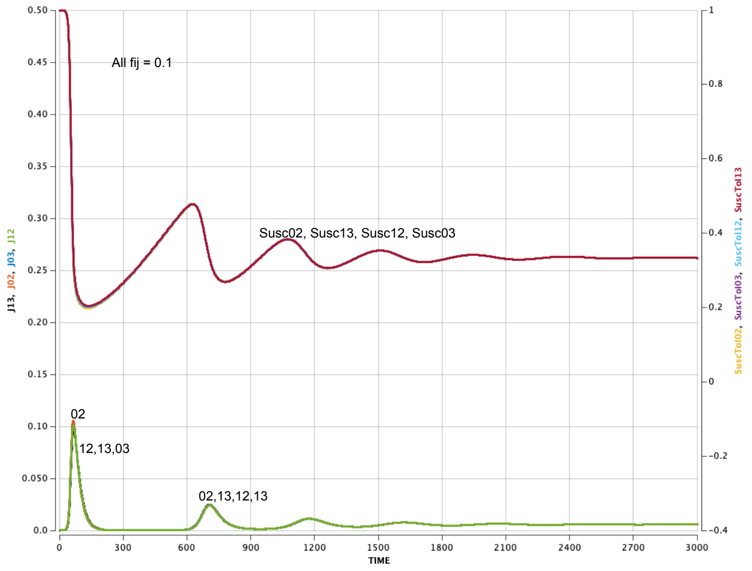

*Figure S22. All mutation rates $f_{ij}$ = 0.1.*

Figure 10 in the text illustrates the case where on mutation rate, say $f_{01}$ is much faster than all the others. Strain 12 and its complement 03 dominate the initial epidemic waves, but founding strain and its complement dominate thereafter.

**Changing the ADE parameter d**

Antibody-Dependent Enhancement (ADE) occurs when pre-existing non-neutralizing antibodies, which had arisen in response to previous infection, lead to enhanced new infection by facilitating the virus's cell entry.

To model ADE, we assign a number $d_i$ to each allele i that reflects how the immune response to that allele can increase transmissibility via ADE. We include $d_0 + d_1 + d_2 + d_3$ in the transmission term, but we set $d_i$ to zero if there has never been an immune response to allele i for those in that compartment is at level 3.

In the subfigures in Figure S23 we maintain the four $b_i$s at 0.2 and the $b_{ij}$s at 0.
We increase the ADE factor $d_i$ from 0 to 0.03 to 0.07 to 1.0. To compare curve heights better, we increased the scale of the left vertical axis. Figure S24 shows that the same story holds for even all $b_i$s = 0.3.

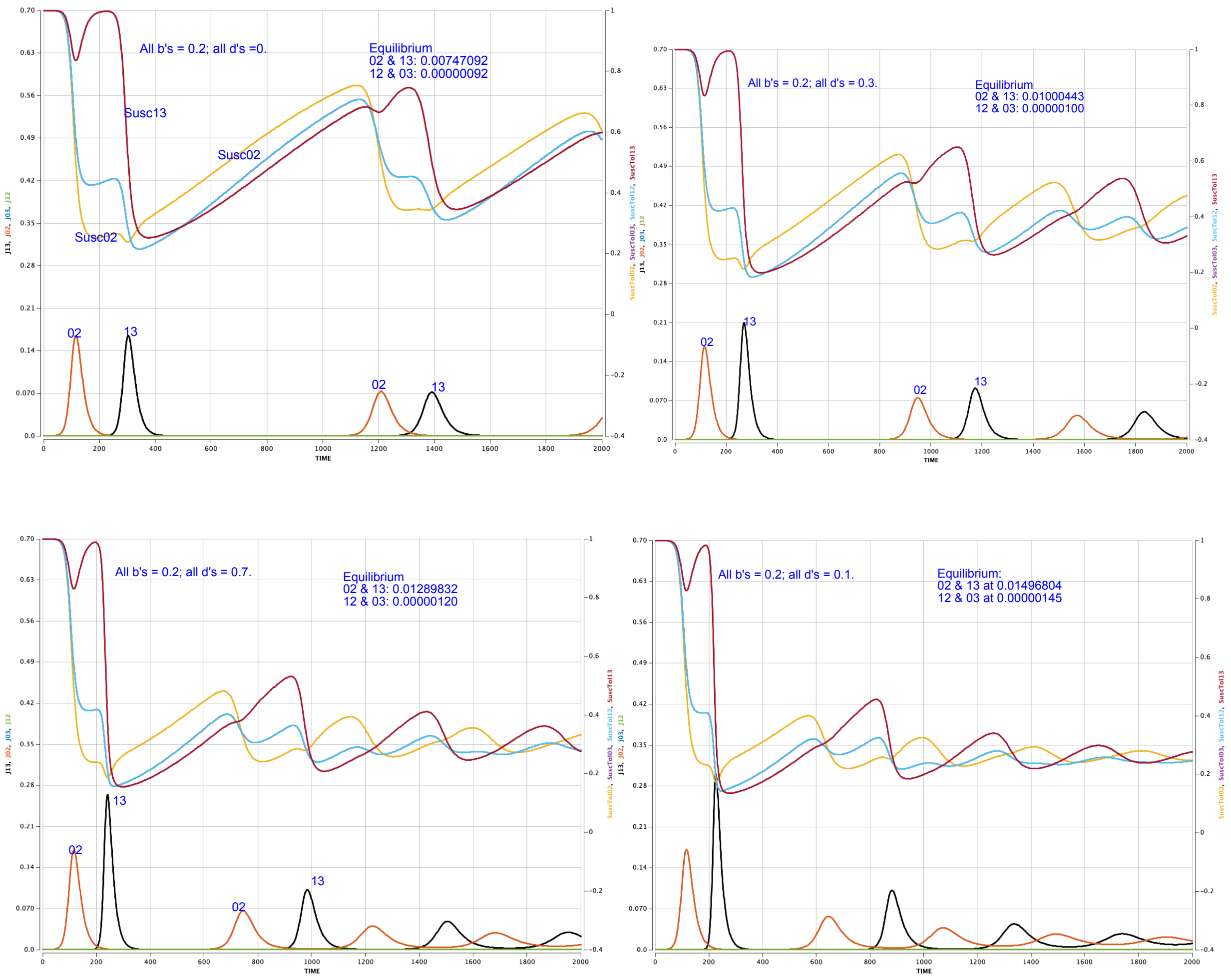

*Figure S23. Prevalence and Susceptibility curves for all $b_i$=0.2 and with ADE factor $d_i$ = 0 (top left), 0.3 (top right), 0.7 (bottom left) and 1.0 (bottom right).*

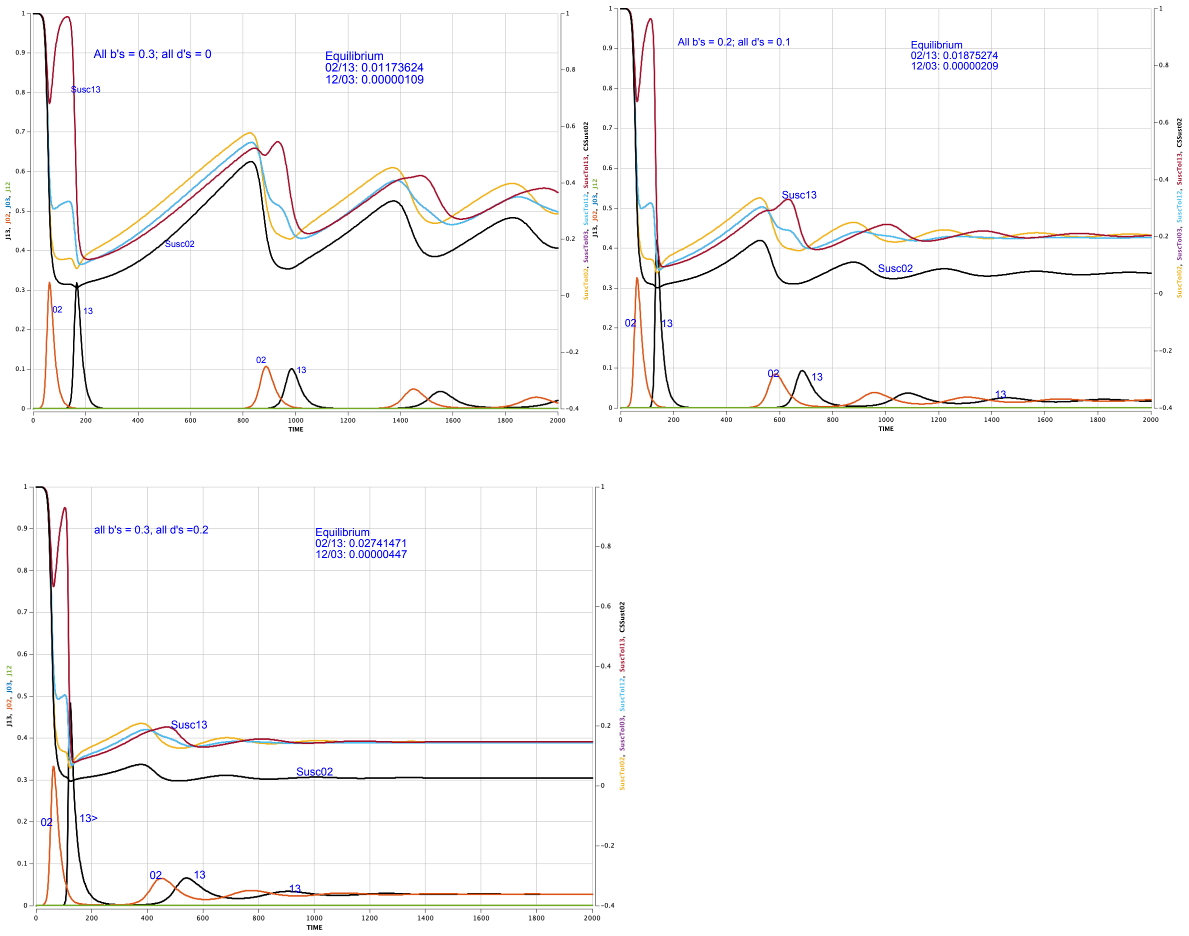

Figure S24. Prevalences and susceptibility curves for all $b_i$s = 0.3 and all $d_j$s = 0 (top left), 0.1 (top right), and 0.2 (bottom).

The ADE parameters play no role at the very beginning of the pandemic -- in a population of never-infected susceptibles. As a result, the inclusion of ADE terms has little effect on the first strain 02 epidemic wave, but it does increase the levels of later 13 epidemic waves. With increasing $d_i$, not only does the peak of the first 13 wave increase, but so does the peak of the second and third 13 waves. Higher levels of infections in the gaps means that the Susc13 curves reach their threshold sooner, and the gaps become shorter. The peaks of the second and third 02 waves decrease with increasing $d_i$. The gaps between subsequent 02 waves decrease, but not as fast as the gaps between subsequent 13 waves.

All this leads to the rare situation in which the height of the initial 02 epidemic is smaller than the height of the succeeding 13 epidemic.

It is natural to ask what happens if we decrease the $b_i$s even further, say to 0.1, while keeping the $d_i$s high. Figure S25 shows the answer: basically nothing happens, at least for the first 6,000 time-steps. The higher values of the $b_i$s are needed to achieve $R_0 > 1$ so the epidemic can get started.

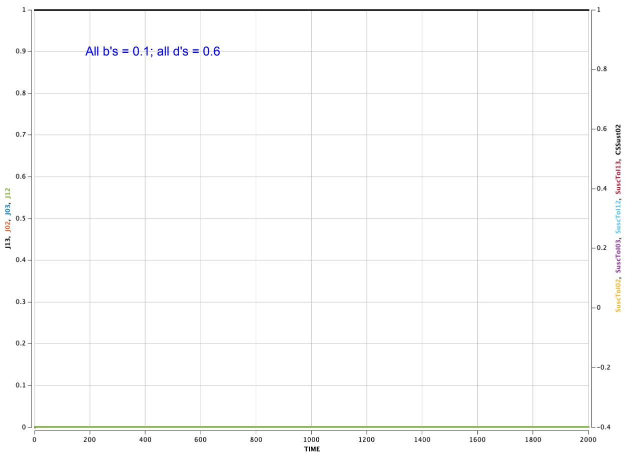

*Figure S25. Prevalences when all $b_i$s = 0.1 and all $d_j$s = 0.6.*

### VI. Future Work:

One pressing next step is to increase the number of sites in our model. But even small increases lead to unwieldy numbers. Consideration of just ten sites with two alleles at each site yields a system with 20 alleles and 1,024 strains. Keeping track of immune history, say with five levels of waning, leads to roughly $10^{14}$ compartments of susceptibles. It may be more reasonable to aim for an in-between goal of four sites with four immunity levels; that would yield about $4^8$ = 65,536 compartments of susceptibles.

This, our first allele-based model, focused on biological processes and avoided sociological concerns since we are especially interested in relating our model to the work of laboratory scientists. Future projects will incorporate population effects, such as age structure, and heterogeneous contact rates and immune responses.

We are especially eager to elaborate the processes in the model to integrate with knowledge generated by laboratory studies, including ultimately a better understanding of how the RNA dependent RNA polymerase (RdRp) of coronaviruses influences coronavirus evolution. Such understanding might link RdRp structure to rates of genetic drifting that affect virus evolution processes.

Three-quarters of the SARS-CoV-2 genome encodes how genetic sequences are translated into specific sequence codes in the virus. The size and function of this translation site are specific to coronaviruses. This is the region in which the enzyme RdRp plays a key role. Our allele-based model does not formulate any effects of RdRp, but it can help discover such effects and indicate how large they may be by formulating processes at the level of allele specific effects, a much more difficult task for the coarser strain-based approach.

### VII. Supplementary Material References

[Dys21] L. Dyson, E.M. Hill, S. Moore, et al. Possible future waves of SARS-CoV-2 infection generated by variants of concern with a range of characteristics. Nature Communications 12:5730. 2021.

[F18] Fisher, R.A. The correlation between relatives on the supposition of Mendelian inheritance. *Trans. Roy. Soc. Edinburgh*. **52**,399–433, 1918.

[H08] Hardy, G. H. "Mendelian Proportions in a Mixed Population." *Science.* ***28*** *(706): 49–50.* (July 1908).

[JS93] JA Jacquez, CP Simon. Qualitative Theory of Compartmental Systems. S.I.A.M. Review, 35 (1993) 43—79, 1993.

[Me23] Meijers et al. Population immunity predicts evolutionary trajectories of SARS-CoV-2. Cell. 2023. doi:10.1016/j.cell. 2023.09.022.

[Sa20] Chadi M. Saad-Roy1, Caroline E. Wagner, Rachel E. Baker, Sinead E. Morris, Jeremy Farrar6, Andrea L. Graham, Simon A. Levin, Michael J. Mina, C. Jessica E. Metcalf, Bryan T. Grenfell. Immune life history, vaccination, and the dynamics of SARS-CoV-2 over the next 5 years. *Science* 370, 811–818 (2020).

[Sa21a] Saad-Roy CM, Levin SA, Metcalf CJE, Grenfell BT. Trajectory of individual immunity and vaccination required for SARS-CoV-2 community immunity: a conceptual investigation. *J. R. Soc. Interface* 18: 20200683. 2021.

[Sa21b] C.M. Saad-Roy, S.E. Morris, C.J.E. Metcalf, et al. Epidemiological and evolutionary considerations of SARS-CoV-2 vaccine dosing regimes. *Science* 372, eabg8663. 2021.

[Sa22] Chadi M Saad-Roy, Jessica E. Metcalf, Bryan T. Grenfell. 2022. "Immuno-epidemiology and the predictability of viral evolution." *Science* 376, 1161-1162.

[Sa23] C.M. Saad-Roy, S.E. Morris, R.E. Baker, et al. Medium-term scenarios of COVID-19 as a function of immune uncertainties and chronic disease. *Royal Society Interface* 20:20230247. 2023.

[Sa24a] Chadi M. Saad-Roy, Sinead E. Morris, Mike Boots, Rachel E. Baker, Bryan L. Lewis, Jeremy Farrar, Madhav V. Marathe6,8, Andrea L. Graham, Simon A. Levin, Caroline E. Wagner, C. Jessica E. Metcalf, Bryan T. Grenfell. 2024. Impact of waning immunity against SARS-CoV-2 severity exacerbated by vaccine hesitancy. *PLOS Computational Biology* | https://doi.org/10.1371/journal.pcbi.1012211. August 5, 2024 1 / 18

[To24]  X. Tong, B. Kellman, M.-J. Avendano, et al. Humoral waning kinetics against SARS-CoV-2 is dictated by disease severity and vaccine platform. medRxiv preprint. 2024. doi:10.1101/2024.10.17.24315607.

[W08] Weinberg, W. "Über den Nachweis der Vererbung beim Menschen". *Jahreshefte des Vereins für vaterländische Naturkunde in Württemberg*. **64**: 368–382. (1908).

[Wa21] C.E. Wagner, C.M. Saad-Roy, S.E. Morris, et al. Vaccine nationalism and the dynamics and control of SARS-CoV-2. *Science* 373(6562): eabj7364. 2021.

[Xi26] Xiong W, Huang X, Thao T et al. Measuring population immunity against influenza using individual antibody titres: a multicountry, retrospective observational study. *The Lancet Infectious Diseases,* 2026; 0.

Python File

```
#!/usr/bin/env python3

# -*- coding: utf-8 -*-

"""

Created on Thu Apr 30 11:18:19 2026
```

```
@author: cpsimon
"""

#!/usr/bin/env python3
"""
Allele-based 2-site, 2-allele SARS-CoV-2 model (submission, ADE wired)
-------------------------------------------------
- Preserves the v2 CLI and styling
- Adds --report-peaks to report the peak (max) value and its time for each strain total:
  J(0,2), J(1,2), J(0,3), J(1,3)
"""

from dataclasses import dataclass
from typing import Dict, Tuple
import numpy as np
from scipy.integrate import solve_ivp
import matplotlib.pyplot as plt
import argparse
from pathlib import Path

# By default, saved plots go in the same folder as this Python file.
DEFAULT_OUTPUT_DIR = Path(__file__).resolve().parent
```

```
# ----------------------------
# Parameter container
# ----------------------------
@dataclass
class Params:
    beta: Dict[int, float]
    e: Dict[Tuple[int,int], float]
    v: Dict[Tuple[int,int], float]
    m: Dict[int, float]
    m_joint: Dict[Tuple[int,int], float]
    nu: Dict[int, float]
    f: Dict[Tuple[int,int], float]
    w2: Dict[int, float]
    w1: Dict[int, float]
    d: Dict[int, float]
    c: float
    mu: float
    tmax: float = 3000.0
    rtol: float = 1e-7
    atol: float = 1e-9
    t_points: int = 1201

def default_params() -> Params:
```

```
    return Params(
        beta={0:0.3,1:0.3,2:0.3,3:0.3},
        e={(0,2):0.0,(1,2):0.0,(0,3):0.0,(1,3):0.0},
        v={(0,2):0.1,(1,2):0.1,(0,3):0.1,(1,3):0.1},
        m={0:0.7,1:0.7,2:0.7,3:0.7},
        m_joint={(0,2):0.0,(1,2):0.0,(0,3):0.0,(1,3):0.0},
        nu={0:0.05,1:0.05,2:0.05,3:0.05},
        f={(0,1):1e-5,(1,0):1e-5,(2,3):1e-5,(3,2):1e-5},
        w2={0:0.003,1:0.003,2:0.003,3:0.003},
        w1={0:0.003,1:0.003,2:0.003,3:0.003},
        d={0:0.0,1:0.0,2:0.0,3:0.0},
        c=0.5,
        mu=0.0001,
    )

# ----------------------------
# Model construction helpers
# ----------------------------
STRAINS = [(0,2),(1,2),(0,3),(1,3)]
KS = [(K0,K1,K2,K3) for K0 in range(4) for K1 in range(4) for K2 in range(4) for K3 in range(4)]
IDX_R = {K:i for i,K in enumerate(KS)}
NR = len(KS)
def idx_I(s_index: int, Hh: int, Hm: int) -> int:
```

```
    return NR + s_index*16 + Hh*4 + Hm
NI = 4*16
NSTATE = NR + NI
S_INDEX = IDX_R[(3,3,3,3)]

def kappa(nu: Dict[int,float], allele: int, K: int) -> float:
    if K == 3: return 0.0
    if K in (1,2): return 1.0
    return nu[allele]
def omega(w2: Dict[int,float], w1: Dict[int,float], allele: int, K: int) -> float:
    if K == 2: return w2[allele]
    if K == 1: return w1[allele]
    return 0.0

def ade_term(P: Params, K: Tuple[int, int, int, int]) -> float:
    """Additive ADE contribution from alleles in the host's immune history.

    K[a] == 3 means no prior immunity/history for allele a.
    K[a] in {0,1,2} means allele a is in the host's immune history,
    so its ADE parameter d[a] contributes to transmission.
    """
    return sum(P.d[a] for a in range(4) if K[a] != 3)
```

```
def precompute_susc_and_Gpref(P: Params):
    susc = {}
    G_pref = {}
    for s_index,(i,j) in enumerate(STRAINS):
        for r_index, K in enumerate(KS):
            kap_i = kappa(P.nu, i, K[i])
            kap_j = kappa(P.nu, j, K[j])
            s = (1.0 - P.m[i]*kap_i) * (1.0 - P.m[j]*kap_j) * (1.0 - P.m_joint[(i,j)]*kap_i*kap_j)
            susc[(s_index, r_index)] = s

            # Transmission factor for strain (i,j) in host immune state K:
            #   [beta_i + beta_j + epistasis_ij + ADE(K)] * susceptibility * contact rate.
            trans = P.beta[i] + P.beta[j] + P.e[(i,j)] + ade_term(P, K)
            G_pref[(s_index, r_index)] = trans * s * P.c
    return susc, G_pref

def build_base_R_for_I():
    """Precompute which R compartments can feed each I compartment.

    For strain (i,j), the inactive alleles are:
      - the alternate allele at site 0: h = 1-i
      - the alternate allele at site 1: m = 5-j
    Hh and Hm record the immunity states for those inactive alleles.
```

```
    """
    base_R_for_I = {}
    for s_index, (i, j) in enumerate(STRAINS):
        h = 1 - i
        m = 5 - j
        for Hh in range(4):
            for Hm in range(4):
                res = []
                for r_index, K in enumerate(KS):
                    if K[h] == Hh and K[m] == Hm:
                        res.append(r_index)
                base_R_for_I[(s_index, Hh, Hm)] = res
    return base_R_for_I

# Initialized after the function definition.
BASE_R_FOR_I = build_base_R_for_I()

def L_of_K(y: np.ndarray, P: Params, K: Tuple[int,int,int,int]) -> float:
    K0,K1,K2,K3 = K
    inc = 0.0
    if K0==2 and K2==2:
        inc += P.v[(0,2)] * y[idx_I(0, K1, K3)]
    if K1==2 and K2==2:
```

```
        inc += P.v[(1,2)] * y[idx_I(1, K0, K3)]
    if K0==2 and K3==2:
        inc += P.v[(0,3)] * y[idx_I(2, K1, K2)]
    if K1==2 and K3==2:
        inc += P.v[(1,3)] * y[idx_I(3, K0, K2)]
    return inc

def psi_factor(P: Params, s_index: int, Hh: int, Hm: int) -> float:
    f = P.f
    if s_index == 0:
        fac = 1.0
        if Hh in (0,3): fac *= (1 - f[(0,1)])
        if Hm in (0,3): fac *= (1 - f[(2,3)])
        return fac
    if s_index == 1:
        fac = 1.0
        if Hh in (0,3): fac *= (1 - f[(1,0)])
        if Hm in (0,3): fac *= (1 - f[(2,3)])
        return fac
    if s_index == 2:
        fac = 1.0
        if Hh in (0,3): fac *= (1 - f[(0,1)])
        if Hm in (0,3): fac *= (1 - f[(2,3)])
```

```
        return fac

    if s_index == 3:

        fac = 1.0

        if Hh in (0,3): fac *= (1 - f[(1,0)])

        if Hm in (0,3): fac *= (1 - f[(3,2)])

        return fac

    return 1.0

def C_inflow(y: np.ndarray, R: np.ndarray, J: np.ndarray, P: Params, G_pref, s_index: int, Hh: int, Hm: int) -> float:

    f = P.f

    inflow = 0.0

    if s_index == 0:  # (0,2)  Hh=K1, Hm=K3

        for K0 in (0,3):

            for r_index, K in enumerate(KS):

                if K[0]==K0 and K[1]==Hh and K[3]==Hm:

                    inflow += G_pref[(1, r_index)] * J[1] * R[r_index] * f[(1,0)]

        for K2 in (0,3):

            for r_index, K in enumerate(KS):

                if K[1]==Hh and K[2]==K2 and K[3]==Hm:

                    inflow += G_pref[(2, r_index)] * J[2] * R[r_index] * f[(3,2)]

        for K0 in (0,3):

            for K2 in (0,3):

                for r_index, K in enumerate(KS):
```

```
            if K[0]==K0 and K[1]==Hh and K[2]==K2 and K[3]==Hm:
                inflow += G_pref[(3, r_index)] * J[3] * R[r_index] * f[(1,0)] * f[(3,2)]
    return inflow

if s_index == 1:  # (1,2)  Hh=K0, Hm=K3
    for K1 in (0,3):
        for r_index, K in enumerate(KS):
            if K[0]==Hh and K[1]==K1 and K[3]==Hm:
                inflow += G_pref[(0, r_index)] * J[0] * R[r_index] * f[(0,1)]
    for K2 in (0,3):
        for r_index, K in enumerate(KS):
            if K[0]==Hh and K[2]==K2 and K[3]==Hm:
                inflow += G_pref[(3, r_index)] * J[3] * R[r_index] * f[(3,2)]
    for K1 in (0,3):
        for K2 in (0,3):
            for r_index, K in enumerate(KS):
                if K[0]==Hh and K[1]==K1 and K[2]==K2 and K[3]==Hm:
                    inflow += G_pref[(2, r_index)] * J[2] * R[r_index] * f[(0,1)] * f[(3,2)]
    return inflow

if s_index == 2:  # (0,3)  Hh=K1, Hm=K2
    for K0 in (0,3):
        for r_index, K in enumerate(KS):
```

```
            if K[0]==K0 and K[1]==Hh and K[2]==Hm:
                inflow += G_pref[(3, r_index)] * J[3] * R[r_index] * f[(1,0)]
    for K3 in (0,3):
        for r_index, K in enumerate(KS):
            if K[1]==Hh and K[2]==Hm and K[3]==K3:
                inflow += G_pref[(0, r_index)] * J[0] * R[r_index] * f[(2,3)]
    for K0 in (0,3):
        for K3 in (0,3):
            for r_index, K in enumerate(KS):
                if K[0]==K0 and K[1]==Hh and K[2]==Hm and K[3]==K3:
                    inflow += G_pref[(1, r_index)] * J[1] * R[r_index] * f[(1,0)] * f[(2,3)]
    return inflow

if s_index == 3:  # (1,3)  Hh=K0, Hm=K2
    for K3 in (0,3):
        for r_index, K in enumerate(KS):
            if K[0]==Hh and K[2]==Hm and K[3]==K3:
                inflow += G_pref[(1, r_index)] * J[1] * R[r_index] * f[(2,3)]
    for K1 in (0,3):
        for r_index, K in enumerate(KS):
            if K[0]==Hh and K[1]==K1 and K[2]==Hm:
                inflow += G_pref[(2, r_index)] * J[2] * R[r_index] * f[(0,1)]
    for K1 in (0,3):
```

```
            for K3 in (0,3):
                for r_index, K in enumerate(KS):
                    if K[0]==Hh and K[1]==K1 and K[2]==Hm and K[3]==K3:
                        inflow += G_pref[(0, r_index)] * J[0] * R[r_index] * f[(0,1)] * f[(2,3)]
        return inflow

    return inflow

def rhs_with_mutation(t: float, y: np.ndarray, P: Params, G_pref):
    dy = np.zeros_like(y)
    R = y[:NR]
    I = y[NR:]

    J = np.zeros(4)
    for s_index in range(4):
        J[s_index] = np.sum(I[s_index*16:(s_index+1)*16])

    N = float(np.sum(y))
    births = P.mu * N

    for r_index, K in enumerate(KS):
        K0,K1,K2,K3 = K
        out_rate = omega(P.w2,P.w1,0,K0) + omega(P.w2,P.w1,1,K1) + omega(P.w2,P.w1,2,K2) +
omega(P.w2,P.w1,3,K3)
```

```
    W = -out_rate * R[r_index]
    if K0 <= 2:
        fromK = (K0+1,K1,K2,K3)
        if K0+1<=3: W += omega(P.w2,P.w1,0,K0+1) * y[IDX_R[fromK]]
    if K1 <= 2:
        fromK = (K0,K1+1,K2,K3)
        if K1+1<=3: W += omega(P.w2,P.w1,1,K1+1) * y[IDX_R[fromK]]
    if K2 <= 2:
        fromK = (K0,K1,K2+1,K3)
        if K2+1<=3: W += omega(P.w2,P.w1,2,K2+1) * y[IDX_R[fromK]]
    if K3 <= 2:
        fromK = (K0,K1,K2,K3+1)
        if K3+1<=3: W += omega(P.w2,P.w1,3,K3+1) * y[IDX_R[fromK]]

    L_in = L_of_K(y, P, K)

    G_out = 0.0
    for s_index in range(4):
        G_out += G_pref[(s_index, r_index)] * J[s_index] * R[r_index]

    dy[r_index] = L_in + W - P.mu*R[r_index] - G_out

dy[S_INDEX] += births
```

```
    for s_index in range(4):
        rec = P.v[STRAINS[s_index]]
        for Hh in range(4):
            for Hm in range(4):
                i_idx = idx_I(s_index, Hh, Hm)
                inflow_same = 0.0
                for r_index in BASE_R_FOR_I[(s_index,Hh,Hm)]:
                    inflow_same += G_pref[(s_index, r_index)] * J[s_index] * R[r_index]
                inflow_same *= psi_factor(P, s_index, Hh, Hm)
                inflow_cross = C_inflow(y, R, J, P, G_pref, s_index, Hh, Hm)
                dy[i_idx] = inflow_same + inflow_cross - rec*y[i_idx] - P.mu*y[i_idx]
    return dy

# ----------------------------
# Simulation and utilities
# ----------------------------
def simulate(P: Params):
    _, G_pref = precompute_susc_and_Gpref(P)
    y0 = np.zeros(NSTATE)
    y0[S_INDEX] = 0.99999
    y0[idx_I(0,3,3)] = 1e-5
    t_eval = np.linspace(0.0, P.tmax, P.t_points)
```

```
    sol = solve_ivp(lambda t, y: rhs_with_mutation(t, y, P, G_pref),
                    (0.0, P.tmax), y0, t_eval=t_eval, method='LSODA',
                    rtol=P.rtol, atol=P.atol, max_step=np.inf)
    if not sol.success:
        raise RuntimeError(f"Integrator failed: {sol.message}")
    return sol

def J_of_solution(sol, s_index: int):
    Jvals = []
    for k in range(sol.y.shape[1]):
        yk = sol.y[:,k]
        block = yk[NR + s_index*16 : NR + (s_index+1)*16]
        Jvals.append(np.sum(block))
    return np.array(Jvals)

def compute_all_J(sol):
    return {
        "(0,2)": J_of_solution(sol, 0),
        "(1,2)": J_of_solution(sol, 1),
        "(0,3)": J_of_solution(sol, 2),
        "(1,3)": J_of_solution(sol, 3),
    }
```

```
def report_peaks(sol, Jdict):
    print("\nPeak values (global maxima) and times:")
    for key in ["(0,2)","(1,2)","(0,3)","(1,3)"]:
        series = Jdict[key]
        idx = int(np.argmax(series))
        print(f"  {key}: peak={series[idx]:.8f} at t={sol.t[idx]:.3f}")

def plot_all_four(sol, Jdict, title: str = "Strain totals over time", save_path=None, t_le=None, show=True):
    """Plot all four strain totals and optionally auto-save a PNG figure.

    This function deliberately saves PNG only. On some Mac/Anaconda/Spyder
    installations, matplotlib PDF export can fail because of fontTools
    permissions; PNG export avoids that backend.
    """
    if t_le is not None:
        mask = sol.t <= t_le
        t_plot = sol.t[mask]
        Jplot = {k: v[mask] for k, v in Jdict.items()}
    else:
        t_plot = sol.t
        Jplot = Jdict

    fig, ax = plt.subplots(figsize=(8, 5))
```

```
# Strong, simple colors suitable for screen, Word, and submission review.
ax.plot(t_plot, Jplot["(0,2)"], label="J(0,2)", color="black", linewidth=2.6, linestyle="-")
ax.plot(t_plot, Jplot["(1,3)"], label="J(1,3)", color="red", linewidth=2.6, linestyle="-")
ax.plot(t_plot, Jplot["(1,2)"], label="J(1,2)", color="blue", linewidth=2.1, linestyle="--")
ax.plot(t_plot, Jplot["(0,3)"], label="J(0,3)", color="green", linewidth=2.1, linestyle="--")

ax.set_xlabel("Time (days)", fontsize=12)
ax.set_ylabel("Infected fraction", fontsize=12)
ax.set_title(title, fontsize=13)
ax.legend(loc="upper right", frameon=False, fontsize=10)
ax.spines["top"].set_visible(False)
ax.spines["right"].set_visible(False)
ax.tick_params(axis="both", labelsize=10)
fig.tight_layout()

if save_path is not None:
    save_path = Path(save_path)
    # Force PNG even if a .pdf name is accidentally supplied.
    if save_path.suffix.lower() != ".png":
        save_path = save_path.with_suffix(".png")
    save_path.parent.mkdir(parents=True, exist_ok=True)
    fig.savefig(str(save_path), dpi=300, bbox_inches="tight", format="png")
```

```
        print(f"Saved PNG plot:\n  {save_path}")

    if show:
        plt.show()
    else:
        plt.close(fig)

# CLI
# ----------------------------
def parse_args():
    ap = argparse.ArgumentParser(description="Allele-based 2-site model; auto-saves PNG plot")
    ap.add_argument("--no-mutation", action="store_true", help="Set all drift f_ij = 0")
    ap.add_argument("--f01", type=float, default=None, help="Drift 0->1")
    ap.add_argument("--f10", type=float, default=None, help="Drift 1->0")
    ap.add_argument("--f23", type=float, default=None, help="Drift 2->3")
    ap.add_argument("--f32", type=float, default=None, help="Drift 3->2")

    ap.add_argument("--beta0", type=float, default=None)
    ap.add_argument("--beta1", type=float, default=None)
    ap.add_argument("--beta2", type=float, default=None)
    ap.add_argument("--beta3", type=float, default=None)

    ap.add_argument("--m0", type=float, default=None)
```

```
ap.add_argument("--m1", type=float, default=None)
ap.add_argument("--m2", type=float, default=None)
ap.add_argument("--m3", type=float, default=None)

ap.add_argument("--nu0", type=float, default=None)
ap.add_argument("--nu1", type=float, default=None)
ap.add_argument("--nu2", type=float, default=None)
ap.add_argument("--nu3", type=float, default=None)

# ADE parameters: additive contribution from immune-history allele a.
ap.add_argument("--d0", type=float, default=None)
ap.add_argument("--d1", type=float, default=None)
ap.add_argument("--d2", type=float, default=None)
ap.add_argument("--d3", type=float, default=None)

# Optional allele-pair epistasis terms.
ap.add_argument("--e02", type=float, default=None)
ap.add_argument("--e12", type=float, default=None)
ap.add_argument("--e03", type=float, default=None)
ap.add_argument("--e13", type=float, default=None)

ap.add_argument("--c", type=float, default=None, help="Contact rate")
ap.add_argument("--mu", type=float, default=None, help="Birth/death rate")
```

```
    ap.add_argument("--tmax", type=float, default=None, help="Final time for integration")
    ap.add_argument("--points", type=int, default=None, help="Number of time samples")
    ap.add_argument("--rtol", type=float, default=None)
    ap.add_argument("--atol", type=float, default=None)
    ap.add_argument("--tle", type=float, default=None, help="Only plot/export up to this time (t ≤ tle)")

    ap.add_argument("--png", type=str, default=None, help="Path to save PNG plot; default is allele_plot.png beside this script")
    ap.add_argument("--no-save", action="store_true", help="Do not auto-save plot files")
    ap.add_argument("--no-show", action="store_true", help="Do not display an on-screen plot")

    ap.add_argument("--report-peaks", action="store_true", help="Print peak value and time for each J(i,j)")

    return ap.parse_args()

def main():
    P = default_params()
    args = parse_args()

    if args.no_mutation:
        P.f = {(0,1):0.0,(1,0):0.0,(2,3):0.0,(3,2):0.0}
    if args.f01 is not None: P.f[(0,1)] = args.f01
```

```
if args.f10 is not None: P.f[(1,0)] = args.f10
if args.f23 is not None: P.f[(2,3)] = args.f23
if args.f32 is not None: P.f[(3,2)] = args.f32

if args.beta0 is not None: P.beta[0]=args.beta0
if args.beta1 is not None: P.beta[1]=args.beta1
if args.beta2 is not None: P.beta[2]=args.beta2
if args.beta3 is not None: P.beta[3]=args.beta3

if args.m0 is not None: P.m[0]=args.m0
if args.m1 is not None: P.m[1]=args.m1
if args.m2 is not None: P.m[2]=args.m2
if args.m3 is not None: P.m[3]=args.m3

if args.nu0 is not None: P.nu[0]=args.nu0
if args.nu1 is not None: P.nu[1]=args.nu1
if args.nu2 is not None: P.nu[2]=args.nu2
if args.nu3 is not None: P.nu[3]=args.nu3

if args.d0 is not None: P.d[0]=args.d0
if args.d1 is not None: P.d[1]=args.d1
if args.d2 is not None: P.d[2]=args.d2
if args.d3 is not None: P.d[3]=args.d3
```

```
if args.e02 is not None: P.e[(0,2)] = args.e02
if args.e12 is not None: P.e[(1,2)] = args.e12
if args.e03 is not None: P.e[(0,3)] = args.e03
if args.e13 is not None: P.e[(1,3)] = args.e13

if args.c is not None: P.c=args.c
if args.mu is not None: P.mu=args.mu

if args.tmax is not None: P.tmax=args.tmax
if args.points is not None: P.t_points=args.points
if args.rtol is not None: P.rtol=args.rtol
if args.atol is not None: P.atol=args.atol

sol = simulate(P)
Jdict = compute_all_J(sol)

print(f"Final J(0,2) at t={P.tmax:.0f}: {Jdict['(0,2)'][-1]:.8f}")
print(f"Final J(1,2) at t={P.tmax:.0f}: {Jdict['(1,2)'][-1]:.8f}")
print(f"Final J(0,3) at t={P.tmax:.0f}: {Jdict['(0,3)'][-1]:.8f}")
print(f"Final J(1,3) at t={P.tmax:.0f}: {Jdict['(1,3)'][-1]:.8f}")
print(f"Total population at t={P.tmax:.0f}: {np.sum(sol.y[:,-1]):.8f}")
```

```
    if args.report_peaks:
        report_peaks(sol, Jdict)

    # Auto-save a PNG plot unless disabled. Defaults save beside this script.
    png_path = None
    if not args.no_save:
        png_path = Path(args.png) if args.png else DEFAULT_OUTPUT_DIR / "allele_plot.png"

    plot_title = "Strain totals over time" if args.tle is None else f"Strain totals over time (t <= {args.tle})"
    plot_all_four(
        sol, Jdict,
        title=plot_title,
        save_path=png_path,
        t_le=args.tle,
        show=(not args.no_show),
    )

if __name__ == "__main__":
    main()
```
